# Dissecting unique and common variance across body and brain health indicators using age prediction

**DOI:** 10.1101/2023.11.03.23298017

**Authors:** Dani Beck, Ann-Marie G. de Lange, Tiril P. Gurholt, Irene Voldsbekk, Ivan I. Maximov, Sivaniya Subramaniapillai, Louise Schindler, Guy Hindley, Esten H. Leonardsen, Zillur Rahman, Dennis van der Meer, Max Korbmacher, Jennifer Linge, Olof D. Leinhard, Karl T. Kalleberg, Andreas Engvig, Ida Sønderby, Ole A. Andreassen, Lars T. Westlye

## Abstract

Ageing is a heterogeneous multisystem process involving different rates of decline in physiological integrity across biological systems. The current study dissects the unique and common variance across body and brain health indicators and parses inter-individual heterogeneity in the multisystem ageing process. Using machine-learning regression models on the UK Biobank dataset (N = 32,593, age range 44.6-82.3, mean age 64.1 years), we first estimated tissue-specific brain age for white and gray matter based on diffusion and T1-weighted magnetic resonance imaging (MRI) data, respectively. Next, bodily health traits including cardiometabolic, anthropometric, and body composition measures of adipose and muscle tissue from bioimpedance and body MRI were combined to predict ‘body age’. The results showed that the body age model demonstrated comparable age prediction accuracy to models trained solely on brain MRI data. The correlation between body age and brain age predictions was 0.62 for the T1 and 0.64 for the diffusion-based model, indicating a degree of unique variance in brain and bodily ageing processes. Bayesian multilevel modelling carried out to quantify the associations between health traits and predicted age discrepancies showed that higher systolic blood pressure and higher muscle-fat infiltration were related to older-appearing body age compared to brain age. Conversely, higher hand-grip strength and muscle volume were related to a younger-appearing body age. Our findings corroborate the common notion of a close connection between somatic and brain health. However, they also suggest that health traits may differentially influence age predictions beyond what is captured by the brain imaging data, potentially contributing to heterogeneous ageing rates across biological systems and individuals.

## 1. Introduction

Ageing has been defined as a multisystem and time-dependent process involving progressive loss of functional and physiological integrity (López-Otín et al., 2013). While advanced age is a primary risk factor for cardiovascular and neurodegenerative diseases, ageing is highly heterogeneous, with differential ageing rates across biological systems and individuals (Cevenini et al., 2008). This has motivated a wealth of research into better understanding the determinants of individual differences in ageing and its relevance to diverse disease processes.

Several biomarkers of ageing including body composition and health traits have been proposed (Cole et al., 2019). Changes in blood lipids, adipose and muscle tissue distribution, blood pressure, heart rate, hand grip strength, and anthropometric measures such as body mass index (BMI) and waist-to-hip ratio (WHR) are all associated with ageing (Massy-Westropp et al., 2011; Mielke et al., 2010; Rodgers et al., 2019; Sebastiani et al., 2017). Despite these measures being classified as markers of normal body function rather than disease-specific biomarkers, recent studies have highlighted their utility for risk detection and disease monitoring across cardiovascular disease and dementia (Brain et al., 2023). For example, research has suggested that dysregulation in lipid metabolism in Alzheimer’s disease may predict cognitive decline (Wong et al., 2017).

Age prediction using machine learning applied to brain magnetic resonance imaging (MRI) data has enabled individual-level age prediction with high accuracy based on brain white (WM) and gray matter (GM) characteristics derived from diffusion and T1-weighted MRI scans (Beck et al., 2021; Cole et al., 2017; Leonardsen et al., 2022), providing neuroanatomical markers of brain health and integrity (Cole & Franke, 2017; Franke et al., 2010). Although bodily health traits have demonstrated their influence on brain ageing (Beck, de Lange, Alnæs, et al., 2022; Beck, de Lange, Pedersen, et al., 2022; de Lange et al., 2020; Franke et al., 2013, 2014; Kolenic et al., 2018; Ronan et al., 2016), rates of brain and body ageing processes may be partly distinct at the individual level.

Recent work has demonstrated the relevance of age prediction based on various organ structures (Tian et al., 2023), reporting that body- and brain-specific age estimates can be differentially influenced by lifestyle and environmental factors. However, a comprehensive understanding of the unique and common variance across body and brain age models, in addition to the contribution of specific bodily health traits, warrants further investigation. For example, bodily health traits such as elevated blood pressure may potentially contribute to a group-level increase in predicted age estimates relative to predictions based solely on brain MRI data. However, due to the impact of elevated blood pressure on the brain (Dintica et al., 2023; George et al., 2023), this variance may already to some extent be captured by brain age models. The present study focuses on parsing inter-individual heterogeneity in the multisystem ageing process by comparing age predictions based on models trained separately on indicators of body and brain health. Specifically, we focus on identifying key health traits that influence age predictions beyond the variance captured by the brain measures.

Using the UK Biobank sample (N = 32,593, mean age = 64.1, SD = 7.5), we assessed the contributions of specific health traits to discrepancies between body and brain age predictions. Bodily health traits included cardiometabolic factors, anthropometric measures, and body composition measures of adipose and muscle tissue from bioimpedance and body MRI. Based on documented connections between brain and body health, we anticipated that age predictions based on bodily health traits would to a large extent resemble predictions from models trained solely on brain MRI data. However, we expected to observe individual variation in the difference between body and brain age, and that this variation would hold relevant information for better understanding the role of specific health traits. Lastly, we assessed the extent to which specific markers of bodily health, such as blood pressure, abdominal adiposity, and muscle volume, contributed to differences in individual age predictions.

## 2. Methodology

### 2.1. Participants and ethical approval

The sample was drawn from the UK Biobank (UKB) (http://www.ukbiobank.ac.uk). All participants provided signed informed consent. UKB has IRB approval from Northwest Multi-centre Research Ethics Committee and its Ethics Advisory Committee (https://www.ukbiobank.ac.uk/ethics) oversees the UKB Ethics & Governance Framework (Miller et al., 2016). Specific details regarding recruitment and data collection procedures have been previously published (Collins, 2007). The present study uses the UKB Resource under Application Number 27412. Participants were excluded from the present study if they reported disorders that affect the brain based on ICD10 diagnoses or a long-standing illness disability, diabetes, or stroke history (N = 210).

To remove poor-quality T1-weighted brain MRI data, participants with Euler numbers (Rosen et al., 2018) ≥ 4 standard deviations below the mean were excluded. For diffusion-weighted (dMRI) data, quality control was assured using the YTTRIUM algorithm (Maximov et al., 2021). A total of N = 160 participants were removed for T1 and dMRI data. Body MRI measurements were quality checked by two independent, trained operators visually inspecting the images prior to upload to UKB and this has been followed by control of all outliers for anatomical correctness.

For health traits (hereby used as an umbrella term including all body composition and health markers, outliers (values ± 5 SD from the mean) were excluded (N = 627) from the analysis by converting the values to NA, thereby keeping the participant in the sample with their respective non-outlier data. SI Figures 1 and 2 show distributions of health traits before and after quality control. SI Figure 3 shows the prevalence of NA/missing data in the final sample. Following cleaning, the final sample consisted of 32,593 individuals (Females: N = 17,200, mean age = 63.6, SD = 7.37, Males: N = 15,393, mean age = 64.71, SD = 7.63) with T1, dMRI, and body health trait data. Table 1 summarises the health trait descriptive statistics.

**Table 1.**
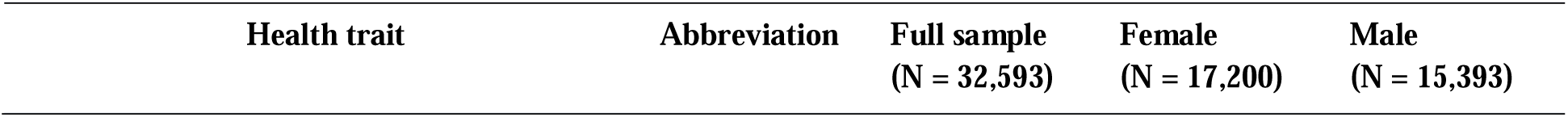
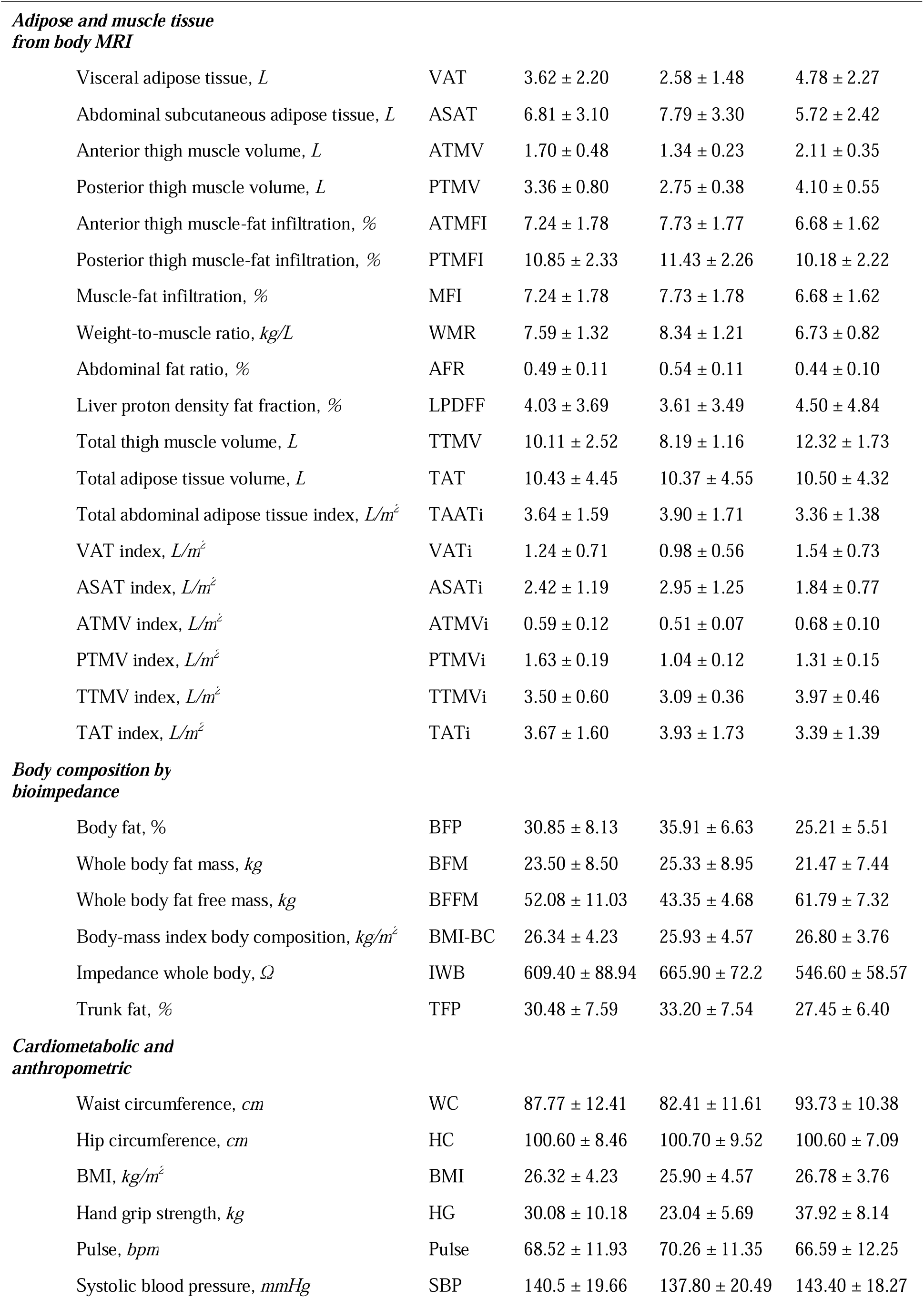
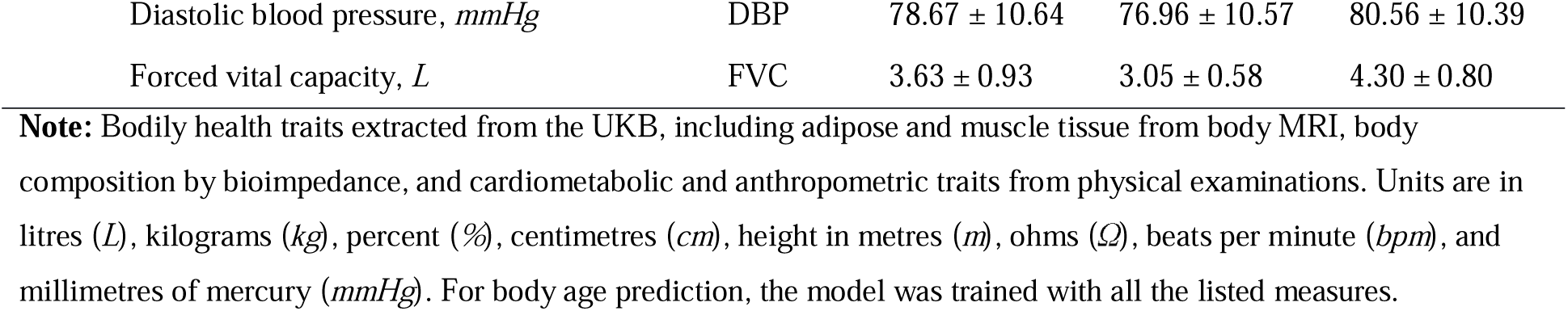
Descriptive statistics pertaining to each health trait, including mean ± standard deviation (SD).

| Health trait | Abbreviation | Full sample<br>(N = 32,593) | Female<br>(N = 17,200) | Male<br>(N = 15,393) |
| --- | --- | --- | --- | --- |
| **Adipose and muscle tissue from body MRI** |  |  |  |  |
| Visceral adipose tissue, *L* | VAT | 3.62 ± 2.20 | 2.58 ± 1.48 | 4.78 ± 2.27 |
| Abdominal subcutaneous adipose tissue, *L* | ASAT | 6.81 ± 3.10 | 7.79 ± 3.30 | 5.72 ± 2.42 |
| Anterior thigh muscle volume, *L* | ATMV | 1.70 ± 0.48 | 1.34 ± 0.23 | 2.11 ± 0.35 |
| Posterior thigh muscle volume, *L* | PTMV | 3.36 ± 0.80 | 2.75 ± 0.38 | 4.10 ± 0.55 |
| Anterior thigh muscle-fat infiltration, % | ATMFI | 7.24 ± 1.78 | 7.73 ± 1.77 | 6.68 ± 1.62 |
| Posterior thigh muscle-fat infiltration, % | PTMFI | 10.85 ± 2.33 | 11.43 ± 2.26 | 10.18 ± 2.22 |
| Muscle-fat infiltration, % | MFI | 7.24 ± 1.78 | 7.73 ± 1.78 | 6.68 ± 1.62 |
| Weight-to-muscle ratio, *kg/L* | WMR | 7.59 ± 1.32 | 8.34 ± 1.21 | 6.73 ± 0.82 |
| Abdominal fat ratio, % | AFR | 0.49 ± 0.11 | 0.54 ± 0.11 | 0.44 ± 0.10 |
| Liver proton density fat fraction, % | LPDFF | 4.03 ± 3.69 | 3.61 ± 3.49 | 4.50 ± 4.84 |
| Total thigh muscle volume, *L* | TTMV | 10.11 ± 2.52 | 8.19 ± 1.16 | 12.32 ± 1.73 |
| Total adipose tissue volume, *L* | TAT | 10.43 ± 4.45 | 10.37 ± 4.55 | 10.50 ± 4.32 |
| Total abdominal adipose tissue index, *L/m<sup>2</sup>* | TAATi | 3.64 ± 1.59 | 3.90 ± 1.71 | 3.36 ± 1.38 |
| VAT index, *L/m<sup>2</sup>* | VATi | 1.24 ± 0.71 | 0.98 ± 0.56 | 1.54 ± 0.73 |
| ASAT index, *L/m<sup>2</sup>* | ASATi | 2.42 ± 1.19 | 2.95 ± 1.25 | 1.84 ± 0.77 |
| ATMV index, *L/m<sup>2</sup>* | ATMVi | 0.59 ± 0.12 | 0.51 ± 0.07 | 0.68 ± 0.10 |
| PTMV index, *L/m<sup>2</sup>* | PTMVi | 1.63 ± 0.19 | 1.04 ± 0.12 | 1.31 ± 0.15 |
| TTMV index, *L/m<sup>2</sup>* | TTMVi | 3.50 ± 0.60 | 3.09 ± 0.36 | 3.97 ± 0.46 |
| TAT index, *L/m<sup>2</sup>* | TATi | 3.67 ± 1.60 | 3.93 ± 1.73 | 3.39 ± 1.39 |
| **Body composition by bioimpedance** |  |  |  |  |
| Body fat, % | BFP | 30.85 ± 8.13 | 35.91 ± 6.63 | 25.21 ± 5.51 |
| Whole body fat mass, *kg* | BFM | 23.50 ± 8.50 | 25.33 ± 8.95 | 21.47 ± 7.44 |
| Whole body fat free mass, *kg* | BFFM | 52.08 ± 11.03 | 43.35 ± 4.68 | 61.79 ± 7.32 |
| Body-mass index body composition, *kg/m<sup>2</sup>* | BMI-BC | 26.34 ± 4.23 | 25.93 ± 4.57 | 26.80 ± 3.76 |
| Impedance whole body, $\Omega$ | IWB | 609.40 ± 88.94 | 665.90 ± 72.2 | 546.60 ± 58.57 |
| Trunk fat, % | TFP | 30.48 ± 7.59 | 33.20 ± 7.54 | 27.45 ± 6.40 |
| **Cardiometabolic and anthropometric** |  |  |  |  |
| Waist circumference, *cm* | WC | 87.77 ± 12.41 | 82.41 ± 11.61 | 93.73 ± 10.38 |
| Hip circumference, *cm* | HC | 100.60 ± 8.46 | 100.70 ± 9.52 | 100.60 ± 7.09 |
| BMI, *kg/m<sup>2</sup>* | BMI | 26.32 ± 4.23 | 25.90 ± 4.57 | 26.78 ± 3.76 |
| Hand grip strength, *kg* | HG | 30.08 ± 10.18 | 23.04 ± 5.69 | 37.92 ± 8.14 |
| Pulse, *bpm* | Pulse | 68.52 ± 11.93 | 70.26 ± 11.35 | 66.59 ± 12.25 |
| Systolic blood pressure, *mmHg* | SBP | 140.5 ± 19.66 | 137.80 ± 20.49 | 143.40 ± 18.27 |
| Diastolic blood pressure, *mmHg* | DBP | 78.67 ± 10.64 | 76.96 ± 10.57 | 80.56 ± 10.39 |
| Forced vital capacity, *L* | FVC | 3.63 ± 0.93 | 3.05 ± 0.58 | 4.30 ± 0.80 |

**Note:** Bodily health traits extracted from the UKB, including adipose and muscle tissue from body MRI, body composition by bioimpedance, and cardiometabolic and anthropometric traits from physical examinations. Units are in litres (*L*), kilograms (*kg*), percent (%), centimetres (*cm*), height in metres (*m*), ohms ( $\Omega$ ), beats per minute (*bpm*), and millimetres of mercury (*mmHg*). For body age prediction, the model was trained with all the listed measures.

### 2.2. MRI data acquisition and processing

A detailed overview of the full UKB data acquisition and image processing protocol is available in Alfaro et al. (2018) and Miller et al. (2016). Briefly, brain MRI data were acquired on a 3 Tesla Siemens 32-channel Skyra scanner. T1-weighted MPRAGE volumes were both acquired in sagittal orientation at 1x1x1 mm^3^. Processing protocols followed a harmonised analysis pipeline, including automated surface-based morphometry and subcortical segmentation using FreeSurfer version 5.3 (Fischl et al., 2002). A standard set of subcortical and cortical summary statistics were used from FreeSurfer (Fischl et al., 2002), as well as a fine-grained cortical parcellation scheme (Glasser et al., 2016) to extract GM cortical thickness, cortical surface area, and volume for 180 regions of interest per hemisphere. This yielded a total set of 1,118 brain imaging features (360/360/360/38 for cortical thickness/area/volume, as well as cerebellar/subcortical and cortical summary statistics, respectively) that were used as input features in the GM-specific age prediction model in line with recent implementations (de Lange et al., 2019; Kaufmann et al., 2019; Schindler et al., 2022).

For dMRI, a conventional Stejskal-Tanner monopolar spin-echo echo-planar imaging sequence was used with multiband factor 3. Diffusion weightings were 1,000 and 2,000 s/mm^2^ and 50 non-coplanar diffusion directions per each diffusion shell. The spatial resolution was 2Lmm^3^ isotropic, and five anterior-posterior versus three anterior-posterior images with *b* =L0 s/mm^2^ were acquired. Data were processed using a previously described pipeline (Maximov et al., 2019). Metrics derived from diffusion tensor imaging (DTI) (Basser, 1995), diffusion kurtosis imaging (DKI) (Jensen et al., 2005), WM tract integrity (WMTI) (Fieremans et al., 2011), and spherical mean technique (SMT) (Kaden et al., 2016) were used as input features in the WM-specific age prediction model, as described in Voldsbekk et al. (2021). Tract-based spatial statistics (TBSS) was used to extract diffusion metrics in WM (Smith et al., 2006) (see Voldsbekk et al. (2021) for full pipeline). For each metric, WM features were extracted based on John Hopkins University (JHU) atlases for WM tracts and labels (with 0 thresholding) (Mori et al., 2006), yielding a total of 910 WM features, including mean values and regional measures for each of the diffusion model metrics, which were used as input features in the WM-specific age prediction model (Beck et al., 2021; Subramaniapillai et al., 2022; Voldsbekk et al., 2021). The diffusion MRI data passed TBSS post-processing quality control using the YTTRIUM algorithm (Maximov et al., 2020), and were residualised with respect to scanning site using linear models.

The methods used to generate the body MRI-derived measurements have been described and evaluated in more detail elsewhere (Borga et al., 2018, 2020; Karlsson et al., 2015; Linge et al., 2018; West et al., 2018). Briefly, the process for fat and muscle compartments includes the following steps: (1) calibration of fat images using fat-referenced MRI, (2) registration of atlases with ground truth labels for fat and muscle compartments to the acquired MRI dataset to produce automatic segmentation, (3) quality control by two independent trained operators including the possibility to adjust and approve the final segmentation, and (4) quantification of fat volumes, muscle volumes and muscle-fat infiltration within the segmented regions. For liver proton density fat fraction (PDFF), nine regions of interest (ROI) were manually placed, evenly distributed in the liver volume, while avoiding major vessels and bile ducts. The total set of features included in the body age prediction model included 40 variables.

### 2.3. Body composition and health traits

Table 1 summarises the descriptive statistics of the health traits used in the study. A detailed description of each variable can be found in Supplementary Information (SI) Section 1.

### 2.4. Age prediction models

Age prediction was carried out using XGBoost regression (eXtreme Gradient Boosting; https://github.com/dmlc/xgboost) in Python 3.8.0, which is based on a decision-tree ensemble algorithm (Chen & Guestrin, 2016) used in several recent age prediction studies (Beck, de Lange, Alnæs, et al., 2022; Beck, de Lange, Pedersen, et al., 2022; Beck et al., 2021; de Lange et al., 2020; Kaufmann et al., 2019; Subramaniapillai et al., 2022; Voldsbekk et al., 2021). XGboost uses advanced regularisation to reduce overfitting, has shown superior performance in machine learning competitions (Chen & Guestrin, 2016), and accommodates the combination of health traits and brain imaging features based on FreeSurfer and FSL-derived values.

Age prediction models were first run using only brain MRI data for WM and GM features separately. Next, we ran a prediction model combining all health traits (Table 1). This resulted in three age prediction models used in the current study: T1 brain age, dMRI brain age, and body age. Parameters were tuned in nested cross-validations with five inner folds for randomised search and 10 outer folds for validating model performance using Scikit-learn (Pedregosa et al., 2011). R^2^, root mean squared error (RMSE), mean absolute error (MAE), and Pearson’s correlations between predicted and true values were calculated to evaluate prediction accuracy. For each model, 10-fold cross-validation was used to obtain brain age and body age for each individual in the full sample. SI Figure 4 shows the feature importance (weight) of the variables in each age prediction model, indicating the relative contribution of the corresponding feature to the prediction model. SI Table 1 shows each of the variables included in the prediction models.

**Figure 4.**
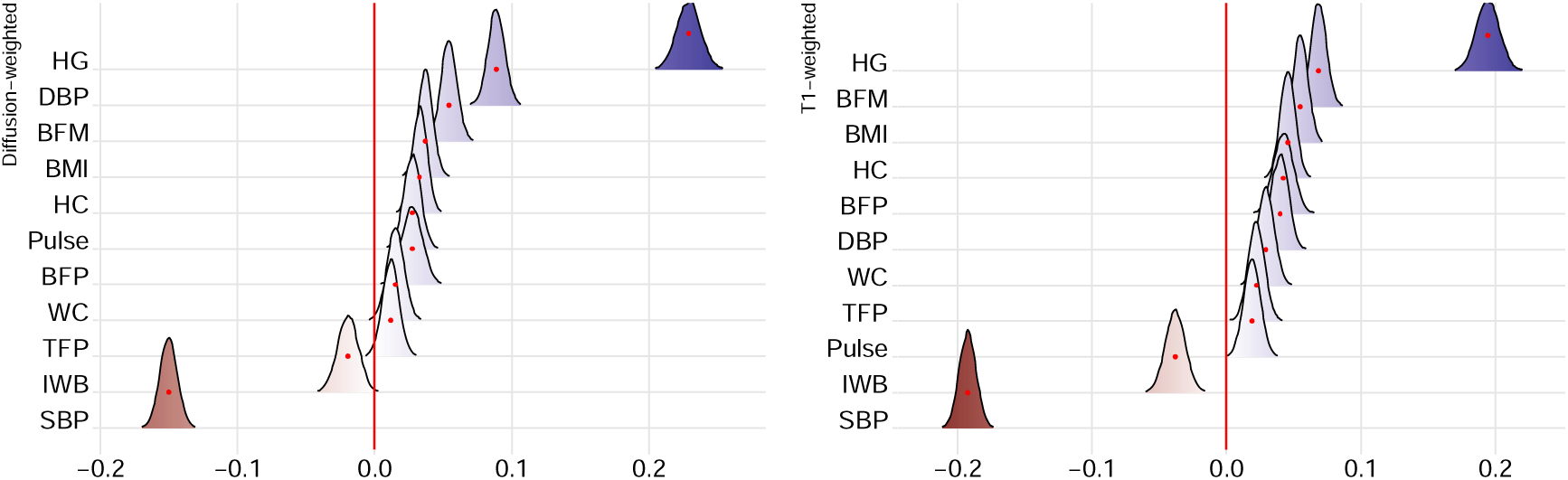
Associations between cardiometabolic, anthropometric, and bioimpedance traits and difference score between brain age models (dMRI and T1) and body age model. The figure shows posterior distributions of the estimates of the standardised coefficient. Estimates for each health trait on dMRI difference score on the left and T1-weighted difference score on the right.

### 2.5. Difference in age predictions by bodily health traits

To investigate the degree to which each of the health traits influenced differences in body and brain age predictions, Bayesian multilevel models were carried out in *Stan* (Stan Development Team, 2023) using the *brms* (Bürkner, 2017, 2018) R-package, where multivariate models are fitted in familiar syntax to comparable frequentist approaches such as a linear mixed effects model using the *lme4* (Bates et al., 2015). We assessed the relationships between age prediction difference scores (predicted brain age minus predicted body age) and each health trait (bar body MRI index scores to reduce redundancy). For each individual, a positive difference score reflects a brain age that is higher than body age, while a negative difference score reflects a body age that is higher than brain age. The difference score (for T1 and dMRI separately) was entered as the dependent variable, with each health trait separately entered as the independent fixed effects variable along with age and sex, with subject ID as the random effect:

Age prediction difference score ∼ Health trait + Age + Sex (1|SubjectID) (1)

To prevent false positives and to regularise the estimated associations, we defined a standard prior around zero with a standard deviation of 0.3 for all coefficients, reflecting a baseline expectation of effects being small but allowing for sufficient flexibility in estimation. All variables bar sex were standardised before running the analyses. Each model was run with 8000 iterations, including 4000 warmup iterations, across four chains. This setup was chosen to ensure robust convergence and adequate sampling from the posterior distributions. For each coefficient of interest, we report the mean estimated value of the posterior distribution (β) and its 95% credible interval (the range of values that with 95% confidence contains the true value of the association), and calculated the Bayes Factor (BF) – provided as evidence ratios in the presented figures – using the Savage-Dickey method (Wagenmakers et al., 2010). Briefly, BF can be interpreted as a measure of the strength of evidence (*extreme, very strong, strong, moderate, anecdotal, none*) in favour of the null or alternative hypothesis. For a pragmatic guide on BF interpretation, see SI Table 2.

## 3. Results

### 3.1. Brain age and body age prediction

Table 2 summarises descriptive and model validation statistics pertaining to each age prediction model. Figure 1 shows the distributions of the age prediction difference scores (predicted T1/dMRI brain age minus predicted body age). SI Figure 5 shows the correlation between the T1 and dMRI-based brain age versus body age difference scores. Figure 2 shows a correlation matrix including the three models’ predicted ages and age gaps. See SI Figure 6 for correlation matrix showing the association between health traits and SI Figure 7 for predicted age as a function of chronological age for each age prediction model.

**Figure 1.**
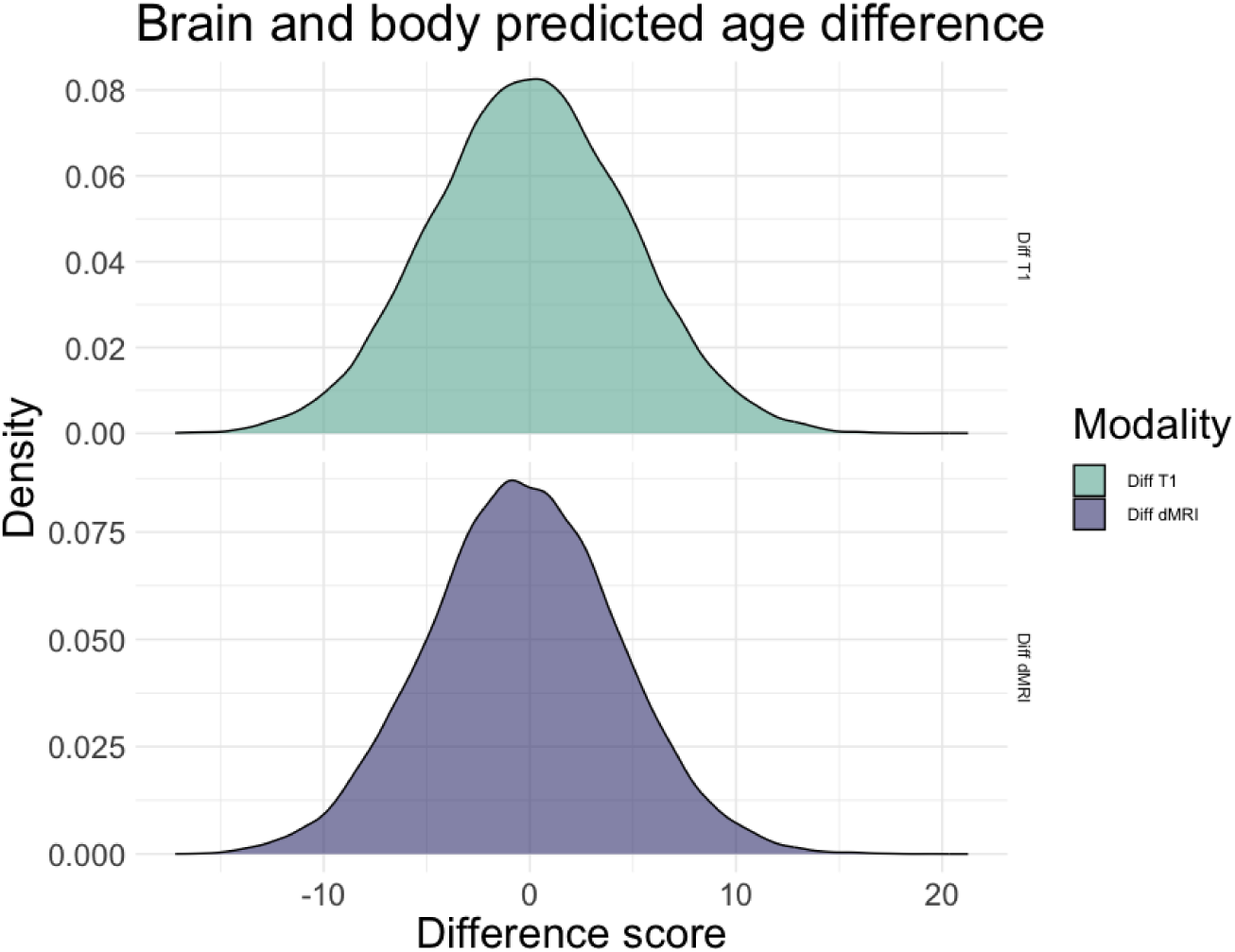
Age difference score distribution (density) for T1 (Diff T1) and dMRI (Diff dMRI) weighted age models. Mean difference scores = 0.028 (T1) and -4.33 (dMRI), with standard deviation (SD) of 4.78 and 4.58, respectively.

**Figure 2.**
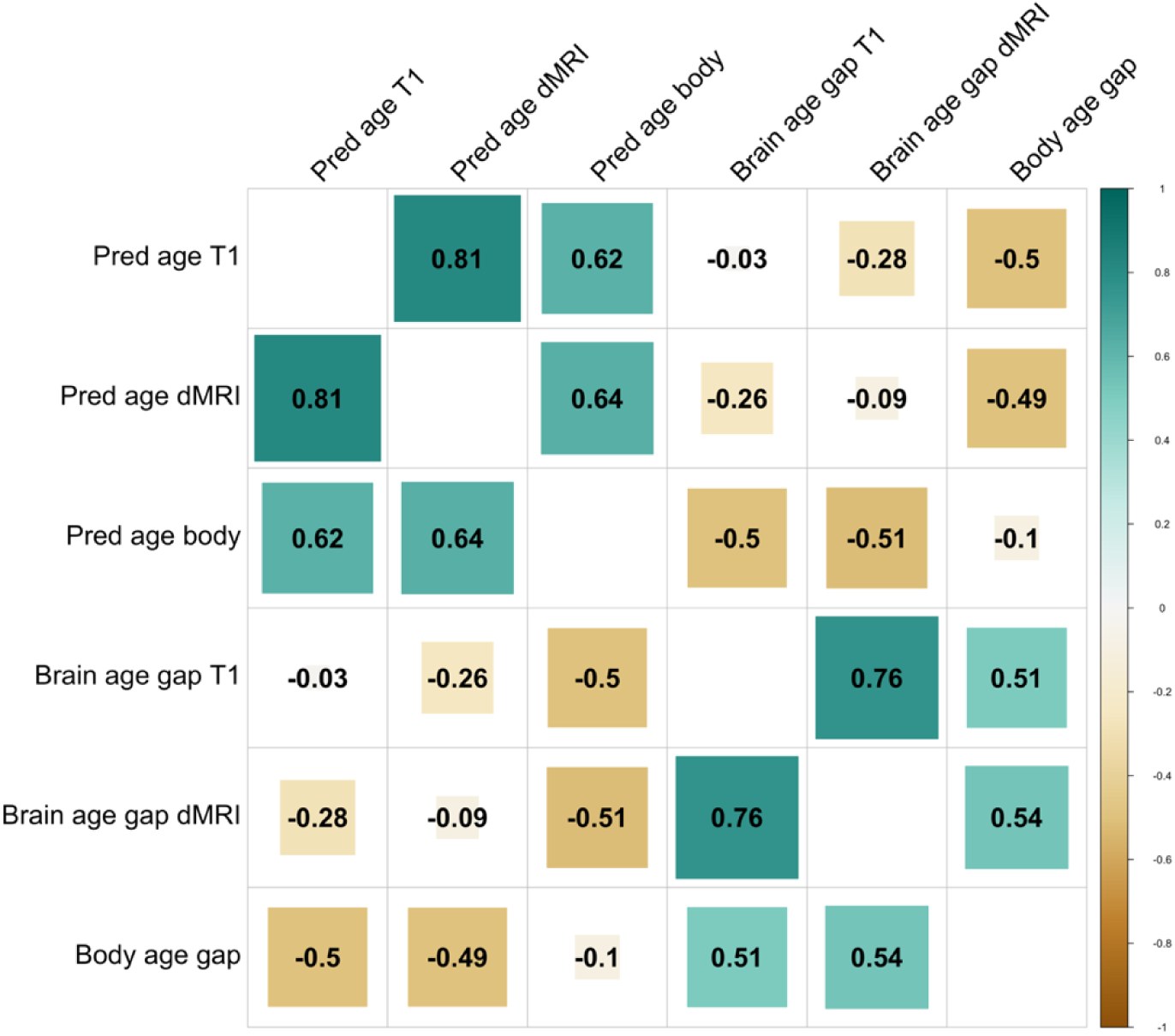
Correlation matrix showing the associations between the predicted ages and age gaps of the three models.

**Table 2.**
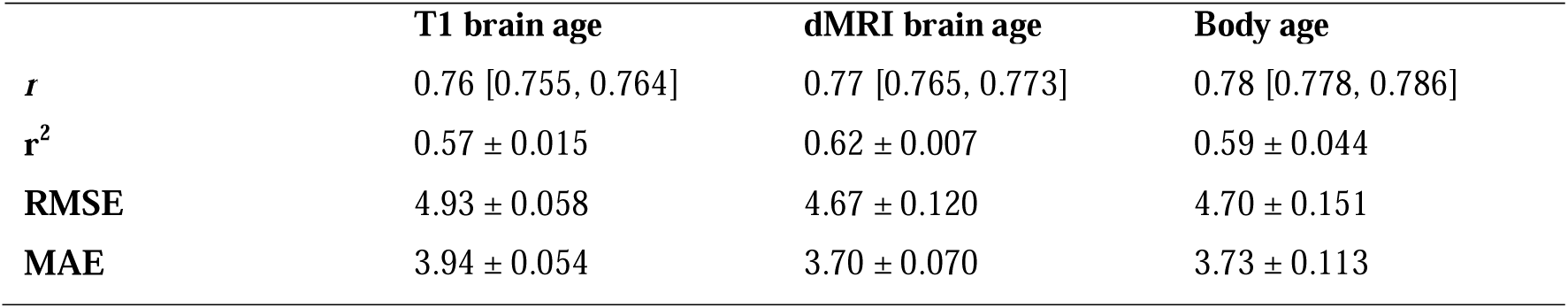
Average R^2^, root mean square error (RMSE), mean absolute error (MAE) standard deviation, and Pearson’s correlations between predicted and true age (*r*) for each age prediction model. 95% confidence intervals on *r* are calculated using Fisher’s z-transformation, adjusted for sample size, and back-transformed to the original scale.

### 3.2. Bayesian multilevel models

Bayesian multilevel modelling tested the associations between each bodily health trait and the difference score (brain-predicted age minus body-predicted age). Due to the large number of included health traits, we present associations between 1) age prediction difference and body MRI measures and 2) age prediction difference and cardiometabolic, anthropometric and bioimpedance measures separately below. The full results are available in SI Tables 3 and 4. For estimated credible intervals, see SI Figures 8 and 9. See SI Table 5 for full and partial linear regressions between age and each of the health trait adjusted for brain-predicted age. See SI Figures 10 and 11 for scatterplots and Pearson’s R reflecting each of the associations between health traits and difference scores between brain age and body age models and SI Figure 12 for scatterplots reflecting associations between health traits and body age gap. Sensitivity analyses using linear mixed effects models showing the results with age-bias corrected scores versus uncorrected scores but with age as a covariate are provided in SI Figures 13 and 14.

#### 3.2.1. Difference scores and body MRI features

Figure 3 shows posterior distributions reflecting the associations between each body MRI feature and the age prediction difference scores. Values increasing from 0 to 1 show evidence of a positive association (where higher values on health traits relate to lower body age relative to brain age) and values decreasing 0 to -1 show evidence of a negative association (where higher values on health traits relate to higher body age relative to brain age).

**Figure 3.**
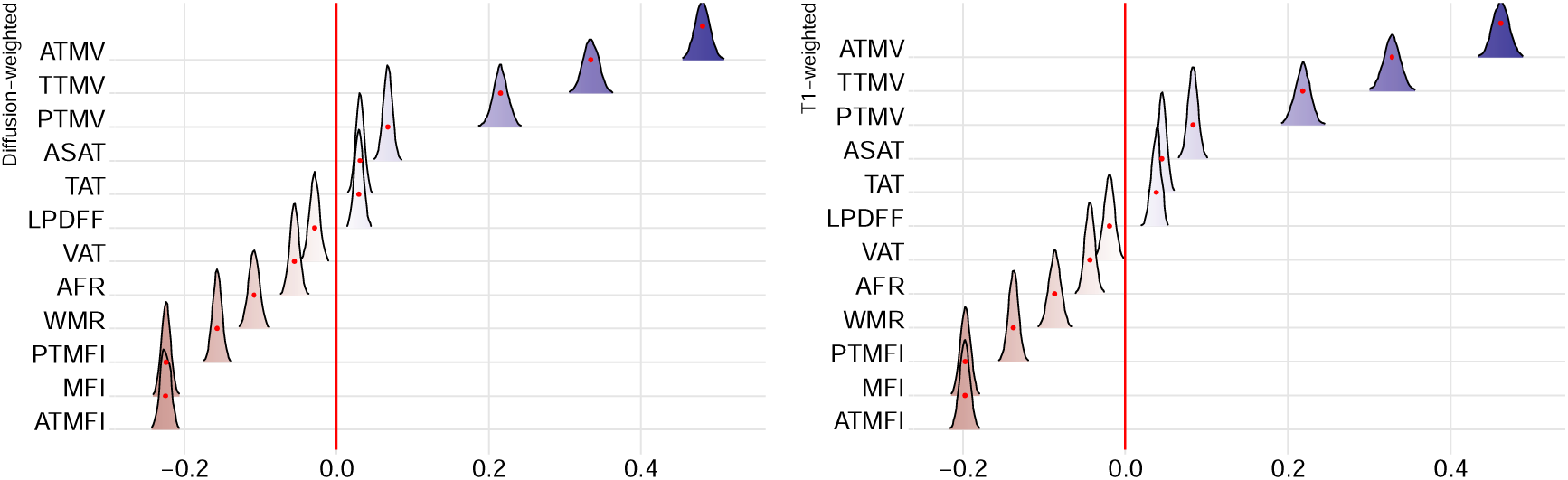
Associations between body MRI features and difference scores between brain-age models and body-age models (dMRI and T1). The figure shows posterior distributions of the estimates of the standardised coefficient. Estimates for each body MRI feature on dMRI difference score on the left and T1-weighted difference score on the right. Colour scale follows the directionality of evidence, with positive (blue) values indicating evidence in favour of positive associations (i.e., larger brain than body age) and negative (red) values indicating evidence in favour of negative associations (i.e., larger body than brain age). The width of the distribution represents the uncertainty of the parameter estimates. For a list of unabbreviated words, see Table 1.

For both dMRI and T1 age difference scores, the tests revealed evidence of a positive association with predicted age difference score (calculated as brain age – body age) for ATMV (dMRI: BF < 0.001, β *=* 0.48; T1: BF < 0.001, β *=* 0.46), TTMV (dMRI: BF < 0.001, β *=* 0.33; T1: BF < 0.001, β *=* 0.33), PTMV (dMRI: BF < 0.001, β *=* 0.22; T1: BF < 0.001, β *=* 0.22), ASAT (dMRI: BF < 0.001, β *=* 0.07; T1: BF < 0.001, β *=* 0.08), TAT (dMRI: BF < 0.001, β *=* 0.03; T1: BF < 0.001, β *=* 0.05), and LPDFF (dMRI: BF < 0.001, β *=* 0.03; T1: BF < 0.001, β *=* 0.04), indicating that higher levels of muscle volume in the thighs, especially anterior, is associated with a positive difference score (i.e., higher predicted brain age than body age; high muscle volume relations with younger-appearing body ageing).

The tests also revealed evidence of a negative association with predicted age difference score for ATMFI (dMRI: BF < 0.001, β *= -*0.23; T1: BF < 0.001, β *= -*0.20), PTMFI (dMRI: BF < 0.001, β *= -*0.16; T1: BF < 0.001, β *= -*0.14), MFI (dMRI: BF < 0.001, β *= -0*.23; T1: BF < 0.001, β *= -*0.20), WMR (dMRI: BF < 0.001, β *= -0*.11; T1: BF < 0.001, β *= -*0.08), and AFR (dMRI: BF < 0.001, β *= -*0.06; T1: BF < 0.001, β *= -*0.04), indicating that higher fat infiltration in the muscles, especially the anterior thighs, were associated with a negative difference score (i.e., higher predicted body age than brain age). There was also evidence of a negative association for dMRI VAT (BF < 0.001, β *= -*0.03) and no association for T1 VAT (BF = 0.30, β *= -*0.02).

#### 3.2.2. Difference scores and cardiometabolic, anthropometric and bioimpedance

Figure 4 shows posterior distributions reflecting the associations between cardiometabolic, anthropometric, and bioimpedance traits and the age prediction difference scores.

For both dMRI and T1 age difference scores, the tests revealed evidence of a positive association with age prediction difference scores for HC (dMRI: BF < 0.001, β *=* 0.03; T1: BF < 0.001, β *=* 0.05), DBP (dMRI: BF < 0.001, β *=* 0.09; T1: BF < 0.001, β *=* 0.04), BFP (dMRI: BF = 0.05, β *=* 0.03; T1: BF < 0.001, β *=* 0.04), HG (dMRI: BF < 0.001, β *=* 0.23; T1: BF < 0.001, β *=* 0.19), BFM (dMRI: BF < 0.001, β *=* 0.05; T1: BF < 0.001, β *=* 0.07), and BMI (dMRI: BF < 0.001, β *=* 0.04; T1: BF < 0.001, β *=* 0.06). There was also a positive association for dMRI Pulse (BF = 0.006, β *=* 0.03), T1 WC (BF < 0.001, β *=* 0.03), and T1 TFP (BF = 0.015, β *=* 0.02), but not for corresponding T1 Pulse (BF = 0.455, β *=* 0.02), dMRI WC (BF < 2.798, β *=* 0.02), and dMRI TFP (BF < 7.783, β *=* 0.01).

In terms of negative associations with age prediction difference scores, effects were found for SBP for both modalities (dMRI: BF < 0.001, β *= -*0.15; T1: BF < 0.001, β *= -*0.19), and T1 IWB (BF < 0.001, β *= -*0.04), but not dMRI IWB (BF = 1.226, β *= -*0.02). The results indicate that high levels of various measures of adiposity and hand grip strength are associated with a higher brain age than body age, with beta coefficients showing the strongest effect for hand grip strength. Systolic blood pressure is implicated as the largest contributor to higher body predicted age.

## 4. Discussion

Evidence of differential ageing rates across different biological systems in the same individual (Cevenini et al., 2008) has led us to conceptualise ageing as a mosaic and heterogeneous construct. One implication is that individual biomarkers studied in isolation may not accurately reflect risk of disease or outcome (Sebastiani et al., 2017), and the use of multiple models in coherence has been recommended (Cevenini et al., 2008; Cole et al., 2019; Kuo et al., 2021). Our analyses revealed that an age prediction model trained on bodily health traits rendered comparably high prediction accuracy compared to the models trained solely on brain MRI data. However, only moderate correlations between body age and brain age predictions were found, indicating a degree of unique variance in brain and bodily ageing processes. Multilevel modelling showed that several elevated body health risk traits differentially contributed to a group level increase or decrease in predicted age, potentially revealing unique and common influences of bodily health traits on body and brain ageing systems.

We ran Bayesian multilevel modelling to quantify the associations between the predicted age difference score and each of the health traits. The results are largely in line with our expectations of measures related to poorer and better somatic health having differential contributions to our age models, whereby poor health is manifested as older-appearing body age and better health as younger-appearing body age. In parsing unique and common influences of different health traits on brain and bodily ageing, we interpret traits with moderate effects on the age prediction difference scores as likely having more of a unique contribution to brain or bodily ageing. For example, the results indicate that thigh muscle volume and hand grip strength showed a larger effect on younger-appearing body ages relative to brain ages, as compared to e.g., liver fat, subcutaneous fat, diastolic blood pressure, BMI, hip and waist circumference, which showed negligible beta coefficients (see Figures 3 and 4 and SI Tables 3 and 4). The latter measures may thus represent health traits that are less likely to uniquely influence brain or bodily ageing while measures related to muscular fitness have a larger impact on a younger body age. Conversely, muscle-fat infiltration and systolic blood pressure have a larger effect on older-appearing body ages relative to brain ages.

However, some findings remain difficult to interpret. For example, we found opposite associations for systolic versus diastolic blood pressure. Previous research has also reported inconsistent effects, with a UKB study reporting higher SBP and DBP associations with greater and lower risk of dementia respectively (Gong et al., 2021). There are also several adiposity-related health traits with positive associations (indicating older-appearing brain age than body age). While these have negligible beta coefficients, the results may be counterintuitive and, based on an extensive literature on the negative effects of obesity, we would typically expect higher adiposity to be related to older-appearing body age relative to the brain. However, other studies have reported similar findings, for example greater scores on anthropometric measures associated with better health outcomes – often referred to as ‘the obesity paradox’ (Amundson et al., 2010; Tutor et al., 2023). While such findings might be influenced by different adiposity measures used across studies (Bosello & Vanzo, 2021) as well as selection biases (Ba et al., 2012; Masters et al., 2013), future research might focus on variations in body-brain relationships across age, sex, and health status (Kivimäki et al., 2018; Subramaniapillai et al., 2022) to better understand these relationships.

An individual may have a brain-predicted age closely aligned with their chronological age but a body age that exceeds it. One theoretical explanation for this may be the involvement of brain maintenance (Nyberg, 2017), where brain health may to some extent be preserved irrespective of bodily health status through a moderating variable such as good muscular fitness. Furthermore, individual differences in cognitive reserve (Stern, 2009, 2012), or resilience to neuropathological changes typically associated with ageing, could influence differences in age-prediction scores. Future studies might therefore aim to investigate these difference scores in the context of cognitive functions known to change with age, such as memory and reaction time (Grady, 2012), as well as reserve-related mechanisms including education, socioeconomic status, and lifestyle (Anatürk et al., 2021). Although speculative, higher body age than brain age may reflect worsening bodily health that has not yet manifested in the brain, which may represent a window of opportunity for intervention. This emphasises the importance of future longitudinal studies. Moreover, it is important to note that a high brain and body age discrepancy does not necessarily mean an old brain and a young body or vice versa, but rather that the brain age is higher or lower relative to the body age. Hence, both brain and body age could be both higher or lower than individual’s chronological age. Future studies may test the relevance of different brain and body age configurations, with the aim to characterise distinct risk profiles related to neural and cardiometabolic health.

Previous studies have demonstrated that variation in predicted brain age is partly explained by individual differences in body composition and health traits, including abdominal fat (Beck, de Lange, Pedersen, et al., 2022; Schindler et al., 2022; Subramaniapillai et al., 2022), muscle-fat infiltration (Beck, de Lange, Alnæs, et al., 2022), hand-grip strength (Cole et al., 2018; Sanders et al., 2021) and muscle volume (Beck, de Lange, Alnæs, et al., 2022). Our findings support these previous reports, but also suggest that health traits may differentially influence age predictions beyond what is captured by the brain imaging measures. However, it could also be the case that the health trait variables do not influence the estimated age in a specific direction on their own, but rather, that the variation reflects the extent to which the given variable is already integrated into the individual’s brain health. Consequently, the difference score would represent a value indicative of the individual in question rather than providing a generalised insight about the health trait itself. There are also a wide range of related health, lifestyle, and environmental factors that our study did not include such as diet, physical activity, socio-economic status, education, social support, and access to healthcare, which may mitigate the effects of our results or reveal larger discrepancies. Future research might consider these aspects by stratifying samples based on different levels of education and lifestyle behaviours, or adjust for the effects of these variables in follow-up analyses.

Further, the correlation (SI Figure 5) between the difference scores in T1- and dMRI-based models indicate that the difference in predictions are related but not identical. As evidenced by Figures 3 and 4, most of our findings related to difference scores were similar across brain MRI modalities. However, for a few select variables, the results suggest there may be some tissue-specific effects. For example, dMRI-Pulse, T1-WC, T1-TFP revealed positive associations while there were no effects for their counterparts (T1-Pulse, dMRI-WC, dMRI-TFP). Similarly, there were negative associations for dMRI-VAT and T1-IWB but no effects for T1-VAT or dMRI-IWB. While these results could reflect subtle, differential discrepancies between tissue-specific neural ageing processes and body age, it is important to note that these discrepancies between imaging modalities may be trivial, as the reported findings for these variables reflect anecdotal evidence. In addition, the large sample size (∼40k) and power of the study which may lead to even trivial differences and small effects being detected as statistically significant. As such, we focus on the beta coefficients and strength of evidence derived from those values to make more practical conclusions. Future studies may consider more comprehensive investigations of tissue-specific and regional MRI phenotypes that may be uniquely associated with discrepancies in brain and body age predictions.

Some strengths and limitations of the study must be discussed. The UK Biobank offers a rich and comprehensive dataset enriched with health-related information, including lifestyle and health factors utilised in the current study. However, selection biases (Brayne & Moffitt, 2022; Tyrrell et al., 2021) such as overall relatively higher education and overrepresentation by individuals of white European descent makes the sample less representative of the wider population. One argument in favour of recruiting relatively healthy individuals at baseline is that some will develop illnesses over the course of the study period, allowing researchers to track changes over time and identify predictors of health decline and therefore targets for intervention strategies. However, our cross-sectional study may be influenced by healthy-volunteer bias, limiting the representativeness of the sample in terms of cardiovascular risk in midlife and older age. A further limitation of the UK Biobank is the limited age range, with participants involved being between 44-82 years of age. Given the importance of tracking changes over time and the potential differences in how bodily health traits may relate to brain health across the lifespan (Kivimäki et al., 2018), and that this may also vary between males and females (Subramaniapillai et al., 2022), future research should include sex-specific models, wider age ranges, and preferably longitudinal data on more diverse and representative samples.

In terms of age prediction, all three models performed comparably well, both in terms of r values and MAE and RSME. However, while brain-based age models had approximately 1000 features, the body age model only included 40. This variation in features warrants caution in interpreting model differences as being driven strictly by biological mechanisms. That said, while prediction accuracies can improve with a larger number of features included, this is not always the case. For example, in Tian et al. (2023), the least accurate organ-specific model included the largest number of features (n = 33) and the best performing model including 11 features. Similarly, our previous studies show that the age-dependency of features may be more indicative of model performance than the sheer number of features included (Anatürk et al., 2021; de Lange et al., 2020), highlighting the importance of feature relevance over quantity in predictive accuracy. This underscores the need for further research to validate age prediction models related to bodily health and organ systems, assessing the optimal balance and significance of feature number versus their age-related associations.

Although we employed nested cross-validation and k-folding to limit overfitting and generate predictions for the full sample in a held-out manner, our models may not generalise due to the lack of external datasets for validation. Moreover, investigating which body-age features drive the discrepancy between brain and body age predictions, whereby the same set of health traits were used to train the body age model, may introduce circularity. Future research may improve generalisability by including independent samples for validation, as well as more comprehensive approaches to assessing brain-versus body-related age predictions and model feature importance. While we provide feature importance based on model weight scores for transparency, inherent limitations in assessing feature importance limit the generalisability of these rankings (Haufe et al., 2014). For example, weight and gain metrics may bias towards features with higher cardinality, or exclude equally age-dependent features from the model due to multicollinearity (Adler & Painsky, 2022). Additionally, these methods often overlook complex feature interactions and the nonlinear nature of models like XGBoost, leading to potential misinterpretation (Goyal et al., 2020). To enhance model transparency and interpretability, future studies might aim to address these challenges by incorporating permutation feature importance, SHAP values, and partial dependence plots (Altmann et al., 2010; Lundberg & Lee, 2017), alongside external validation on independent datasets as well as pre-modeling strategies like principal component analysis (PCA) to mitigate multicollinearity. Lastly, deep neural network models for age prediction have shown superior performance in recent years (Leonardsen et al., 2022) and should be considered to improve robustness of the methodology.

To summarise, we found that age prediction using bodily health traits performed comparably well to models using brain MRI data alone, and that specific health traits may differentially influence brain and body ageing systems. Our results emphasise the relevance of considering both body and brain measures for a more comprehensive understanding of biological ageing. The current study thus contributes to the dissection of the unique and common variance across body and brain health indicators, which is key towards the aim of parsing inter-individual heterogeneity in the multisystem ageing process. Future research should attempt to better understand the clinical relevance of individual-level discrepancies between different age prediction models in relation to early life exposures, lifestyle factors, genetic architecture, and their relation to risk for cardiovascular disease and age-related neurodegenerative and cognitive disorders.

## 5. Data availability

The UK Biobank resource is open for eligible researchers upon application (http://www.ukbiobank.ac.uk/register-apply/).

## 6. Funding and acknowledgements

The Research Council of Norway (#223273, #324252, #300767, #324499); South-Eastern Norway Regional Health Authority (#2017112; #2019101, #2022080, #2020060); European Union’s Horizon 2020 Research and Innovation Programme (CoMorMent project, Grant #847776; BRAINMINT project, Grant #802998), the German Federal Ministry of Education and Research (01ZX1904A), and the Swiss National Science Foundation (#PZ00P3_193658, #TMPFP3_217174). We performed this work on the *Services for sensitive data* (TSD), University of Oslo, Norway, with resources provided by UNINETT Sigma2 - the National Infrastructure for High-Performance Computing and Data Storage in Norway. We conducted this research using the UK Biobank Resource under Application Number 27412.

## References

Adler, A. I., & Painsky, A. (2022). Feature Importance in Gradient Boosting Trees with Cross-Validation Feature Selection. Entropy, 24(5), Article 5. 10.3390/e24050687

Alfaro, F. J., Gavrieli, A., Saade-Lemus, P., Lioutas, V.-A., Upadhyay, J., & Novak, V. (2018). White matter microstructure and cognitive decline in metabolic syndrome: A review of diffusion tensor imaging. Metabolism: Clinical and Experimental, 78, 52–68. 10.1016/j.metabol.2017.08.009

Altmann, A., Toloşi, L., Sander, O., & Lengauer, T. (2010). Permutation importance: A corrected feature importance measure. Bioinformatics, 26(10), 1340–1347. 10.1093/bioinformatics/btq134

Amundson, D. E., Djurkovic, S., & Matwiyoff, G. N. (2010). The Obesity Paradox. Critical Care Clinics, 26(4), 583–596. 10.1016/j.ccc.2010.06.004

Anatürk, M., Kaufmann, T., Cole, J. H., Suri, S., Griffanti, L., Zsoldos, E., Filippini, N., Singh-Manoux, A., Kivimäki, M., Westlye, L. T., Ebmeier, K. P., & Lange, A.-M. G. de. (2021). Prediction of brain age and cognitive age: Quantifying brain and cognitive maintenance in aging. Human Brain Mapping, 42(6), 1626–1640. 10.1002/hbm.25316

Ba, G., Vt, C., Ma, E., S, K., Hl, W., & Mh, C. (2012). The older the better: Are elderly study participants more non-representative? A cross-sectional analysis of clinical trial and observational study samples. BMJ Open, 2(6). 10.1136/bmjopen-2012-000833

Basser, P. J. (1995). Inferring microstructural features and the physiological state of tissues from diffusion-weighted images. NMR in Biomedicine, 8(7), 333–344. 10.1002/nbm.1940080707

Bates, D., Mächler, M., Bolker, B., & Walker, S. (2015). Fitting Linear Mixed-Effects Models Using lme4. Journal of Statistical Software, 67(1). 10.18637/jss.v067.i01

Beck, D., de Lange, A.-M. G., Alnæs, D., Maximov, I. I., Pedersen, M. L., Leinhard, O. D., Linge, J., Simon, R., Richard, G., Ulrichsen, K. M., Dørum, E. S., Kolskår, K. K., Sanders, A.-M., Winterton, A., Gurholt, T. P., Kaufmann, T., Steen, N. E., Nordvik, J. E., Andreassen, O. A., & Westlye, L. T. (2022). Adipose tissue distribution from body MRI is associated with cross-sectional and longitudinal brain age in adults. NeuroImage: Clinical, 33, 102949. 10.1016/j.nicl.2022.102949

Beck, D., de Lange, A.-M. G., Maximov, I. I., Richard, G., Andreassen, O. A., Nordvik, J. E., & Westlye, L. T. (2021). White matter microstructure across the adult lifespan: A mixed longitudinal and cross-sectional study using advanced diffusion models and brain-age prediction. NeuroImage, 224, 117441. 10.1016/j.neuroimage.2020.117441

Beck, D., de Lange, A.-M. G., Pedersen, M. L., Alnæs, D., Maximov, I. I., Voldsbekk, I., Richard, G., Sanders, A.-M., Ulrichsen, K. M., Dørum, E. S., Kolskår, K. K., Høgestøl, E. A., Steen, N. E., Djurovic, S., Andreassen, O. A., Nordvik, J. E., Kaufmann, T., & Westlye, L. T. (2022). Cardiometabolic risk factors associated with brain age and accelerate brain ageing. Human Brain Mapping, 43(2), 700–720. 10.1002/hbm.25680

Borga, M., Ahlgren, A., Romu, T., Widholm, P., Dahlqvist Leinhard, O., & West, J. (2020). Reproducibility and repeatability of MRI-based body composition analysis. Magnetic Resonance in Medicine, 84(6), 3146–3156. 10.1002/mrm.28360

Borga, M., West, J., Bell, J. D., Harvey, N. C., Romu, T., Heymsfield, S. B., & Dahlqvist Leinhard, O. (2018). Advanced body composition assessment: From body mass index to body composition profiling. Journal of Investigative Medicine: The Official Publication of the American Federation for Clinical Research, 66(5), 1–9. 10.1136/jim-2018-000722

Bosello, O., & Vanzo, A. (2021). Obesity paradox and aging. Eating and Weight Disorders - Studies on Anorexia, Bulimia and Obesity, 26(1), 27–35. 10.1007/s40519-019-00815-4

Brain, J., Greene, L., Tang, E. Y. H., Louise, J., Salter, A., Beach, S., Turnbull, D., Siervo, M., Stephan, B. C. M., & Tully, P. J. (2023). Cardiovascular disease, associated risk factors, and risk of dementia: An umbrella review of meta-analyses. Frontiers in Epidemiology, 3. https://www.frontiersin.org/articles/10.3389/fepid.2023.1095236

Brayne, C., & Moffitt, T. E. (2022). The limitations of large-scale volunteer databases to address inequalities and global challenges in health and aging. Nature Aging, 2(9), Article 9. 10.1038/s43587-022-00277-x

Bürkner, P.-C. (2017). brms: An *R* Package for Bayesian Multilevel Models Using *Stan*. Journal of Statistical Software, 80(1). 10.18637/jss.v080.i01

Bürkner, P.-C. (2018). Advanced Bayesian Multilevel Modeling with the R Package brms. The R Journal, 10(1), 395. 10.32614/RJ-2018-017

Cevenini, E., Invidia, L., Lescai, F., Salvioli, S., Tieri, P., Castellani, G., & Franceschi, C. (2008). Human models of aging and longevity. Expert Opinion on Biological Therapy, 8(9), 1393–1405. 10.1517/14712598.8.9.1393

Chen, T., & Guestrin, C. (2016). XGBoost: A Scalable Tree Boosting System. Proceedings of the 22nd ACM SIGKDD International Conference on Knowledge Discovery and Data Mining, 785–794. 10.1145/2939672.2939785

Cole, & Franke, K. (2017). Predicting Age Using Neuroimaging: Innovative Brain Ageing Biomarkers. Trends in Neurosciences, 40(12), 681–690. 10.1016/j.tins.2017.10.001

Cole, J. H., Marioni, R. E., Harris, S. E., & Deary, I. J. (2019). Brain age and other bodily ‘ages’: Implications for neuropsychiatry. Molecular Psychiatry, 24(2), 266–281. 10.1038/s41380-018-0098-1

Cole, J. H., Poudel, R. P. K., Tsagkrasoulis, D., Caan, M. W. A., Steves, C., Spector, T. D., & Montana, G. (2017). Predicting brain age with deep learning from raw imaging data results in a reliable and heritable biomarker. NeuroImage, 163, 115–124. 10.1016/j.neuroimage.2017.07.059

Cole, J. H., Ritchie, S. J., Bastin, M. E., Hernández, M. C. V., Maniega, S. M., Royle, N., Corley, J., Pattie, A., Harris, S. E., Zhang, Q., Wray, N. R., Redmond, P., Marioni, R. E., Starr, J. M., Cox, S. R., Wardlaw, J. M., Sharp, D. J., & Deary, I. J. (2018). Brain age predicts mortality. Molecular Psychiatry, 23(5), 1385–1392. 10.1038/mp.2017.62

Collins, R. (2007). UK Biobank Protocol.

de Lange, A.-M. G., Anatürk, M., Suri, S., Kaufmann, T., Cole, J. H., Griffanti, L., Zsoldos, E., Jensen, D. E. A., Filippini, N., Singh-Manoux, A., Kivimäki, M., Westlye, L. T., & Ebmeier, K. P. (2020). Multimodal brain-age prediction and cardiovascular risk: The Whitehall II MRI sub-study. NeuroImage, 222, 117292. 10.1016/j.neuroimage.2020.117292

de Lange, A.-M. G., Kaufmann, T., van der Meer, D., Maglanoc, L. A., Alnæs, D., Moberget, T., Douaud, G., Andreassen, O. A., & Westlye, L. T. (2019). Population-based neuroimaging reveals traces of childbirth in the maternal brain. Proceedings of the National Academy of Sciences, 116(44), 22341–22346. 10.1073/pnas.1910666116

Dintica, C. S., Habes, M., Erus, G., Vittinghoff, E., Davatzikos, C., Nasrallah, I. M., Launer, L. J., Sidney, S., & Yaffe, K. (2023). Elevated blood pressure is associated with advanced brain aging in mid-life: A 30-year follow-up of The CARDIA Study. Alzheimer’s & Dementia, 19(3), 924–932. 10.1002/alz.12725

Fieremans, E., Jensen, J. H., & Helpern, J. A. (2011). White matter characterization with diffusional kurtosis imaging. NeuroImage, 58(1), 177–188. 10.1016/j.neuroimage.2011.06.006

Fischl, B., Salat, D. H., Busa, E., Albert, M., Dieterich, M., Haselgrove, C., van der Kouwe, A., Killiany, R., Kennedy, D., Klaveness, S., Montillo, A., Makris, N., Rosen, B., & Dale, A. M. (2002). Whole Brain Segmentation. Neuron, 33(3), 341–355. 10.1016/S0896-6273(02)00569-X

Franke, K., Gaser, C., Manor, B., & Novak, V. (2013). Advanced BrainAGE in older adults with type 2 diabetes mellitus. Frontiers in Aging Neuroscience, 5. 10.3389/fnagi.2013.00090

Franke, K., Ristow, M., & Gaser, C. (2014). Gender-specific impact of personal health parameters on individual brain aging in cognitively unimpaired elderly subjects. Frontiers in Aging Neuroscience, 6. 10.3389/fnagi.2014.00094

Franke, K., Ziegler, G., Klöppel, S., & Gaser, C. (2010). Estimating the age of healthy subjects from T1-weighted MRI scans using kernel methods: Exploring the influence of various parameters. NeuroImage, 50(3), 883–892. 10.1016/j.neuroimage.2010.01.005

George, K. M., Maillard, P., Gilsanz, P., Fletcher, E., Peterson, R. L., Fong, J., Mayeda, E. R., Mungas, D. M., Barnes, L. L., Glymour, M. M., DeCarli, C., & Whitmer, R. A. (2023). Association of Early Adulthood Hypertension and Blood Pressure Change With Late-Life Neuroimaging Biomarkers. JAMA Network Open, 6(4), e236431. 10.1001/jamanetworkopen.2023.6431

Glasser, M. F., Coalson, T. S., Robinson, E. C., Hacker, C. D., Harwell, J., Yacoub, E., Ugurbil, K., Andersson, J., Beckmann, C. F., Jenkinson, M., Smith, S. M., & Van Essen, D. C. (2016). A multi-modal parcellation of human cerebral cortex. Nature, 536(7615), Article 7615. 10.1038/nature18933

Gong, J., Harris, K., Peters, S. A. E., & Woodward, M. (2021). Sex differences in the association between major cardiovascular risk factors in midlife and dementia: A cohort study using data from the UK Biobank. BMC Medicine, 19(1), 110. 10.1186/s12916-021-01980-z

Goyal, K., Dumancic, S., & Blockeel, H. (2020). Feature Interactions in XGBoost (arXiv:2007.05758). arXiv. 10.48550/arXiv.2007.05758

Grady, C. (2012). The cognitive neuroscience of ageing. Nature Reviews Neuroscience, 13(7), 491–505. 10.1038/nrn3256

Haufe, S., Meinecke, F., Görgen, K., Dähne, S., Haynes, J.-D., Blankertz, B., & Bießmann, F. (2014). On the interpretation of weight vectors of linear models in multivariate neuroimaging. NeuroImage, 87, 96–110. 10.1016/j.neuroimage.2013.10.067

Jensen, J. H., Helpern, J. A., Ramani, A., Lu, H., & Kaczynski, K. (2005). Diffusional kurtosis imaging: The quantification of non-gaussian water diffusion by means of magnetic resonance imaging. Magnetic Resonance in Medicine, 53(6), 1432–1440. 10.1002/mrm.20508

Kaden, E., Kelm, N. D., Carson, R. P., Does, M. D., & Alexander, D. C. (2016). Multi-compartment microscopic diffusion imaging. NeuroImage, 139, 346–359. 10.1016/j.neuroimage.2016.06.002

Karlsson, A., Rosander, J., Romu, T., Tallberg, J., Grönqvist, A., Borga, M., & Dahlqvist Leinhard, O. (2015). Automatic and quantitative assessment of regional muscle volume by multi-atlas segmentation using whole-body water-fat MRI. Journal of Magnetic Resonance Imaging: JMRI, 41(6), 1558–1569. 10.1002/jmri.24726

Kaufmann, T., van der Meer, D., Doan, N. T., Schwarz, E., Lund, M. J., Agartz, I., Alnæs, D., Barch, D. M., Baur-Streubel, R., Bertolino, A., Bettella, F., Beyer, M. K., Bøen, E., Borgwardt, S., Brandt, C. L., Buitelaar, J., Celius, E. G., Cervenka, S., Conzelmann, A., … Westlye, L. T. (2019). Common brain disorders are associated with heritable patterns of apparent aging of the brain. Nature Neuroscience, 22(10), Article 10. 10.1038/s41593-019-0471-7

Kivimäki, M., Luukkonen, R., Batty, G. D., Ferrie, J. E., Pentti, J., Nyberg, S. T., Shipley, M. J., Alfredsson, L., Fransson, E. I., Goldberg, M., Knutsson, A., Koskenvuo, M., Kuosma, E., Nordin, M., Suominen, S. B., Theorell, T., Vuoksimaa, E., Westerholm, P., Westerlund, H., … Jokela, M. (2018). Body mass index and risk of dementia: Analysis of individual-level data from 1.3 million individuals. Alzheimer’s & Dementia, 14(5), 601–609. 10.1016/j.jalz.2017.09.016

Kolenic, M., Franke, K., Hlinka, J., Matejka, M., Capkova, J., Pausova, Z., Uher, R., Alda, M., Spaniel, F., & Hajek, T. (2018). Obesity, dyslipidemia and brain age in first-episode psychosis. Journal of Psychiatric Research, 99, 151–158. 10.1016/j.jpsychires.2018.02.012

Kuo, C.-Y., Tai, T.-M., Lee, P.-L., Tseng, C.-W., Chen, C.-Y., Chen, L.-K., Lee, C.-K., Chou, K.-H., See, S., & Lin, C.-P. (2021). Improving Individual Brain Age Prediction Using an Ensemble Deep Learning Framework. Frontiers in Psychiatry, 12, 308. 10.3389/fpsyt.2021.626677

Leonardsen, E. H., Peng, H., Kaufmann, T., Agartz, I., Andreassen, O. A., Celius, E. G., Espeseth, T., Harbo, H. F., Høgestøl, E. A., Lange, A.-M. de, Marquand, A. F., Vidal-Piñeiro, D., Roe, J. M., Selbæk, G., Sørensen, Ø., Smith, S. M., Westlye, L. T., Wolfers, T., & Wang, Y. (2022). Deep neural networks learn general and clinically relevant representations of the ageing brain. NeuroImage, 256, 119210. 10.1016/j.neuroimage.2022.119210

Linge, J., Borga, M., West, J., Tuthill, T., Miller, M. R., Dumitriu, A., Thomas, E. L., Romu, T., Tunón, P., Bell, J. D., & Dahlqvist Leinhard, O. (2018). Body Composition Profiling in the UK Biobank Imaging Study: Body Composition Profiling in UK Biobank. Obesity, 26(11), 1785–1795. 10.1002/oby.22210

López-Otín, C., Blasco, M. A., Partridge, L., Serrano, M., & Kroemer, G. (2013). The Hallmarks of Aging. Cell, 153(6), 1194–1217. 10.1016/j.cell.2013.05.039

Lundberg, S., & Lee, S.-I. (2017). A Unified Approach to Interpreting Model Predictions (arXiv:1705.07874). arXiv. 10.48550/arXiv.1705.07874

Massy-Westropp, N. M., Gill, T. K., Taylor, A. W., Bohannon, R. W., & Hill, C. L. (2011). Hand Grip Strength: Age and gender stratified normative data in a population-based study. BMC Research Notes, 4, 127. 10.1186/1756-0500-4-127

Masters, R. K., Powers, D. A., & Link, B. G. (2013). Obesity and US Mortality Risk Over the Adult Life Course. American Journal of Epidemiology, 177(5), 431–442. 10.1093/aje/kws325

Maximov, I. I., Alnæs, D., & Westlye, L. T. (2019). Towards an optimised processing pipeline for diffusion magnetic resonance imaging data: Effects of artefact corrections on diffusion metrics and their age associations in UK Biobank. Human Brain Mapping, 40(14), 4146–4162. 10.1002/hbm.24691

Maximov, I. I., van der Meer, D., de Lange, A.-M. G., Kaufmann, T., Shadrin, A., Frei, O., Wolfers, T., & Westlye, L. T. (2021). Fast qualitY conTrol meThod foR derIved diffUsion Metrics (YTTRIUM) in big data analysis: U.K. Biobank 18,608 example. Human Brain Mapping, 42(10), 3141–3155. 10.1002/hbm.25424

Maximov, I. I., van der Meer, D., de Lange, A.-M., Kaufmann, T., Shadrin, A., Frei, O., Wolfers, T., & Westlye, L. T. (2020). Fast qualit **Y** con **T** rol me **T** hod fo **R** der **I** ved diff **U** sion **M** etrics ( **YTTRIUM** ) in big data analysis: UK Biobank 18608 example [Preprint]. Neuroscience. 10.1101/2020.02.17.952697

Mielke, M. M., Bandaru, V. V. R., Haughey, N. J., Rabins, P. V., Lyketsos, C. G., & Carlson, M. C. (2010). Serum sphingomyelins and ceramides are early predictors of memory impairment. Neurobiology of Aging, 31(1), 17–24. 10.1016/j.neurobiolaging.2008.03.011

Miller, K. L., Alfaro-Almagro, F., Bangerter, N. K., Thomas, D. L., Yacoub, E., Xu, J., Bartsch, A. J., Jbabdi, S., Sotiropoulos, S. N., Andersson, J. L. R., Griffanti, L., Douaud, G., Okell, T. W., Weale, P., Dragonu, I., Garratt, S., Hudson, S., Collins, R., Jenkinson, M., … Smith, S. M. (2016). Multimodal population brain imaging in the UK Biobank prospective epidemiological study. Nature Neuroscience, 19(11), 1523– 1536. 10.1038/nn.4393

Mori, S., Wakana, Nagae-Poetscher, & van Zijl, P. C. M. (2006). MRI Atlas of Human White Matter. AJNR: American Journal of Neuroradiology, 27(6), 1384–1385.

Nyberg, L. (2017). Neuroimaging in aging: Brain maintenance. F1000Research, 6, 1215. 10.12688/f1000research.11419.1

Pedregosa, F., Varoquaux, G., Gramfort, A., Michel, V., Thirion, B., Grisel, O., Blondel, M., Prettenhofer, P., Weiss, R., Dubourg, V., Vanderplas, J., Passos, A., Cournapeau, D., Brucher, M., Perrot, M., & Duchesnay, É. (2011). Scikit-learn: Machine Learning in Python. The Journal of Machine Learning Research, 12(null), 2825–2830.

Rodgers, J. L., Jones, J., Bolleddu, S. I., Vanthenapalli, S., Rodgers, L. E., Shah, K., Karia, K., & Panguluri, S. K. (2019). Cardiovascular Risks Associated with Gender and Aging. Journal of Cardiovascular Development and Disease, 6(2), Article 2. 10.3390/jcdd6020019

Ronan, L., Alexander-Bloch, A. F., Wagstyl, K., Farooqi, S., Brayne, C., Tyler, L. K., & Fletcher, P. C. (2016). Obesity associated with increased brain age from midlife. Neurobiology of Aging, 47, 63–70. 10.1016/j.neurobiolaging.2016.07.010

Sanders, A.-M., Richard, G., Kolskår, K., Ulrichsen, K. M., Kaufmann, T., Alnæs, D., Beck, D., Dørum, E. S., de Lange, A.-M. G., Egil Nordvik, J., & Westlye, L. T. (2021). Linking objective measures of physical activity and capability with brain structure in healthy community dwelling older adults. NeuroImage: Clinical, 31, 102767. 10.1016/j.nicl.2021.102767

Schindler, L. S., Subramaniapillai, S., Barth, C., van der Meer, D., Pedersen, M. L., Kaufmann, T., Maximov, I. I., Linge, J., Leinhard, O. D., Beck, D., Gurholt, T. P., Voldsbekk, I., Suri, S., Ebmeier, K. P., Draganski, B., Andreassen, O. A., Westlye, L. T., & de Lange, A.-M. G. (2022). Associations between abdominal adipose tissue, reproductive span, and brain characteristics in post-menopausal women. NeuroImage: Clinical, 36, 103239. 10.1016/j.nicl.2022.103239

Sebastiani, P., Thyagarajan, B., Sun, F., Schupf, N., Newman, A. B., Montano, M., & Perls, T. T. (2017). Biomarker signatures of aging. Aging Cell, 16(2), 329–338. 10.1111/acel.12557

Smith, S. M., Jenkinson, M., Johansen-Berg, H., Rueckert, D., Nichols, T. E., Mackay, C. E., Watkins, K. E., Ciccarelli, O., Cader, M. Z., Matthews, P. M., & Behrens, T. E. J. (2006). Tract-based spatial statistics: Voxelwise analysis of multi-subject diffusion data. NeuroImage, 31(4), 1487–1505. 10.1016/j.neuroimage.2006.02.024

Stern, Y. (2009). Cognitive reserve. Neuropsychologia, 47(10), 2015–2028. 10.1016/j.neuropsychologia.2009.03.004

Stern, Y. (2012). Cognitive reserve in ageing and Alzheimer’s disease. The Lancet Neurology, 11(11), 1006–1012. 10.1016/S1474-4422(12)70191-6

Subramaniapillai, S., Suri, S., Barth, C., Maximov, I. I., Voldsbekk, I., van der Meer, D., Gurholt, T. P., Beck, D., Draganski, B., Andreassen, O. A., Ebmeier, K. P., Westlye, L. T., & de Lange, A.-M. G. (2022). Sex- and age-specific associations between cardiometabolic risk and white matter brain age in the UK Biobank cohort. Human Brain Mapping, n/a(n/a). 10.1002/hbm.25882

Tian, Y. E., Cropley, V., Maier, A. B., Lautenschlager, N. T., Breakspear, M., & Zalesky, A. (2023). Heterogeneous aging across multiple organ systems and prediction of chronic disease and mortality. Nature Medicine, 1–11. 10.1038/s41591-023-02296-6

Tutor, A. W., Lavie, C. J., Kachur, S., Milani, R. V., & Ventura, H. O. (2023). Updates on obesity and the obesity paradox in cardiovascular diseases. Progress in Cardiovascular Diseases, 78, 2–10. 10.1016/j.pcad.2022.11.013

Tyrrell, J., Zheng, J., Beaumont, R., Hinton, K., Richardson, T. G., Wood, A. R., Davey Smith, G., Frayling, T. M., & Tilling, K. (2021). Genetic predictors of participation in optional components of UK Biobank. Nature Communications, 12(1), Article 1. 10.1038/s41467-021-21073-y

Voldsbekk, I., Barth, C., Maximov, I. I., Kaufmann, T., Beck, D., Richard, G., Moberget, T., Westlye, L. T., & de Lange, A.-M. G. (2021). A history of previous childbirths is linked to women’s white matter brain age in midlife and older age. Human Brain Mapping, 42(13), 4372–4386. 10.1002/hbm.25553

Wagenmakers, E.-J., Lodewyckx, T., Kuriyal, H., & Grasman, R. (2010). Bayesian hypothesis testing for psychologists: A tutorial on the Savage–Dickey method. Cognitive Psychology, 60(3), 158–189. 10.1016/j.cogpsych.2009.12.001

West, J., Romu, T., Thorell, S., Lindblom, H., Berin, E., Holm, A.-C. S., Åstrand, L. L., Karlsson, A., Borga, M., Hammar, M., & Leinhard, O. D. (2018). Precision of MRI- based body composition measurements of postmenopausal women. PloS One, 13(2), e0192495. 10.1371/journal.pone.0192495

Wong, M. W., Braidy, N., Poljak, A., Pickford, R., Thambisetty, M., & Sachdev, P. S. (2017). Dysregulation of lipids in Alzheimer’s disease and their role as potential biomarkers. Alzheimer’s & Dementia, 13(7), 810–827. 10.1016/j.jalz.2017.01.008

