## Supplementary Material for "Dissecting unique and common variance across body and brain health indicators using age prediction"

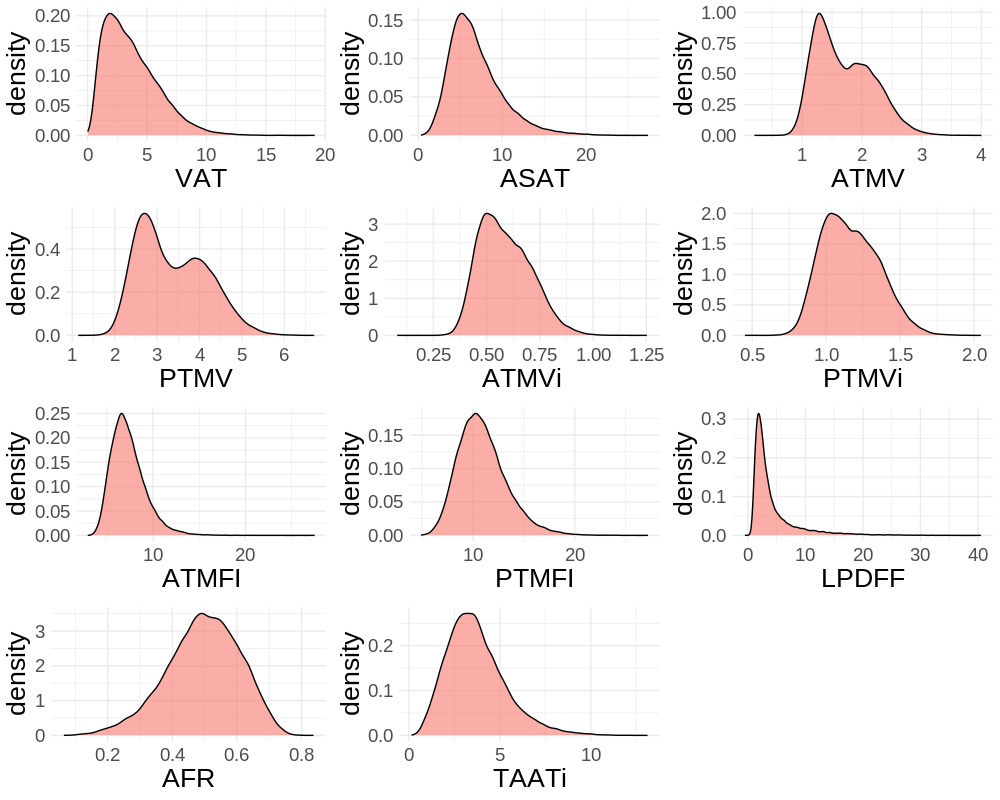

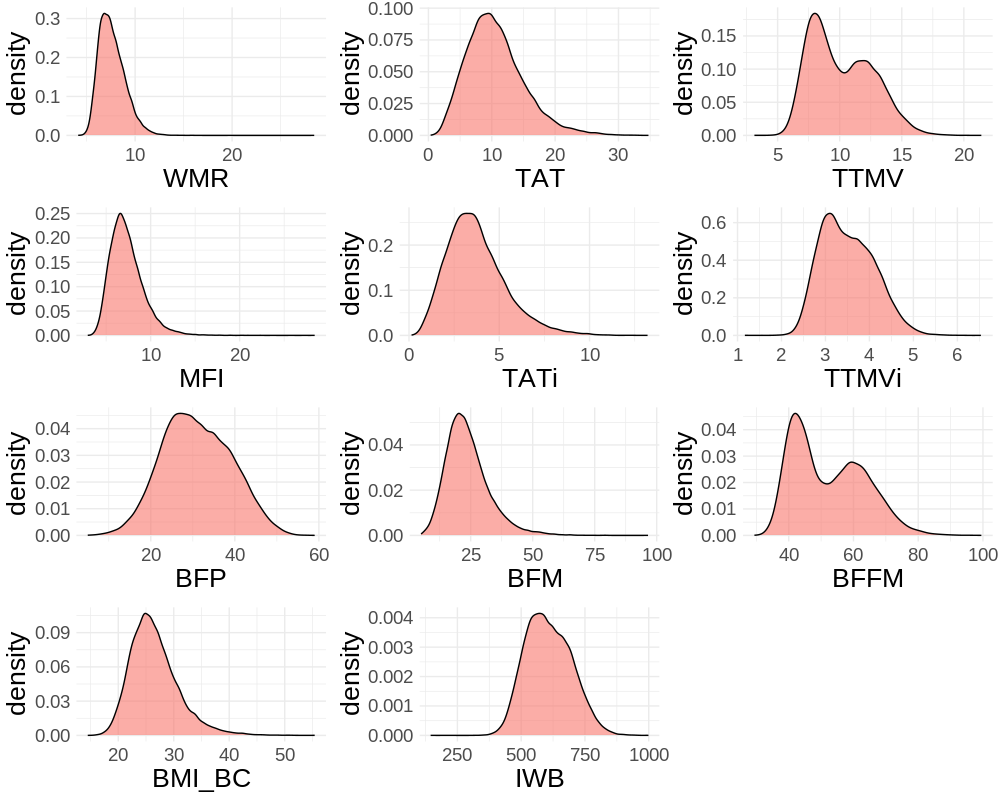

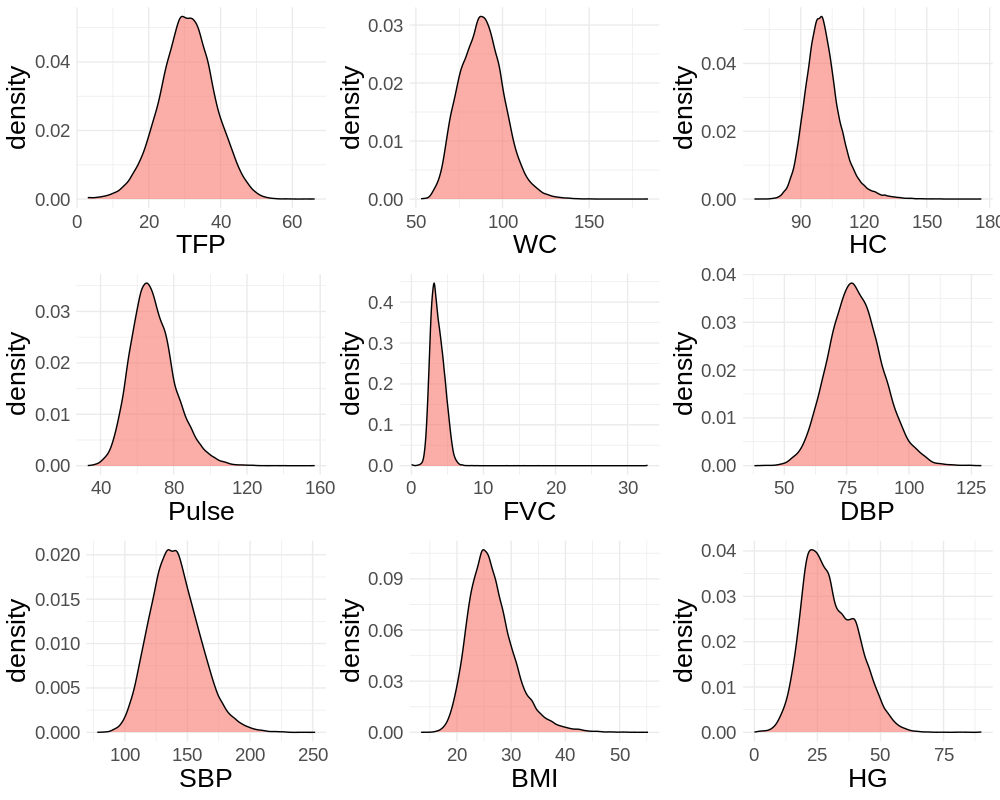

**SI Figure 1**. Distribution of health traits *before* quality checking procedure.

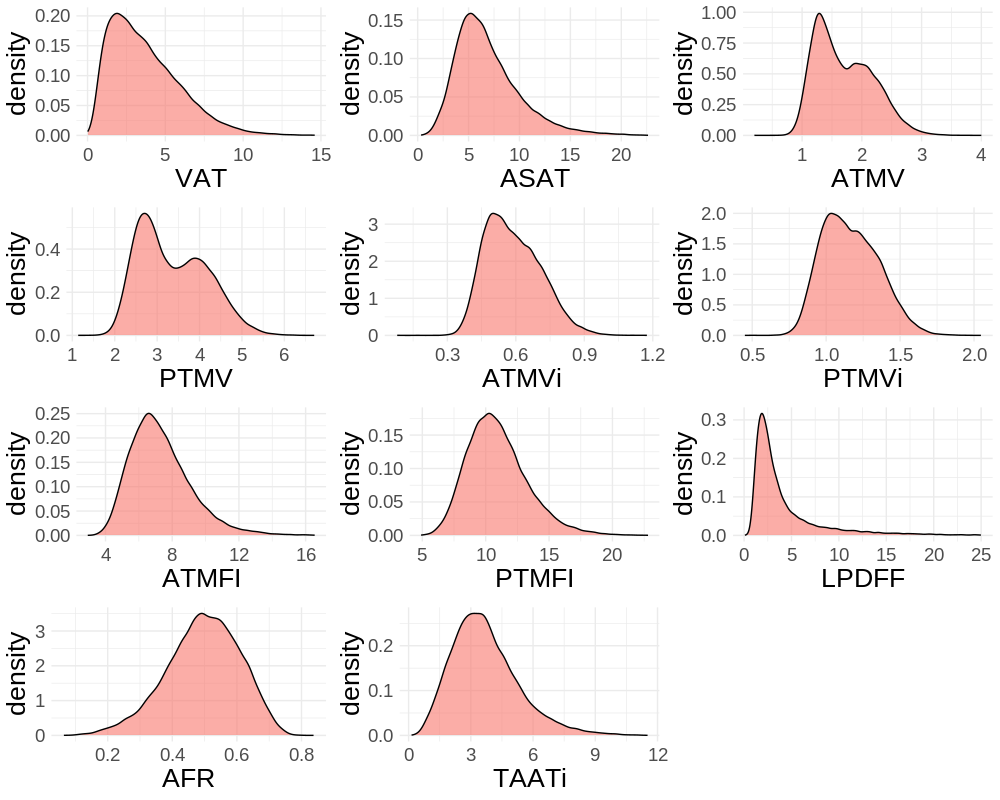

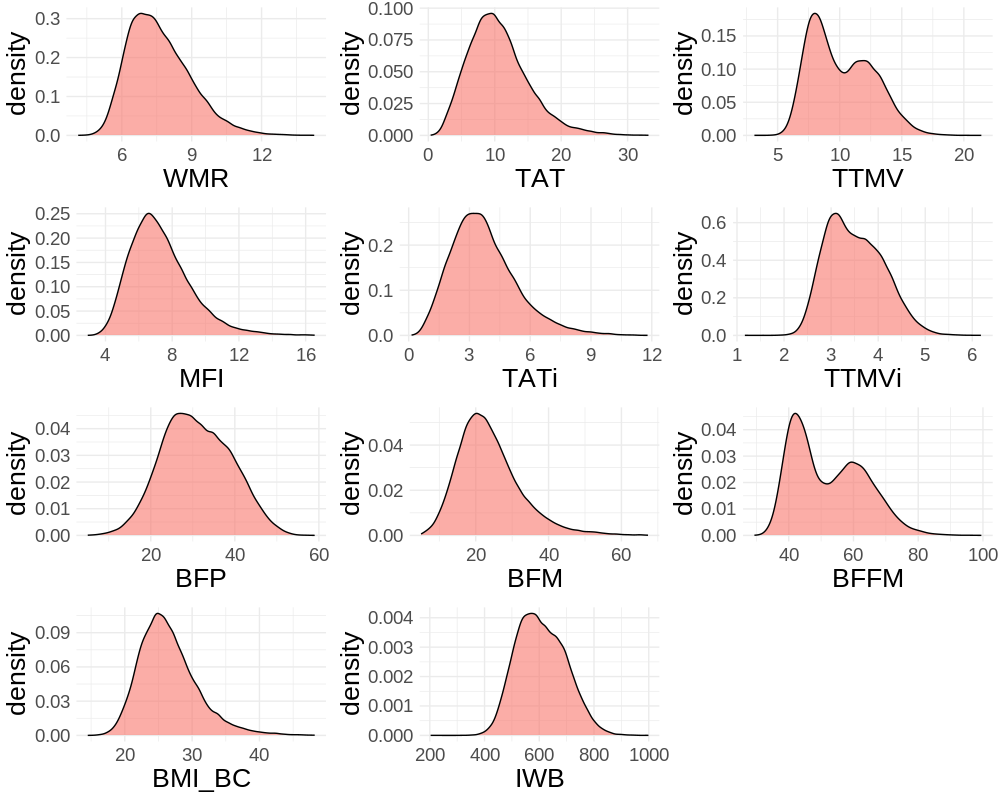

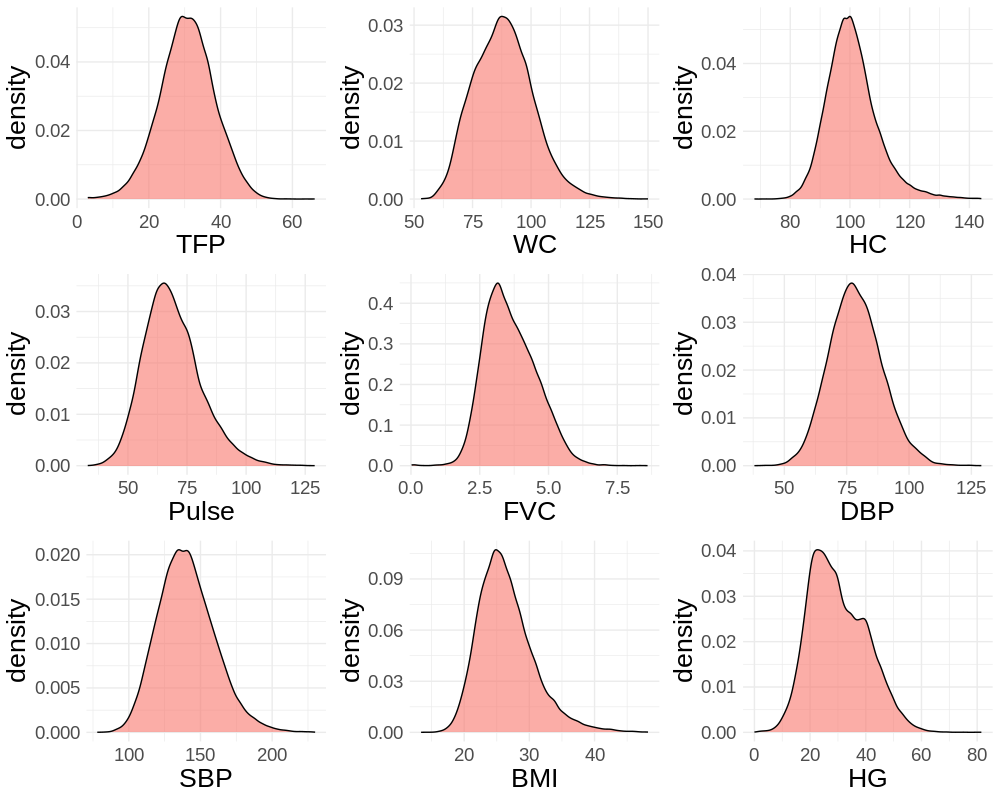

**SI Figure 2**. Distribution of health traits *after* quality checking procedure.

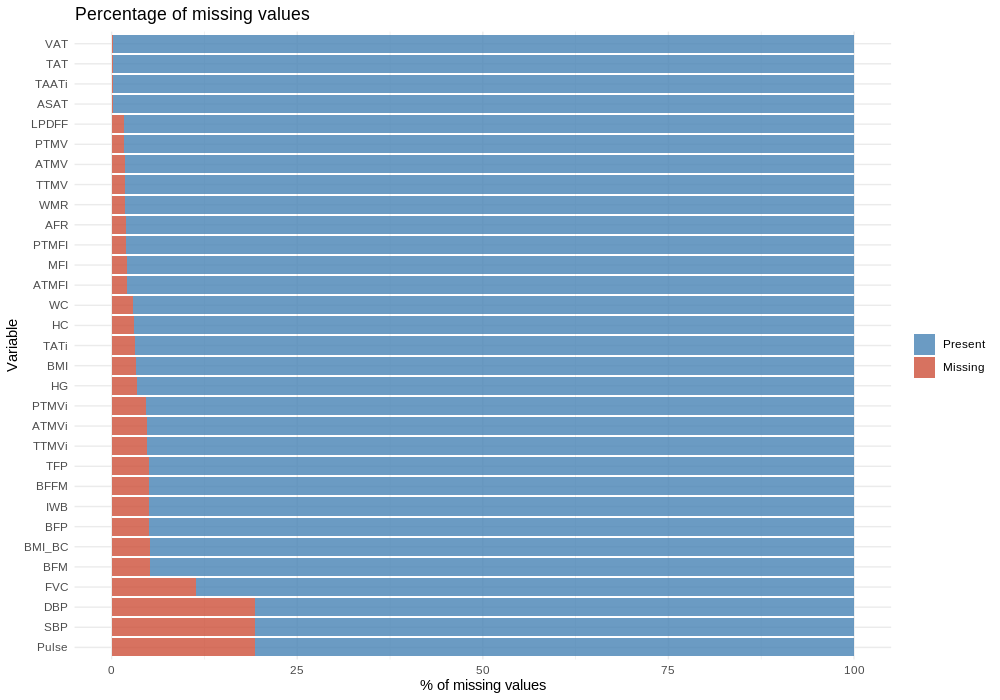

**SI Figure 3.** Showing the missingness of the health trait data post-cleaning. Missing values consist of missing data, previous outlier values, and responses representing refusal to respond to questions.

**SI Section 1**. Description of each bodily health trait included in the study.

We extracted a selection of body composition and health traits (See also UKB online documentation (<http://biobank.ctsu.ox.ac.uk/showcase/>)). Specifically, we extracted the following measures of adipose and muscle tissue from body magnetic resonance imaging (MRI) (highlighted in Table 1 of the main manuscript): liver fat (LPDFF), describing the proton density fat fraction in the liver, measured as a percentage (%); visceral adipose tissue (VAT), describing the intra-abdominal fat surrounding the organs, measured in litres (L); abdominal subcutaneous adipose tissue (ASAT), defined as the adipose tissue underneath the skin, measured in L; total (TTMV), anterior (ATMV), and posterior (PTMV) thigh fat-free muscle volume, describing muscle volume in the thighs, measured in L; total (MFI), anterior (ATMFI), and posterior (PTMFI) muscle fat infiltration, describing the intra-muscular fat in the thighs, measured as a %; total abdominal tissue volume (TAT), which is the total VAT and ASAT, measured in L; abdominal fat ratio (AFR), which is the total abdominal tissue volume (TAT) divided by itself (TAT) and thigh muscle volume (TTMV), measured as a %; weight-to-muscle ratio (WMR), which is body weight divided by thigh muscle volume, measured as a kg/L. For VAT, ASAT, ATMV, PTMV, TTMV and TAT, we computed index measures by dividing these measures by the squared standing height in meters (e.g., VATi is VAT/m^2^). Additionally, a TAATi measure consisting of the total *abdominal* adipose tissue index was calculated. This is done since weight, adipose tissue, and lean tissue compartments scale to approximate height squared.

For body composition by bioimpedance measures, we extracted the following measures: body fat percentage (BFP), describing body fat percentage in a range between 1-75% in 0.1% increments; whole body fat mass (BFM), describing total fat mass in kilograms (kg); whole body fat-free mass (BFFM), describing fat-free mass in kg; body-mass index body composition (BMI-BC), describing the BMI measurement taken at body impedance (kg/m^2^); impedance of whole body (IWB), describing electrical resistance in ohm (Range 150 - 1200ohms in 1ohm increments); trunk fat percentage (TFP), describing fat in range 1-75% and 0.1% increments for the area that contains the chest, abdomen, pelvis, and back (also known as torso).

For cardiometabolic and anthropometric measure from physical examinations, we extracted the following measures: waist circumference (WC), describing the circumference (in centimetres [cm]) of the midpoint area between the top of the hipbone and lowest rib bone; Hip circumference (HC), describing the circumference (cm) of the midpoint area of the hipbone; BMI, calculated as weight (kg) divided by height^2^ (m^2^); hand grip strength (HG), describing the force exerted by the hand muscles when gripping a hand dynamometer, measured in kg; pulse, describing rhythmic expansion and contraction of the arteries caused by the heartbeat and measured as beats per minute (bpm); systolic blood pressure (SBP), describing the average of two automated measures of systolic (highest level of pressure exerted on the walls of the arteries when the heart contracts and pumps blood) BP by means of the Omron device, measured in millimetres (mm) of mercury (mmHg); diastolic blood pressure (DBP), describing the average of two automated measures taken moments apart, measured in mmHg. DBP refers to the lowest level of pressure exerted on the walls of the arteries when the heart is at rest between contractions; and lastly, forced vital capacity (FVC), describing the maximum amount of air the individual can forcefully exhale (spirometry blow) after taking a deep breath, measured in litres (L).

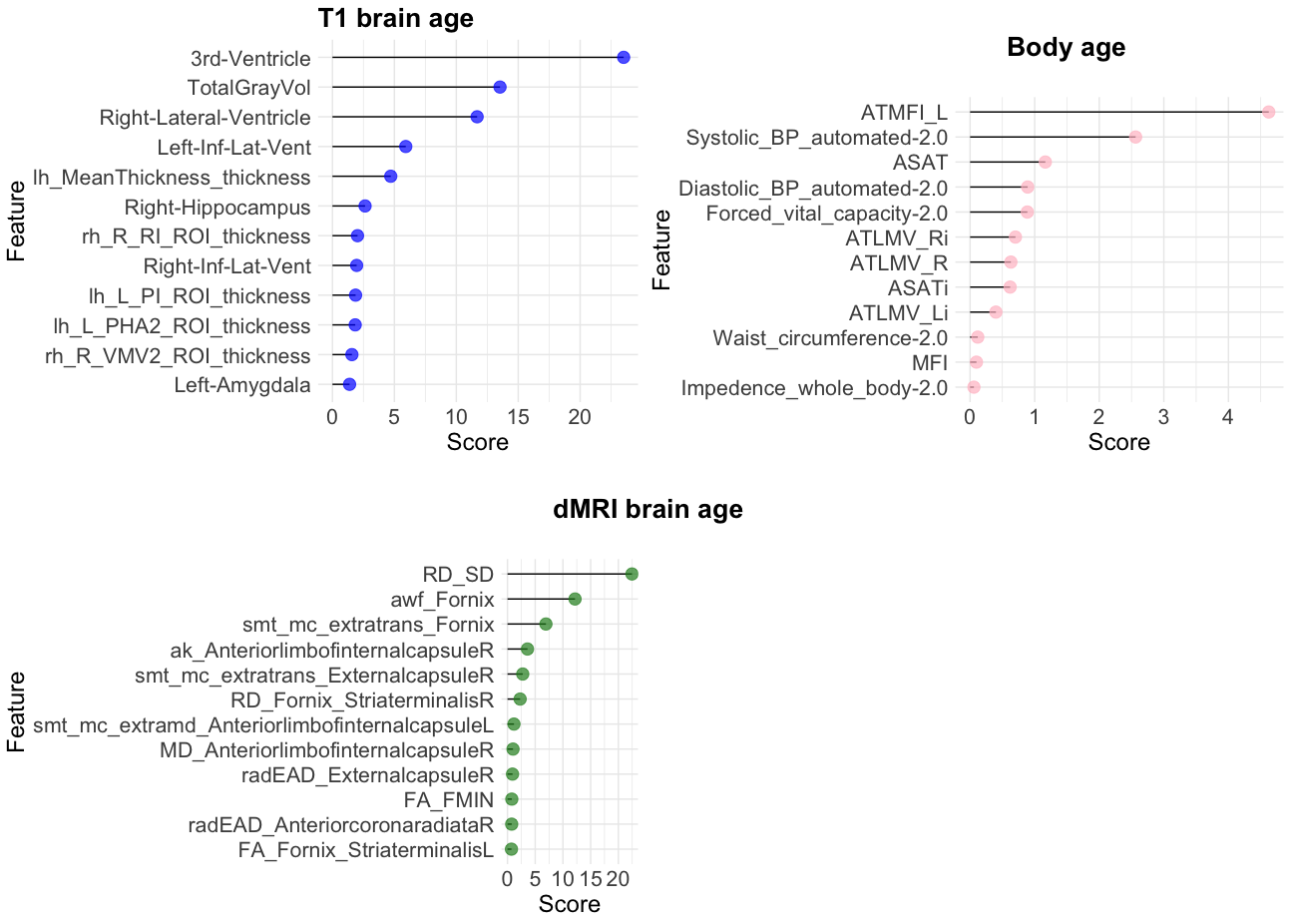
**SI Figure 4**. **Feature importance.** Showing the weight contribution of the twelve top-ranking (scaled) variables of each age prediction model, which indicate the relative contribution of the corresponding feature to the prediction model, calculated based on each feature’s contribution for each tree in the model.

**SI Table 1. Model features**. Showing each of the variables included in the brain age prediction models for T1, dMRI, and body age models as titled in the UK Biobank.

| Model | Variable name list |
| --- | --- |
| T1 | lh_L_V1_ROI_thickness, lh_L_MST_ROI_thickness, lh_L_V6_ROI_thickness, lh_L_V2_ROI_thickness, lh_L_V3_ROI_thickness, lh_L_V4_ROI_thickness, lh_L_V8_ROI_thickness, lh_L_4_ROI_thickness, lh_L_3b_ROI_thickness, lh_L_FEF_ROI_thickness, lh_L_PEF_ROI_thickness, lh_L_55b_ROI_thickness, lh_L_V3A_ROI_thickness, lh_L_RSC_ROI_thickness, lh_L_POS2_ROI_thickness, lh_L_V7_ROI_thickness, lh_L_IPS1_ROI_thickness, lh_L_FFC_ROI_thickness, lh_L_V3B_ROI_thickness, lh_L_LO1_ROI_thickness, lh_L_LO2_ROI_thickness, lh_L_PIT_ROI_thickness, lh_L_MT_ROI_thickness, lh_L_A1_ROI_thickness, lh_L_PSL_ROI_thickness, lh_L_SFL_ROI_thickness, lh_L_PCV_ROI_thickness, lh_L_STV_ROI_thickness, lh_L_7Pm_ROI_thickness, lh_L_7m_ROI_thickness, lh_L_POS1_ROI_thickness, lh_L_23d_ROI_thickness, lh_L_v23ab_ROI_thickness, lh_L_d23ab_ROI_thickness, lh_L_31pv_ROI_thickness, lh_L_5m_ROI_thickness, lh_L_5mv_ROI_thickness, lh_L_23c_ROI_thickness, lh_L_5L_ROI_thickness, lh_L_24dd_ROI_thickness, lh_L_24dv_ROI_thickness, lh_L_7AL_ROI_thickness, lh_L_SCEF_ROI_thickness, lh_L_6ma_ROI_thickness, lh_L_7Am_ROI_thickness, lh_L_7PL_ROI_thickness, lh_L_7PC_ROI_thickness, lh_L_LIPv_ROI_thickness, lh_L_VIP_ROI_thickness, lh_L_MIP_ROI_thickness, lh_L_1_ROI_thickness, lh_L_2_ROI_thickness, lh_L_3a_ROI_thickness, lh_L_6d_ROI_thickness, lh_L_6mp_ROI_thickness, lh_L_6v_ROI_thickness, lh_L_p24pr_ROI_thickness, lh_L_33pr_ROI_thickness, lh_L_a24pr_ROI_thickness, lh_L_p32pr_ROI_thickness, lh_L_a24_ROI_thickness, lh_L_d32_ROI_thickness, lh_L_8BM_ROI_thickness, lh_L_p32_ROI_thickness, lh_L_10r_ROI_thickness, lh_L_47m_ROI_thickness, lh_L_8Av_ROI_thickness, lh_L_8Ad_ROI_thickness, lh_L_9m_ROI_thickness, lh_L_8BL_ROI_thickness, lh_L_9p_ROI_thickness, lh_L_10d_ROI_thickness, lh_L_8C_ROI_thickness, lh_L_44_ROI_thickness, lh_L_45_ROI_thickness, lh_L_47l_ROI_thickness, lh_L_a47r_ROI_thickness, lh_L_6r_ROI_thickness, lh_L_IFJa_ROI_thickness, lh_L_IFJp_ROI_thickness, lh_L_IFSp_ROI_thickness, lh_L_IFSa_ROI_thickness, lh_L_p9-46v_ROI_thickness, lh_L_46_ROI_thickness, lh_L_a9-46v_ROI_thickness, lh_L_9-46d_ROI_thickness, lh_L_9a_ROI_thickness, lh_L_10v_ROI_thickness, lh_L_a10p_ROI_thickness, lh_L_10pp_ROI_thickness, lh_L_11l_ROI_thickness, lh_L_13l_ROI_thickness, lh_L_OFC_ROI_thickness, lh_L_47s_ROI_thickness, lh_L_LIPd_ROI_thickness, lh_L_6a_ROI_thickness, lh_L_i6-8_ROI_thickness, lh_L_s6-8_ROI_thickness, lh_L_43_ROI_thickness, lh_L_OP4_ROI_thickness, lh_L_OP1_ROI_thickness, lh_L_OP2-3_ROI_thickness, lh_L_52_ROI_thickness, lh_L_RI_ROI_thickness, lh_L_PFcm_ROI_thickness, lh_L_PoI2_ROI_thickness, lh_L_TA2_ROI_thickness, lh_L_FOP4_ROI_thickness, lh_L_MI_ROI_thickness, lh_L_Pir_ROI_thickness, lh_L_AVI_ROI_thickness, lh_L_AAIC_ROI_thickness, lh_L_FOP1_ROI_thickness, lh_L_FOP3_ROI_thickness, lh_L_FOP2_ROI_thickness, lh_L_PFt_ROI_thickness, lh_L_AIP_ROI_thickness, lh_L_EC_ROI_thickness, lh_L_PreS_ROI_thickness, lh_L_H_ROI_thickness, lh_L_ProS_ROI_thickness, lh_L_PeEc_ROI_thickness, lh_L_STGa_ROI_thickness, lh_L_PBelt_ROI_thickness, lh_L_A5_ROI_thickness, lh_L_PHA1_ROI_thickness, lh_L_PHA3_ROI_thickness, lh_L_STSda_ROI_thickness, lh_L_STSdp_ROI_thickness, lh_L_STSvp_ROI_thickness, lh_L_TGd_ROI_thickness, lh_L_TE1a_ROI_thickness, lh_L_TE1p_ROI_thickness, lh_L_TE2a_ROI_thickness, lh_L_TF_ROI_thickness, lh_L_TE2p_ROI_thickness, lh_L_PHT_ROI_thickness, lh_L_PH_ROI_thickness, lh_L_TPOJ1_ROI_thickness, lh_L_TPOJ2_ROI_thickness, lh_L_TPOJ3_ROI_thickness, lh_L_DVT_ROI_thickness, lh_L_PGp_ROI_thickness, lh_L_IP2_ROI_thickness, lh_L_IP1_ROI_thickness, lh_L_IP0_ROI_thickness, lh_L_PFop_ROI_thickness, lh_L_PF_ROI_thickness, lh_L_PFm_ROI_thickness, lh_L_PGi_ROI_thickness, lh_L_PGs_ROI_thickness, lh_L_V6A_ROI_thickness, lh_L_VMV1_ROI_thickness, lh_L_VMV3_ROI_thickness, lh_L_PHA2_ROI_thickness, lh_L_V4t_ROI_thickness, lh_L_FST_ROI_thickness, lh_L_V3CD_ROI_thickness, lh_L_LO3_ROI_thickness, lh_L_VMV2_ROI_thickness, lh_L_31pd_ROI_thickness, lh_L_31a_ROI_thickness, lh_L_VVC_ROI_thickness, lh_L_25_ROI_thickness, lh_L_s32_ROI_thickness, lh_L_pOFC_ROI_thickness, lh_L_PoI1_ROI_thickness, lh_L_Ig_ROI_thickness, lh_L_FOP5_ROI_thickness, lh_L_p10p_ROI_thickness, lh_L_p47r_ROI_thickness, lh_L_TGv_ROI_thickness, lh_L_MBelt_ROI_thickness, lh_L_LBelt_ROI_thickness, lh_L_A4_ROI_thickness, lh_L_STSva_ROI_thickness, lh_L_TE1m_ROI_thickness, lh_L_PI_ROI_thickness, lh_L_a32pr_ROI_thickness, lh_L_p24_ROI_thickness, lh_MeanThickness_thickness, rh_R_V1_ROI_thickness, rh_R_MST_ROI_thickness, rh_R_V6_ROI_thickness, rh_R_V2_ROI_thickness, rh_R_V3_ROI_thickness, rh_R_V4_ROI_thickness, rh_R_V8_ROI_thickness, rh_R_4_ROI_thickness, rh_R_3b_ROI_thickness, rh_R_FEF_ROI_thickness, rh_R_PEF_ROI_thickness, rh_R_55b_ROI_thickness, rh_R_V3A_ROI_thickness, rh_R_RSC_ROI_thickness, rh_R_POS2_ROI_thickness, rh_R_V7_ROI_thickness, rh_R_IPS1_ROI_thickness, rh_R_FFC_ROI_thickness, rh_R_V3B_ROI_thickness, rh_R_LO1_ROI_thickness, rh_R_LO2_ROI_thickness, rh_R_PIT_ROI_thickness, rh_R_MT_ROI_thickness, rh_R_A1_ROI_thickness, rh_R_PSL_ROI_thickness, rh_R_SFL_ROI_thickness, rh_R_PCV_ROI_thickness, rh_R_STV_ROI_thickness, rh_R_7Pm_ROI_thickness, rh_R_7m_ROI_thickness, rh_R_POS1_ROI_thickness, rh_R_23d_ROI_thickness, rh_R_v23ab_ROI_thickness, rh_R_d23ab_ROI_thickness, rh_R_31pv_ROI_thickness, rh_R_5m_ROI_thickness, rh_R_5mv_ROI_thickness, rh_R_23c_ROI_thickness, rh_R_5L_ROI_thickness, rh_R_24dd_ROI_thickness, rh_R_24dv_ROI_thickness, rh_R_7AL_ROI_thickness, rh_R_SCEF_ROI_thickness, rh_R_6ma_ROI_thickness, rh_R_7Am_ROI_thickness, rh_R_7PL_ROI_thickness, rh_R_7PC_ROI_thickness, rh_R_LIPv_ROI_thickness, rh_R_VIP_ROI_thickness, rh_R_MIP_ROI_thickness, rh_R_1_ROI_thickness, rh_R_2_ROI_thickness, rh_R_3a_ROI_thickness, rh_R_6d_ROI_thickness, rh_R_6mp_ROI_thickness, rh_R_6v_ROI_thickness, rh_R_p24pr_ROI_thickness, rh_R_33pr_ROI_thickness, rh_R_a24pr_ROI_thickness, rh_R_p32pr_ROI_thickness, rh_R_a24_ROI_thickness, rh_R_d32_ROI_thickness, rh_R_8BM_ROI_thickness, rh_R_p32_ROI_thickness, rh_R_10r_ROI_thickness, rh_R_47m_ROI_thickness, rh_R_8Av_ROI_thickness, rh_R_8Ad_ROI_thickness, rh_R_9m_ROI_thickness, rh_R_8BL_ROI_thickness, rh_R_9p_ROI_thickness, rh_R_10d_ROI_thickness, rh_R_8C_ROI_thickness, rh_R_44_ROI_thickness, rh_R_45_ROI_thickness, rh_R_47l_ROI_thickness, rh_R_a47r_ROI_thickness, rh_R_6r_ROI_thickness, rh_R_IFJa_ROI_thickness, rh_R_IFJp_ROI_thickness, rh_R_IFSp_ROI_thickness, rh_R_IFSa_ROI_thickness, rh_R_p9-46v_ROI_thickness, rh_R_46_ROI_thickness, rh_R_a9-46v_ROI_thickness, rh_R_9-46d_ROI_thickness, rh_R_9a_ROI_thickness, rh_R_10v_ROI_thickness, rh_R_a10p_ROI_thickness, rh_R_10pp_ROI_thickness, rh_R_11l_ROI_thickness, rh_R_13l_ROI_thickness, rh_R_OFC_ROI_thickness, rh_R_47s_ROI_thickness, rh_R_LIPd_ROI_thickness, rh_R_6a_ROI_thickness, rh_R_i6-8_ROI_thickness, rh_R_s6-8_ROI_thickness, rh_R_43_ROI_thickness, rh_R_OP4_ROI_thickness, rh_R_OP1_ROI_thickness, rh_R_OP2-3_ROI_thickness, rh_R_52_ROI_thickness, rh_R_RI_ROI_thickness, rh_R_PFcm_ROI_thickness, rh_R_PoI2_ROI_thickness, rh_R_TA2_ROI_thickness, rh_R_FOP4_ROI_thickness, rh_R_MI_ROI_thickness, rh_R_Pir_ROI_thickness, rh_R_AVI_ROI_thickness, rh_R_AAIC_ROI_thickness, rh_R_FOP1_ROI_thickness, rh_R_FOP3_ROI_thickness, rh_R_FOP2_ROI_thickness, rh_R_PFt_ROI_thickness, rh_R_AIP_ROI_thickness, rh_R_EC_ROI_thickness, rh_R_PreS_ROI_thickness, rh_R_H_ROI_thickness, rh_R_ProS_ROI_thickness, rh_R_PeEc_ROI_thickness, rh_R_STGa_ROI_thickness, rh_R_PBelt_ROI_thickness, rh_R_A5_ROI_thickness, rh_R_PHA1_ROI_thickness, rh_R_PHA3_ROI_thickness, rh_R_STSda_ROI_thickness, rh_R_STSdp_ROI_thickness, rh_R_STSvp_ROI_thickness, rh_R_TGd_ROI_thickness, rh_R_TE1a_ROI_thickness, rh_R_TE1p_ROI_thickness, rh_R_TE2a_ROI_thickness, rh_R_TF_ROI_thickness, rh_R_TE2p_ROI_thickness, rh_R_PHT_ROI_thickness, rh_R_PH_ROI_thickness, rh_R_TPOJ1_ROI_thickness, rh_R_TPOJ2_ROI_thickness, rh_R_TPOJ3_ROI_thickness, rh_R_DVT_ROI_thickness, rh_R_PGp_ROI_thickness, rh_R_IP2_ROI_thickness, rh_R_IP1_ROI_thickness, rh_R_IP0_ROI_thickness, rh_R_PFop_ROI_thickness, rh_R_PF_ROI_thickness, rh_R_PFm_ROI_thickness, rh_R_PGi_ROI_thickness, rh_R_PGs_ROI_thickness, rh_R_V6A_ROI_thickness, rh_R_VMV1_ROI_thickness, rh_R_VMV3_ROI_thickness, rh_R_PHA2_ROI_thickness, rh_R_V4t_ROI_thickness, rh_R_FST_ROI_thickness, rh_R_V3CD_ROI_thickness, rh_R_LO3_ROI_thickness, rh_R_VMV2_ROI_thickness, rh_R_31pd_ROI_thickness, rh_R_31a_ROI_thickness, rh_R_VVC_ROI_thickness, rh_R_25_ROI_thickness, rh_R_s32_ROI_thickness, rh_R_pOFC_ROI_thickness, rh_R_PoI1_ROI_thickness, rh_R_Ig_ROI_thickness, rh_R_FOP5_ROI_thickness, rh_R_p10p_ROI_thickness, rh_R_p47r_ROI_thickness, rh_R_TGv_ROI_thickness, rh_R_MBelt_ROI_thickness, rh_R_LBelt_ROI_thickness, rh_R_A4_ROI_thickness, rh_R_STSva_ROI_thickness, rh_R_TE1m_ROI_thickness, rh_R_PI_ROI_thickness, rh_R_a32pr_ROI_thickness, rh_R_p24_ROI_thickness, rh_MeanThickness_thickness, lh_L_V1_ROI_volume, lh_L_MST_ROI_volume, lh_L_V6_ROI_volume, lh_L_V2_ROI_volume, lh_L_V3_ROI_volume, lh_L_V4_ROI_volume, lh_L_V8_ROI_volume, lh_L_4_ROI_volume, lh_L_3b_ROI_volume, lh_L_FEF_ROI_volume, lh_L_PEF_ROI_volume, lh_L_55b_ROI_volume, lh_L_V3A_ROI_volume, lh_L_RSC_ROI_volume, lh_L_POS2_ROI_volume, lh_L_V7_ROI_volume, lh_L_IPS1_ROI_volume, lh_L_FFC_ROI_volume, lh_L_V3B_ROI_volume, lh_L_LO1_ROI_volume, lh_L_LO2_ROI_volume, lh_L_PIT_ROI_volume, lh_L_MT_ROI_volume, lh_L_A1_ROI_volume, lh_L_PSL_ROI_volume, lh_L_SFL_ROI_volume, lh_L_PCV_ROI_volume, lh_L_STV_ROI_volume, lh_L_7Pm_ROI_volume, lh_L_7m_ROI_volume, lh_L_POS1_ROI_volume, lh_L_23d_ROI_volume, lh_L_v23ab_ROI_volume, lh_L_d23ab_ROI_volume, lh_L_31pv_ROI_volume, lh_L_5m_ROI_volume, lh_L_5mv_ROI_volume, lh_L_23c_ROI_volume, lh_L_5L_ROI_volume, lh_L_24dd_ROI_volume, lh_L_24dv_ROI_volume, lh_L_7AL_ROI_volume, lh_L_SCEF_ROI_volume, lh_L_6ma_ROI_volume, lh_L_7Am_ROI_volume, lh_L_7PL_ROI_volume, lh_L_7PC_ROI_volume, lh_L_LIPv_ROI_volume, lh_L_VIP_ROI_volume, lh_L_MIP_ROI_volume, lh_L_1_ROI_volume, lh_L_2_ROI_volume, lh_L_3a_ROI_volume, lh_L_6d_ROI_volume, lh_L_6mp_ROI_volume, lh_L_6v_ROI_volume, lh_L_p24pr_ROI_volume, lh_L_33pr_ROI_volume, lh_L_a24pr_ROI_volume, lh_L_p32pr_ROI_volume, lh_L_a24_ROI_volume, lh_L_d32_ROI_volume, lh_L_8BM_ROI_volume, lh_L_p32_ROI_volume, lh_L_10r_ROI_volume, lh_L_47m_ROI_volume, lh_L_8Av_ROI_volume, lh_L_8Ad_ROI_volume, lh_L_9m_ROI_volume, lh_L_8BL_ROI_volume, lh_L_9p_ROI_volume, lh_L_10d_ROI_volume, lh_L_8C_ROI_volume, lh_L_44_ROI_volume, lh_L_45_ROI_volume, lh_L_47l_ROI_volume, lh_L_a47r_ROI_volume, lh_L_6r_ROI_volume, lh_L_IFJa_ROI_volume, lh_L_IFJp_ROI_volume, lh_L_IFSp_ROI_volume, lh_L_IFSa_ROI_volume, lh_L_p9-46v_ROI_volume, lh_L_46_ROI_volume, lh_L_a9-46v_ROI_volume, lh_L_9-46d_ROI_volume, lh_L_9a_ROI_volume, lh_L_10v_ROI_volume, lh_L_a10p_ROI_volume, lh_L_10pp_ROI_volume, lh_L_11l_ROI_volume, lh_L_13l_ROI_volume, lh_L_OFC_ROI_volume, lh_L_47s_ROI_volume, lh_L_LIPd_ROI_volume, lh_L_6a_ROI_volume, lh_L_i6-8_ROI_volume, lh_L_s6-8_ROI_volume, lh_L_43_ROI_volume, lh_L_OP4_ROI_volume, lh_L_OP1_ROI_volume, lh_L_OP2-3_ROI_volume, lh_L_52_ROI_volume, lh_L_RI_ROI_volume, lh_L_PFcm_ROI_volume, lh_L_PoI2_ROI_volume, lh_L_TA2_ROI_volume, lh_L_FOP4_ROI_volume, lh_L_MI_ROI_volume, lh_L_Pir_ROI_volume, lh_L_AVI_ROI_volume, lh_L_AAIC_ROI_volume, lh_L_FOP1_ROI_volume, lh_L_FOP3_ROI_volume, lh_L_FOP2_ROI_volume, lh_L_PFt_ROI_volume, lh_L_AIP_ROI_volume, lh_L_EC_ROI_volume, lh_L_PreS_ROI_volume, lh_L_H_ROI_volume, lh_L_ProS_ROI_volume, lh_L_PeEc_ROI_volume, lh_L_STGa_ROI_volume, lh_L_PBelt_ROI_volume, lh_L_A5_ROI_volume, lh_L_PHA1_ROI_volume, lh_L_PHA3_ROI_volume, lh_L_STSda_ROI_volume, lh_L_STSdp_ROI_volume, lh_L_STSvp_ROI_volume, lh_L_TGd_ROI_volume, lh_L_TE1a_ROI_volume, lh_L_TE1p_ROI_volume, lh_L_TE2a_ROI_volume, lh_L_TF_ROI_volume, lh_L_TE2p_ROI_volume, lh_L_PHT_ROI_volume, lh_L_PH_ROI_volume, lh_L_TPOJ1_ROI_volume, lh_L_TPOJ2_ROI_volume, lh_L_TPOJ3_ROI_volume, lh_L_DVT_ROI_volume, lh_L_PGp_ROI_volume, lh_L_IP2_ROI_volume, lh_L_IP1_ROI_volume, lh_L_IP0_ROI_volume, lh_L_PFop_ROI_volume, lh_L_PF_ROI_volume, lh_L_PFm_ROI_volume, lh_L_PGi_ROI_volume, lh_L_PGs_ROI_volume, lh_L_V6A_ROI_volume, lh_L_VMV1_ROI_volume, lh_L_VMV3_ROI_volume, lh_L_PHA2_ROI_volume, lh_L_V4t_ROI_volume, lh_L_FST_ROI_volume, lh_L_V3CD_ROI_volume, lh_L_LO3_ROI_volume, lh_L_VMV2_ROI_volume, lh_L_31pd_ROI_volume, lh_L_31a_ROI_volume, lh_L_VVC_ROI_volume, lh_L_25_ROI_volume, lh_L_s32_ROI_volume, lh_L_pOFC_ROI_volume, lh_L_PoI1_ROI_volume, lh_L_Ig_ROI_volume, lh_L_FOP5_ROI_volume, lh_L_p10p_ROI_volume, lh_L_p47r_ROI_volume, lh_L_TGv_ROI_volume, lh_L_MBelt_ROI_volume, lh_L_LBelt_ROI_volume, lh_L_A4_ROI_volume, lh_L_STSva_ROI_volume, lh_L_TE1m_ROI_volume, lh_L_PI_ROI_volume, lh_L_a32pr_ROI_volume, lh_L_p24_ROI_volume, rh_R_V1_ROI_volume, rh_R_MST_ROI_volume, rh_R_V6_ROI_volume, rh_R_V2_ROI_volume, rh_R_V3_ROI_volume, rh_R_V4_ROI_volume, rh_R_V8_ROI_volume, rh_R_4_ROI_volume, rh_R_3b_ROI_volume, rh_R_FEF_ROI_volume, rh_R_PEF_ROI_volume, rh_R_55b_ROI_volume, rh_R_V3A_ROI_volume, rh_R_RSC_ROI_volume, rh_R_POS2_ROI_volume, rh_R_V7_ROI_volume, rh_R_IPS1_ROI_volume, rh_R_FFC_ROI_volume, rh_R_V3B_ROI_volume, rh_R_LO1_ROI_volume, rh_R_LO2_ROI_volume, rh_R_PIT_ROI_volume, rh_R_MT_ROI_volume, rh_R_A1_ROI_volume, rh_R_PSL_ROI_volume, rh_R_SFL_ROI_volume, rh_R_PCV_ROI_volume, rh_R_STV_ROI_volume, rh_R_7Pm_ROI_volume, rh_R_7m_ROI_volume, rh_R_POS1_ROI_volume, rh_R_23d_ROI_volume, rh_R_v23ab_ROI_volume, rh_R_d23ab_ROI_volume, rh_R_31pv_ROI_volume, rh_R_5m_ROI_volume, rh_R_5mv_ROI_volume, rh_R_23c_ROI_volume, rh_R_5L_ROI_volume, rh_R_24dd_ROI_volume, rh_R_24dv_ROI_volume, rh_R_7AL_ROI_volume, rh_R_SCEF_ROI_volume, rh_R_6ma_ROI_volume, rh_R_7Am_ROI_volume, rh_R_7PL_ROI_volume, rh_R_7PC_ROI_volume, rh_R_LIPv_ROI_volume, rh_R_VIP_ROI_volume, rh_R_MIP_ROI_volume, rh_R_1_ROI_volume, rh_R_2_ROI_volume, rh_R_3a_ROI_volume, rh_R_6d_ROI_volume, rh_R_6mp_ROI_volume, rh_R_6v_ROI_volume, rh_R_p24pr_ROI_volume, rh_R_33pr_ROI_volume, rh_R_a24pr_ROI_volume, rh_R_p32pr_ROI_volume, rh_R_a24_ROI_volume, rh_R_d32_ROI_volume, rh_R_8BM_ROI_volume, rh_R_p32_ROI_volume, rh_R_10r_ROI_volume, rh_R_47m_ROI_volume, rh_R_8Av_ROI_volume, rh_R_8Ad_ROI_volume, rh_R_9m_ROI_volume, rh_R_8BL_ROI_volume, rh_R_9p_ROI_volume, rh_R_10d_ROI_volume, rh_R_8C_ROI_volume, rh_R_44_ROI_volume, rh_R_45_ROI_volume, rh_R_47l_ROI_volume, rh_R_a47r_ROI_volume, rh_R_6r_ROI_volume, rh_R_IFJa_ROI_volume, rh_R_IFJp_ROI_volume, rh_R_IFSp_ROI_volume, rh_R_IFSa_ROI_volume, rh_R_p9-46v_ROI_volume, rh_R_46_ROI_volume, rh_R_a9-46v_ROI_volume, rh_R_9-46d_ROI_volume, rh_R_9a_ROI_volume, rh_R_10v_ROI_volume, rh_R_a10p_ROI_volume, rh_R_10pp_ROI_volume, rh_R_11l_ROI_volume, rh_R_13l_ROI_volume, rh_R_OFC_ROI_volume, rh_R_47s_ROI_volume, rh_R_LIPd_ROI_volume, rh_R_6a_ROI_volume, rh_R_i6-8_ROI_volume, rh_R_s6-8_ROI_volume, rh_R_43_ROI_volume, rh_R_OP4_ROI_volume, rh_R_OP1_ROI_volume, rh_R_OP2-3_ROI_volume, rh_R_52_ROI_volume, rh_R_RI_ROI_volume, rh_R_PFcm_ROI_volume, rh_R_PoI2_ROI_volume, rh_R_TA2_ROI_volume, rh_R_FOP4_ROI_volume, rh_R_MI_ROI_volume, rh_R_Pir_ROI_volume, rh_R_AVI_ROI_volume, rh_R_AAIC_ROI_volume, rh_R_FOP1_ROI_volume, rh_R_FOP3_ROI_volume, rh_R_FOP2_ROI_volume, rh_R_PFt_ROI_volume, rh_R_AIP_ROI_volume, rh_R_EC_ROI_volume, rh_R_PreS_ROI_volume, rh_R_H_ROI_volume, rh_R_ProS_ROI_volume, rh_R_PeEc_ROI_volume, rh_R_STGa_ROI_volume, rh_R_PBelt_ROI_volume, rh_R_A5_ROI_volume, rh_R_PHA1_ROI_volume, rh_R_PHA3_ROI_volume, rh_R_STSda_ROI_volume, rh_R_STSdp_ROI_volume, rh_R_STSvp_ROI_volume, rh_R_TGd_ROI_volume, rh_R_TE1a_ROI_volume, rh_R_TE1p_ROI_volume, rh_R_TE2a_ROI_volume, rh_R_TF_ROI_volume, rh_R_TE2p_ROI_volume, rh_R_PHT_ROI_volume, rh_R_PH_ROI_volume, rh_R_TPOJ1_ROI_volume, rh_R_TPOJ2_ROI_volume, rh_R_TPOJ3_ROI_volume, rh_R_DVT_ROI_volume, rh_R_PGp_ROI_volume, rh_R_IP2_ROI_volume, rh_R_IP1_ROI_volume, rh_R_IP0_ROI_volume, rh_R_PFop_ROI_volume, rh_R_PF_ROI_volume, rh_R_PFm_ROI_volume, rh_R_PGi_ROI_volume, rh_R_PGs_ROI_volume, rh_R_V6A_ROI_volume, rh_R_VMV1_ROI_volume, rh_R_VMV3_ROI_volume, rh_R_PHA2_ROI_volume, rh_R_V4t_ROI_volume, rh_R_FST_ROI_volume, rh_R_V3CD_ROI_volume, rh_R_LO3_ROI_volume, rh_R_VMV2_ROI_volume, rh_R_31pd_ROI_volume, rh_R_31a_ROI_volume, rh_R_VVC_ROI_volume, rh_R_25_ROI_volume, rh_R_s32_ROI_volume, rh_R_pOFC_ROI_volume, rh_R_PoI1_ROI_volume, rh_R_Ig_ROI_volume, rh_R_FOP5_ROI_volume, rh_R_p10p_ROI_volume, rh_R_p47r_ROI_volume, rh_R_TGv_ROI_volume, rh_R_MBelt_ROI_volume, rh_R_LBelt_ROI_volume, rh_R_A4_ROI_volume, rh_R_STSva_ROI_volume, rh_R_TE1m_ROI_volume, rh_R_PI_ROI_volume, rh_R_a32pr_ROI_volume, rh_R_p24_ROI_volume, lh_L_V1_ROI_area, lh_L_MST_ROI_area, lh_L_V6_ROI_area, lh_L_V2_ROI_area, lh_L_V3_ROI_area, lh_L_V4_ROI_area, lh_L_V8_ROI_area, lh_L_4_ROI_area, lh_L_3b_ROI_area, lh_L_FEF_ROI_area, lh_L_PEF_ROI_area, lh_L_55b_ROI_area, lh_L_V3A_ROI_area, lh_L_RSC_ROI_area, lh_L_POS2_ROI_area, lh_L_V7_ROI_area, lh_L_IPS1_ROI_area, lh_L_FFC_ROI_area, lh_L_V3B_ROI_area, lh_L_LO1_ROI_area, lh_L_LO2_ROI_area, lh_L_PIT_ROI_area, lh_L_MT_ROI_area, lh_L_A1_ROI_area, lh_L_PSL_ROI_area, lh_L_SFL_ROI_area, lh_L_PCV_ROI_area, lh_L_STV_ROI_area, lh_L_7Pm_ROI_area, lh_L_7m_ROI_area, lh_L_POS1_ROI_area, lh_L_23d_ROI_area, lh_L_v23ab_ROI_area, lh_L_d23ab_ROI_area, lh_L_31pv_ROI_area, lh_L_5m_ROI_area, lh_L_5mv_ROI_area, lh_L_23c_ROI_area, lh_L_5L_ROI_area, lh_L_24dd_ROI_area, lh_L_24dv_ROI_area, lh_L_7AL_ROI_area, lh_L_SCEF_ROI_area, lh_L_6ma_ROI_area, lh_L_7Am_ROI_area, lh_L_7PL_ROI_area, lh_L_7PC_ROI_area, lh_L_LIPv_ROI_area, lh_L_VIP_ROI_area, lh_L_MIP_ROI_area, lh_L_1_ROI_area, lh_L_2_ROI_area, lh_L_3a_ROI_area, lh_L_6d_ROI_area, lh_L_6mp_ROI_area, lh_L_6v_ROI_area, lh_L_p24pr_ROI_area, lh_L_33pr_ROI_area, lh_L_a24pr_ROI_area, lh_L_p32pr_ROI_area, lh_L_a24_ROI_area, lh_L_d32_ROI_area, lh_L_8BM_ROI_area, lh_L_p32_ROI_area, lh_L_10r_ROI_area, lh_L_47m_ROI_area, lh_L_8Av_ROI_area, lh_L_8Ad_ROI_area, lh_L_9m_ROI_area, lh_L_8BL_ROI_area, lh_L_9p_ROI_area, lh_L_10d_ROI_area, lh_L_8C_ROI_area, lh_L_44_ROI_area, lh_L_45_ROI_area, lh_L_47l_ROI_area, lh_L_a47r_ROI_area, lh_L_6r_ROI_area, lh_L_IFJa_ROI_area, lh_L_IFJp_ROI_area, lh_L_IFSp_ROI_area, lh_L_IFSa_ROI_area, lh_L_p9-46v_ROI_area, lh_L_46_ROI_area, lh_L_a9-46v_ROI_area, lh_L_9-46d_ROI_area, lh_L_9a_ROI_area, lh_L_10v_ROI_area, lh_L_a10p_ROI_area, lh_L_10pp_ROI_area, lh_L_11l_ROI_area, lh_L_13l_ROI_area, lh_L_OFC_ROI_area, lh_L_47s_ROI_area, lh_L_LIPd_ROI_area, lh_L_6a_ROI_area, lh_L_i6-8_ROI_area, lh_L_s6-8_ROI_area, lh_L_43_ROI_area, lh_L_OP4_ROI_area, lh_L_OP1_ROI_area, lh_L_OP2-3_ROI_area, lh_L_52_ROI_area, lh_L_RI_ROI_area, lh_L_PFcm_ROI_area, lh_L_PoI2_ROI_area, lh_L_TA2_ROI_area, lh_L_FOP4_ROI_area, lh_L_MI_ROI_area, lh_L_Pir_ROI_area, lh_L_AVI_ROI_area, lh_L_AAIC_ROI_area, lh_L_FOP1_ROI_area, lh_L_FOP3_ROI_area, lh_L_FOP2_ROI_area, lh_L_PFt_ROI_area, lh_L_AIP_ROI_area, lh_L_EC_ROI_area, lh_L_PreS_ROI_area, lh_L_H_ROI_area, lh_L_ProS_ROI_area, lh_L_PeEc_ROI_area, lh_L_STGa_ROI_area, lh_L_PBelt_ROI_area, lh_L_A5_ROI_area, lh_L_PHA1_ROI_area, lh_L_PHA3_ROI_area, lh_L_STSda_ROI_area, lh_L_STSdp_ROI_area, lh_L_STSvp_ROI_area, lh_L_TGd_ROI_area, lh_L_TE1a_ROI_area, lh_L_TE1p_ROI_area, lh_L_TE2a_ROI_area, lh_L_TF_ROI_area, lh_L_TE2p_ROI_area, lh_L_PHT_ROI_area, lh_L_PH_ROI_area, lh_L_TPOJ1_ROI_area, lh_L_TPOJ2_ROI_area, lh_L_TPOJ3_ROI_area, lh_L_DVT_ROI_area, lh_L_PGp_ROI_area, lh_L_IP2_ROI_area, lh_L_IP1_ROI_area, lh_L_IP0_ROI_area, lh_L_PFop_ROI_area, lh_L_PF_ROI_area, lh_L_PFm_ROI_area, lh_L_PGi_ROI_area, lh_L_PGs_ROI_area, lh_L_V6A_ROI_area, lh_L_VMV1_ROI_area, lh_L_VMV3_ROI_area, lh_L_PHA2_ROI_area, lh_L_V4t_ROI_area, lh_L_FST_ROI_area, lh_L_V3CD_ROI_area, lh_L_LO3_ROI_area, lh_L_VMV2_ROI_area, lh_L_31pd_ROI_area, lh_L_31a_ROI_area, lh_L_VVC_ROI_area, lh_L_25_ROI_area, lh_L_s32_ROI_area, lh_L_pOFC_ROI_area, lh_L_PoI1_ROI_area, lh_L_Ig_ROI_area, lh_L_FOP5_ROI_area, lh_L_p10p_ROI_area, lh_L_p47r_ROI_area, lh_L_TGv_ROI_area, lh_L_MBelt_ROI_area, lh_L_LBelt_ROI_area, lh_L_A4_ROI_area, lh_L_STSva_ROI_area, lh_L_TE1m_ROI_area, lh_L_PI_ROI_area, lh_L_a32pr_ROI_area, lh_L_p24_ROI_area, rh_R_V1_ROI_area, rh_R_MST_ROI_area, rh_R_V6_ROI_area, rh_R_V2_ROI_area, rh_R_V3_ROI_area, rh_R_V4_ROI_area, rh_R_V8_ROI_area, rh_R_4_ROI_area, rh_R_3b_ROI_area, rh_R_FEF_ROI_area, rh_R_PEF_ROI_area, rh_R_55b_ROI_area, rh_R_V3A_ROI_area, rh_R_RSC_ROI_area, rh_R_POS2_ROI_area, rh_R_V7_ROI_area, rh_R_IPS1_ROI_area, rh_R_FFC_ROI_area, rh_R_V3B_ROI_area, rh_R_LO1_ROI_area, rh_R_LO2_ROI_area, rh_R_PIT_ROI_area, rh_R_MT_ROI_area, rh_R_A1_ROI_area, rh_R_PSL_ROI_area, rh_R_SFL_ROI_area, rh_R_PCV_ROI_area, rh_R_STV_ROI_area, rh_R_7Pm_ROI_area, rh_R_7m_ROI_area, rh_R_POS1_ROI_area, rh_R_23d_ROI_area, rh_R_v23ab_ROI_area, rh_R_d23ab_ROI_area, rh_R_31pv_ROI_area, rh_R_5m_ROI_area, rh_R_5mv_ROI_area, rh_R_23c_ROI_area, rh_R_5L_ROI_area, rh_R_24dd_ROI_area, rh_R_24dv_ROI_area, rh_R_7AL_ROI_area, rh_R_SCEF_ROI_area, rh_R_6ma_ROI_area, rh_R_7Am_ROI_area, rh_R_7PL_ROI_area, rh_R_7PC_ROI_area, rh_R_LIPv_ROI_area, rh_R_VIP_ROI_area, rh_R_MIP_ROI_area, rh_R_1_ROI_area, rh_R_2_ROI_area, rh_R_3a_ROI_area, rh_R_6d_ROI_area, rh_R_6mp_ROI_area, rh_R_6v_ROI_area, rh_R_p24pr_ROI_area, rh_R_33pr_ROI_area, rh_R_a24pr_ROI_area, rh_R_p32pr_ROI_area, rh_R_a24_ROI_area, rh_R_d32_ROI_area, rh_R_8BM_ROI_area, rh_R_p32_ROI_area, rh_R_10r_ROI_area, rh_R_47m_ROI_area, rh_R_8Av_ROI_area, rh_R_8Ad_ROI_area, rh_R_9m_ROI_area, rh_R_8BL_ROI_area, rh_R_9p_ROI_area, rh_R_10d_ROI_area, rh_R_8C_ROI_area, rh_R_44_ROI_area, rh_R_45_ROI_area, rh_R_47l_ROI_area, rh_R_a47r_ROI_area, rh_R_6r_ROI_area, rh_R_IFJa_ROI_area, rh_R_IFJp_ROI_area, rh_R_IFSp_ROI_area, rh_R_IFSa_ROI_area, rh_R_p9-46v_ROI_area, rh_R_46_ROI_area, rh_R_a9-46v_ROI_area, rh_R_9-46d_ROI_area, rh_R_9a_ROI_area, rh_R_10v_ROI_area, rh_R_a10p_ROI_area, rh_R_10pp_ROI_area, rh_R_11l_ROI_area, rh_R_13l_ROI_area, rh_R_OFC_ROI_area, rh_R_47s_ROI_area, rh_R_LIPd_ROI_area, rh_R_6a_ROI_area, rh_R_i6-8_ROI_area, rh_R_s6-8_ROI_area, rh_R_43_ROI_area, rh_R_OP4_ROI_area, rh_R_OP1_ROI_area, rh_R_OP2-3_ROI_area, rh_R_52_ROI_area, rh_R_RI_ROI_area, rh_R_PFcm_ROI_area, rh_R_PoI2_ROI_area, rh_R_TA2_ROI_area, rh_R_FOP4_ROI_area, rh_R_MI_ROI_area, rh_R_Pir_ROI_area, rh_R_AVI_ROI_area, rh_R_AAIC_ROI_area, rh_R_FOP1_ROI_area, rh_R_FOP3_ROI_area, rh_R_FOP2_ROI_area, rh_R_PFt_ROI_area, rh_R_AIP_ROI_area, rh_R_EC_ROI_area, rh_R_PreS_ROI_area, rh_R_H_ROI_area, rh_R_ProS_ROI_area, rh_R_PeEc_ROI_area, rh_R_STGa_ROI_area, rh_R_PBelt_ROI_area, rh_R_A5_ROI_area, rh_R_PHA1_ROI_area, rh_R_PHA3_ROI_area, rh_R_STSda_ROI_area, rh_R_STSdp_ROI_area, rh_R_STSvp_ROI_area, rh_R_TGd_ROI_area, rh_R_TE1a_ROI_area, rh_R_TE1p_ROI_area, rh_R_TE2a_ROI_area, rh_R_TF_ROI_area, rh_R_TE2p_ROI_area, rh_R_PHT_ROI_area, rh_R_PH_ROI_area, rh_R_TPOJ1_ROI_area, rh_R_TPOJ2_ROI_area, rh_R_TPOJ3_ROI_area, rh_R_DVT_ROI_area, rh_R_PGp_ROI_area, rh_R_IP2_ROI_area, rh_R_IP1_ROI_area, rh_R_IP0_ROI_area, rh_R_PFop_ROI_area, rh_R_PF_ROI_area, rh_R_PFm_ROI_area, rh_R_PGi_ROI_area, rh_R_PGs_ROI_area, rh_R_V6A_ROI_area, rh_R_VMV1_ROI_area, rh_R_VMV3_ROI_area, rh_R_PHA2_ROI_area, rh_R_V4t_ROI_area, rh_R_FST_ROI_area, rh_R_V3CD_ROI_area, rh_R_LO3_ROI_area, rh_R_VMV2_ROI_area, rh_R_31pd_ROI_area, rh_R_31a_ROI_area, rh_R_VVC_ROI_area, rh_R_25_ROI_area, rh_R_s32_ROI_area, rh_R_pOFC_ROI_area, rh_R_PoI1_ROI_area, rh_R_Ig_ROI_area, rh_R_FOP5_ROI_area, rh_R_p10p_ROI_area, rh_R_p47r_ROI_area, rh_R_TGv_ROI_area, rh_R_MBelt_ROI_area, rh_R_LBelt_ROI_area, rh_R_A4_ROI_area, rh_R_STSva_ROI_area, rh_R_TE1m_ROI_area, rh_R_PI_ROI_area, rh_R_a32pr_ROI_area, rh_R_p24_ROI_area, Left-Lateral-Ventricle, Left-Inf-Lat-Vent, Left-Cerebellum-White-Matter, Left-Cerebellum-Cortex, Left-Thalamus-Proper, Left-Caudate, Left-Putamen, Left-Pallidum, 3rd-Ventricle, 4th-Ventricle, Brain-Stem, Left-Hippocampus, Left-Amygdala, Left-Accumbens-area, Right-Lateral-Ventricle, Right-Inf-Lat-Vent, Right-Cerebellum-White-Matter, Right-Cerebellum-Cortex, Right-Thalamus-Proper, Right-Caudate, Right-Putamen, Right-Pallidum, Right-Hippocampus, Right-Amygdala, Right-Accumbens-area, CC_Posterior, CC_Mid_Posterior, CC_Central, CC_Mid_Anterior, CC_Anterior, lhCortexVol, rhCortexVol, lhCorticalWhiteMatterVol, rhCorticalWhiteMatterVol, TotalGrayVol, SupraTentorialVol |
| dMRI | FA_ATRL, FA_ATRR, FA_CSTL, FA_CSTR, FA_CGL, FA_CGR, FA_FMAJ, FA_FMIN, FA_IFOFL, FA_IFOFR, FA_ILFL, FA_ILFR, FA_SLFL, FA_SLFR, FA_UFL, FA_UFR, FA_SLFTL, FA_SLTFR, FA_Middlecerebellarpeduncle, FA_Pontine, FA_GenuCC, FA_BodyCC, FA_SpleniumCC, FA_Fornix, FA_CorticospinaltractR, FA_CorticospinaltractL, FA_MediallemniscusR, FA_MediallemniscusL, FA_InferiorcerebellarpeduncleR, FA_InferiorcerebellarpeduncleL, FA_SuperiorcerebellarpeduncleR, FA_SuperiorcerebellarpeduncleL, FA_CerebralpeduncleR, FA_CerebralpeduncleL, FA_AnteriorlimbofinternalcapsuleR, FA_AnteriorlimbofinternalcapsuleL, FA_ PosteriorlimbofinternalcapsuleR, FA_PosteriorlimbofinternalcapsuleL, FA_RetrolenticularpartofinternalcapsuleR, FA_RetrolenticularpartofinternalcapsuleL, FA_AnteriorcoronaradiataR, FA_AnteriorcoronaradiataL, FA_SuperiorcoronaradiataR, FA_SuperiorcoronaradiataL, FA_PosteriorcoronaradiataR, FA_PosteriorcoronaradiataL, FA_PosteriorthalamicradiationR, FA_PosteriorthalamicradiationL, FA_SagittalstratumR, FA_SagittalstratumL, FA_ExternalcapsuleR, FA_ExternalcapsuleL, FA_CingulumcingulategyrusR, FA_CingulumcingulategyrusL, FA_CingulumhippocampusR, FA_CingulumhippocampusL, FA_Fornix_StriaterminalisR, FA_Fornix_StriaterminalisL, FA_SuperiorlongitudinalfasciculusR, FA_SuperiorlongitudinalfasciculusL, FA_SuperiorfrontooccipitalfasciculusR, FA_SuperiorfrontooccipitalfasciculusL, FA_UncinatefasciculusR, FA_UncinatefasciculusL, FA_TapetumR, FA_TapetumL, FA_Mean, FA_SD, FA_CINGL, FA_CINGR, MD_ATRL, MD_ATRR, MD_CSTL, MD_CSTR, MD_CGL, MD_CGR, MD_FMAJ, MD_FMIN, MD_IFOFL, MD_IFOFR, MD_ILFL, MD_ILFR, MD_SLFL, MD_SLFR, MD_UFL, MD_UFR, MD_SLFTL, MD_SLTFR, MD_Middlecerebellarpeduncle, MD_Pontine, MD_GenuCC, MD_BodyCC, MD_SpleniumCC, MD_Fornix, MD_CorticospinaltractR, MD_CorticospinaltractL, MD_MediallemniscusR, MD_MediallemniscusL, MD_InferiorcerebellarpeduncleR, MD_InferiorcerebellarpeduncleL, MD_SuperiorcerebellarpeduncleR, MD_SuperiorcerebellarpeduncleL, MD_CerebralpeduncleR, MD_CerebralpeduncleL, MD_AnteriorlimbofinternalcapsuleR, MD_AnteriorlimbofinternalcapsuleL, MD_ PosteriorlimbofinternalcapsuleR, MD_PosteriorlimbofinternalcapsuleL, MD_RetrolenticularpartofinternalcapsuleR, MD_RetrolenticularpartofinternalcapsuleL, MD_AnteriorcoronaradiataR, MD_AnteriorcoronaradiataL, MD_SuperiorcoronaradiataR, MD_SuperiorcoronaradiataL, MD_PosteriorcoronaradiataR, MD_PosteriorcoronaradiataL, MD_PosteriorthalamicradiationR, MD_PosteriorthalamicradiationL, MD_SagittalstratumR, MD_SagittalstratumL, MD_ExternalcapsuleR, MD_ExternalcapsuleL, MD_CingulumcingulategyrusR, MD_CingulumcingulategyrusL, MD_CingulumhippocampusR, MD_CingulumhippocampusL, MD_Fornix_StriaterminalisR, MD_Fornix_StriaterminalisL, MD_SuperiorlongitudinalfasciculusR, MD_SuperiorlongitudinalfasciculusL, MD_SuperiorfrontooccipitalfasciculusR, MD_SuperiorfrontooccipitalfasciculusL, MD_UncinatefasciculusR, MD_UncinatefasciculusL, MD_TapetumR, MD_TapetumL, MD_Mean, MD_SD, MD_CINGL, MD_CINGR, RD_ATRL, RD_ATRR, RD_CSTL, RD_CSTR, RD_CGL, RD_CGR, RD_FMAJ, RD_FMIN, RD_IFOFL, RD_IFOFR, RD_ILFL, RD_ILFR, RD_SLFL, RD_SLFR, RD_UFL, RD_UFR, RD_SLFTL, RD_SLTFR, RD_Middlecerebellarpeduncle, RD_Pontine, RD_GenuCC, RD_BodyCC, RD_SpleniumCC, RD_Fornix, RD_CorticospinaltractR, RD_CorticospinaltractL, RD_MediallemniscusR, RD_MediallemniscusL, RD_InferiorcerebellarpeduncleR, RD_InferiorcerebellarpeduncleL, RD_SuperiorcerebellarpeduncleR, RD_SuperiorcerebellarpeduncleL, RD_CerebralpeduncleR, RD_CerebralpeduncleL, RD_AnteriorlimbofinternalcapsuleR, RD_AnteriorlimbofinternalcapsuleL, RD_ PosteriorlimbofinternalcapsuleR, RD_PosteriorlimbofinternalcapsuleL, RD_RetrolenticularpartofinternalcapsuleR, RD_RetrolenticularpartofinternalcapsuleL, RD_AnteriorcoronaradiataR, RD_AnteriorcoronaradiataL, RD_SuperiorcoronaradiataR, RD_SuperiorcoronaradiataL, RD_PosteriorcoronaradiataR, RD_PosteriorcoronaradiataL, RD_PosteriorthalamicradiationR, RD_PosteriorthalamicradiationL, RD_SagittalstratumR, RD_SagittalstratumL, RD_ExternalcapsuleR, RD_ExternalcapsuleL, RD_CingulumcingulategyrusR, RD_CingulumcingulategyrusL, RD_CingulumhippocampusR, RD_CingulumhippocampusL, RD_Fornix_StriaterminalisR, RD_Fornix_StriaterminalisL, RD_SuperiorlongitudinalfasciculusR, RD_SuperiorlongitudinalfasciculusL, RD_SuperiorfrontooccipitalfasciculusR, RD_SuperiorfrontooccipitalfasciculusL, RD_UncinatefasciculusR, RD_UncinatefasciculusL, RD_TapetumR, RD_TapetumL, RD_Mean, RD_SD, RD_CINGL, RD_CINGR, AD_ATRL, AD_ATRR, AD_CSTL, AD_CSTR, AD_CGL, AD_CGR, AD_FMAJ, AD_FMIN, AD_IFOFL, AD_IFOFR, AD_ILFL, AD_ILFR, AD_SLFL, AD_SLFR, AD_UFL, AD_UFR, AD_SLFTL, AD_SLTFR, AD_Middlecerebellarpeduncle, AD_Pontine, AD_GenuCC, AD_BodyCC, AD_SpleniumCC, AD_Fornix, AD_CorticospinaltractR, AD_CorticospinaltractL, AD_MediallemniscusR, AD_MediallemniscusL, AD_InferiorcerebellarpeduncleR, AD_InferiorcerebellarpeduncleL, AD_SuperiorcerebellarpeduncleR, AD_SuperiorcerebellarpeduncleL, AD_CerebralpeduncleR, AD_CerebralpeduncleL, AD_AnteriorlimbofinternalcapsuleR, AD_AnteriorlimbofinternalcapsuleL, AD_ PosteriorlimbofinternalcapsuleR, AD_PosteriorlimbofinternalcapsuleL, AD_RetrolenticularpartofinternalcapsuleR, AD_RetrolenticularpartofinternalcapsuleL, AD_AnteriorcoronaradiataR, AD_AnteriorcoronaradiataL, AD_SuperiorcoronaradiataR, AD_SuperiorcoronaradiataL, AD_PosteriorcoronaradiataR, AD_PosteriorcoronaradiataL, AD_PosteriorthalamicradiationR, AD_PosteriorthalamicradiationL, AD_SagittalstratumR, AD_SagittalstratumL, AD_ExternalcapsuleR, AD_ExternalcapsuleL, AD_CingulumcingulategyrusR, AD_CingulumcingulategyrusL, AD_CingulumhippocampusR, AD_CingulumhippocampusL, AD_Fornix_StriaterminalisR, AD_Fornix_StriaterminalisL, AD_SuperiorlongitudinalfasciculusR, AD_SuperiorlongitudinalfasciculusL, AD_SuperiorfrontooccipitalfasciculusR, AD_SuperiorfrontooccipitalfasciculusL, AD_UncinatefasciculusR, AD_UncinatefasciculusL, AD_TapetumR, AD_TapetumL, AD_Mean, AD_SD, AD_CINGL, AD_CINGR, axEAD_ATRL, axEAD_ATRR, axEAD_CSTL, axEAD_CSTR, axEAD_CGL, axEAD_CGR, axEAD_FMAJ, axEAD_FMIN, axEAD_IFOFL, axEAD_IFOFR, axEAD_ILFL, axEAD_ILFR, axEAD_SLFL, axEAD_SLFR, axEAD_UFL, axEAD_UFR, axEAD_SLFTL, axEAD_SLTFR, axEAD_Middlecerebellarpeduncle, axEAD_Pontine, axEAD_GenuCC, axEAD_BodyCC, axEAD_SpleniumCC, axEAD_Fornix, axEAD_CorticospinaltractR, axEAD_CorticospinaltractL, axEAD_MediallemniscusR, axEAD_MediallemniscusL, axEAD_InferiorcerebellarpeduncleR, axEAD_InferiorcerebellarpeduncleL, axEAD_SuperiorcerebellarpeduncleR, axEAD_SuperiorcerebellarpeduncleL, axEAD_CerebralpeduncleR, axEAD_CerebralpeduncleL, axEAD_AnteriorlimbofinternalcapsuleR, axEAD_AnteriorlimbofinternalcapsuleL, axEAD_ PosteriorlimbofinternalcapsuleR, axEAD_PosteriorlimbofinternalcapsuleL, axEAD_RetrolenticularpartofinternalcapsuleR, axEAD_RetrolenticularpartofinternalcapsuleL, axEAD_AnteriorcoronaradiataR, axEAD_AnteriorcoronaradiataL, axEAD_SuperiorcoronaradiataR, axEAD_SuperiorcoronaradiataL, axEAD_PosteriorcoronaradiataR, axEAD_PosteriorcoronaradiataL, axEAD_PosteriorthalamicradiationR, axEAD_PosteriorthalamicradiationL, axEAD_SagittalstratumR, axEAD_SagittalstratumL, axEAD_ExternalcapsuleR, axEAD_ExternalcapsuleL, axEAD_CingulumcingulategyrusR, axEAD_CingulumcingulategyrusL, axEAD_CingulumhippocampusR, axEAD_CingulumhippocampusL, axEAD_Fornix_StriaterminalisR, axEAD_Fornix_StriaterminalisL, axEAD_SuperiorlongitudinalfasciculusR, axEAD_SuperiorlongitudinalfasciculusL, axEAD_SuperiorfrontooccipitalfasciculusR, axEAD_SuperiorfrontooccipitalfasciculusL, axEAD_UncinatefasciculusR, axEAD_UncinatefasciculusL, axEAD_TapetumR, axEAD_TapetumL, axEAD_Mean, axEAD_SD, axEAD_CINGL, axEAD_CINGR, awf_ATRL, awf_ATRR, awf_CSTL, awf_CSTR, awf_CGL, awf_CGR, awf_FMAJ, awf_FMIN, awf_IFOFL, awf_IFOFR, awf_ILFL, awf_ILFR, awf_SLFL, awf_SLFR, awf_UFL, awf_UFR, awf_SLFTL, awf_SLTFR, awf_Middlecerebellarpeduncle, awf_Pontine, awf_GenuCC, awf_BodyCC, awf_SpleniumCC, awf_Fornix, awf_CorticospinaltractR, awf_CorticospinaltractL, awf_MediallemniscusR, awf_MediallemniscusL, awf_InferiorcerebellarpeduncleR, awf_InferiorcerebellarpeduncleL, awf_SuperiorcerebellarpeduncleR, awf_SuperiorcerebellarpeduncleL, awf_CerebralpeduncleR, awf_CerebralpeduncleL, awf_AnteriorlimbofinternalcapsuleR, awf_AnteriorlimbofinternalcapsuleL, awf_ PosteriorlimbofinternalcapsuleR, awf_PosteriorlimbofinternalcapsuleL, awf_RetrolenticularpartofinternalcapsuleR, awf_RetrolenticularpartofinternalcapsuleL, awf_AnteriorcoronaradiataR, awf_AnteriorcoronaradiataL, awf_SuperiorcoronaradiataR, awf_SuperiorcoronaradiataL, awf_PosteriorcoronaradiataR, awf_PosteriorcoronaradiataL, awf_PosteriorthalamicradiationR, awf_PosteriorthalamicradiationL, awf_SagittalstratumR, awf_SagittalstratumL, awf_ExternalcapsuleR, awf_ExternalcapsuleL, awf_CingulumcingulategyrusR, awf_CingulumcingulategyrusL, awf_CingulumhippocampusR, awf_CingulumhippocampusL, awf_Fornix_StriaterminalisR, awf_Fornix_StriaterminalisL, awf_SuperiorlongitudinalfasciculusR, awf_SuperiorlongitudinalfasciculusL, awf_SuperiorfrontooccipitalfasciculusR, awf_SuperiorfrontooccipitalfasciculusL, awf_UncinatefasciculusR, awf_UncinatefasciculusL, awf_TapetumR, awf_TapetumL, awf_Mean, awf_SD, awf_CINGL, awf_CINGR, radEAD_ATRL, radEAD_ATRR, radEAD_CSTL, radEAD_CSTR, radEAD_CGL, radEAD_CGR, radEAD_FMAJ, radEAD_FMIN, radEAD_IFOFL, radEAD_IFOFR, radEAD_ILFL, radEAD_ILFR, radEAD_SLFL, radEAD_SLFR, radEAD_UFL, radEAD_UFR, radEAD_SLFTL, radEAD_SLTFR, radEAD_Middlecerebellarpeduncle, radEAD_Pontine, radEAD_GenuCC, radEAD_BodyCC, radEAD_SpleniumCC, radEAD_Fornix, radEAD_CorticospinaltractR, radEAD_CorticospinaltractL, radEAD_MediallemniscusR, radEAD_MediallemniscusL, radEAD_InferiorcerebellarpeduncleR, radEAD_InferiorcerebellarpeduncleL, radEAD_SuperiorcerebellarpeduncleR, radEAD_SuperiorcerebellarpeduncleL, radEAD_CerebralpeduncleR, radEAD_CerebralpeduncleL, radEAD_AnteriorlimbofinternalcapsuleR, radEAD_AnteriorlimbofinternalcapsuleL, radEAD_ PosteriorlimbofinternalcapsuleR, radEAD_PosteriorlimbofinternalcapsuleL, radEAD_RetrolenticularpartofinternalcapsuleR, radEAD_RetrolenticularpartofinternalcapsuleL, radEAD_AnteriorcoronaradiataR, radEAD_AnteriorcoronaradiataL, radEAD_SuperiorcoronaradiataR, radEAD_SuperiorcoronaradiataL, radEAD_PosteriorcoronaradiataR, radEAD_PosteriorcoronaradiataL, radEAD_PosteriorthalamicradiationR, radEAD_PosteriorthalamicradiationL, radEAD_SagittalstratumR, radEAD_SagittalstratumL, radEAD_ExternalcapsuleR, radEAD_ExternalcapsuleL, radEAD_CingulumcingulategyrusR, radEAD_CingulumcingulategyrusL, radEAD_CingulumhippocampusR, radEAD_CingulumhippocampusL, radEAD_Fornix_StriaterminalisR, radEAD_Fornix_StriaterminalisL, radEAD_SuperiorlongitudinalfasciculusR, radEAD_SuperiorlongitudinalfasciculusL, radEAD_SuperiorfrontooccipitalfasciculusR, radEAD_SuperiorfrontooccipitalfasciculusL, radEAD_UncinatefasciculusR, radEAD_UncinatefasciculusL, radEAD_TapetumR, radEAD_TapetumL, radEAD_Mean, radEAD_SD, radEAD_CINGL, radEAD_CINGR, rk_ATRL, rk_ATRR, rk_CSTL, rk_CSTR, rk_CGL, rk_CGR, rk_FMAJ, rk_FMIN, rk_IFOFL, rk_IFOFR, rk_ILFL, rk_ILFR, rk_SLFL, rk_SLFR, rk_UFL, rk_UFR, rk_SLFTL, rk_SLTFR, rk_Middlecerebellarpeduncle, rk_Pontine, rk_GenuCC, rk_BodyCC, rk_SpleniumCC, rk_Fornix, rk_CorticospinaltractR, rk_CorticospinaltractL, rk_MediallemniscusR, rk_MediallemniscusL, rk_InferiorcerebellarpeduncleR, rk_InferiorcerebellarpeduncleL, rk_SuperiorcerebellarpeduncleR, rk_SuperiorcerebellarpeduncleL, rk_CerebralpeduncleR, rk_CerebralpeduncleL, rk_AnteriorlimbofinternalcapsuleR, rk_AnteriorlimbofinternalcapsuleL, rk_ PosteriorlimbofinternalcapsuleR, rk_PosteriorlimbofinternalcapsuleL, rk_RetrolenticularpartofinternalcapsuleR, rk_RetrolenticularpartofinternalcapsuleL, rk_AnteriorcoronaradiataR, rk_AnteriorcoronaradiataL, rk_SuperiorcoronaradiataR, rk_SuperiorcoronaradiataL, rk_PosteriorcoronaradiataR, rk_PosteriorcoronaradiataL, rk_PosteriorthalamicradiationR, rk_PosteriorthalamicradiationL, rk_SagittalstratumR, rk_SagittalstratumL, rk_ExternalcapsuleR, rk_ExternalcapsuleL, rk_CingulumcingulategyrusR, rk_CingulumcingulategyrusL, rk_CingulumhippocampusR, rk_CingulumhippocampusL, rk_Fornix_StriaterminalisR, rk_Fornix_StriaterminalisL, rk_SuperiorlongitudinalfasciculusR, rk_SuperiorlongitudinalfasciculusL, rk_SuperiorfrontooccipitalfasciculusR, rk_SuperiorfrontooccipitalfasciculusL, rk_UncinatefasciculusR, rk_UncinatefasciculusL, rk_TapetumR, rk_TapetumL, rk_Mean, rk_SD, rk_CINGL, rk_CINGR, ak_ATRL, ak_ATRR, ak_CSTL, ak_CSTR, ak_CGL, ak_CGR, ak_FMAJ, ak_FMIN, ak_IFOFL, ak_IFOFR, ak_ILFL, ak_ILFR, ak_SLFL, ak_SLFR, ak_UFL, ak_UFR, ak_SLFTL, ak_SLTFR, ak_Middlecerebellarpeduncle, ak_Pontine, ak_GenuCC, ak_BodyCC, ak_SpleniumCC, ak_Fornix, ak_CorticospinaltractR, ak_CorticospinaltractL, ak_MediallemniscusR, ak_MediallemniscusL, ak_InferiorcerebellarpeduncleR, ak_InferiorcerebellarpeduncleL, ak_SuperiorcerebellarpeduncleR, ak_SuperiorcerebellarpeduncleL, ak_CerebralpeduncleR, ak_CerebralpeduncleL, ak_AnteriorlimbofinternalcapsuleR, ak_AnteriorlimbofinternalcapsuleL, ak_ PosteriorlimbofinternalcapsuleR, ak_PosteriorlimbofinternalcapsuleL, ak_RetrolenticularpartofinternalcapsuleR, ak_RetrolenticularpartofinternalcapsuleL, ak_AnteriorcoronaradiataR, ak_AnteriorcoronaradiataL, ak_SuperiorcoronaradiataR, ak_SuperiorcoronaradiataL, ak_PosteriorcoronaradiataR, ak_PosteriorcoronaradiataL, ak_PosteriorthalamicradiationR, ak_PosteriorthalamicradiationL, ak_SagittalstratumR, ak_SagittalstratumL, ak_ExternalcapsuleR, ak_ExternalcapsuleL, ak_CingulumcingulategyrusR, ak_CingulumcingulategyrusL, ak_CingulumhippocampusR, ak_CingulumhippocampusL, ak_Fornix_StriaterminalisR, ak_Fornix_StriaterminalisL, ak_SuperiorlongitudinalfasciculusR, ak_SuperiorlongitudinalfasciculusL, ak_SuperiorfrontooccipitalfasciculusR, ak_SuperiorfrontooccipitalfasciculusL, ak_UncinatefasciculusR, ak_UncinatefasciculusL, ak_TapetumR, ak_TapetumL, ak_Mean, ak_SD, ak_CINGL, ak_CINGR, mk_ATRL, mk_ATRR, mk_CSTL, mk_CSTR, mk_CGL, mk_CGR, mk_FMAJ, mk_FMIN, mk_IFOFL, mk_IFOFR, mk_ILFL, mk_ILFR, mk_SLFL, mk_SLFR, mk_UFL, mk_UFR, mk_SLFTL, mk_SLTFR, mk_Middlecerebellarpeduncle, mk_Pontine, mk_GenuCC, mk_BodyCC, mk_SpleniumCC, mk_Fornix, mk_CorticospinaltractR, mk_CorticospinaltractL, mk_MediallemniscusR, mk_MediallemniscusL, mk_InferiorcerebellarpeduncleR, mk_InferiorcerebellarpeduncleL, mk_SuperiorcerebellarpeduncleR, mk_SuperiorcerebellarpeduncleL, mk_CerebralpeduncleR, mk_CerebralpeduncleL, mk_AnteriorlimbofinternalcapsuleR, mk_AnteriorlimbofinternalcapsuleL, mk_ PosteriorlimbofinternalcapsuleR, mk_PosteriorlimbofinternalcapsuleL, mk_RetrolenticularpartofinternalcapsuleR, mk_RetrolenticularpartofinternalcapsuleL, mk_AnteriorcoronaradiataR, mk_AnteriorcoronaradiataL, mk_SuperiorcoronaradiataR, mk_SuperiorcoronaradiataL, mk_PosteriorcoronaradiataR, mk_PosteriorcoronaradiataL, mk_PosteriorthalamicradiationR, mk_PosteriorthalamicradiationL, mk_SagittalstratumR, mk_SagittalstratumL, mk_ExternalcapsuleR, mk_ExternalcapsuleL, mk_CingulumcingulategyrusR, mk_CingulumcingulategyrusL, mk_CingulumhippocampusR, mk_CingulumhippocampusL, mk_Fornix_StriaterminalisR, mk_Fornix_StriaterminalisL, mk_SuperiorlongitudinalfasciculusR, mk_SuperiorlongitudinalfasciculusL, mk_SuperiorfrontooccipitalfasciculusR, mk_SuperiorfrontooccipitalfasciculusL, mk_UncinatefasciculusR, mk_UncinatefasciculusL, mk_TapetumR, mk_TapetumL, mk_Mean, mk_SD, mk_CINGL, mk_CINGR, smt_mc_intra_ATRL, smt_mc_intra_ATRR, smt_mc_intra_CSTL, smt_mc_intra_CSTR, smt_mc_intra_CGL, smt_mc_intra_CGR, smt_mc_intra_FMAJ, smt_mc_intra_FMIN, smt_mc_intra_IFOFL, smt_mc_intra_IFOFR, smt_mc_intra_ILFL, smt_mc_intra_ILFR, smt_mc_intra_SLFL, smt_mc_intra_SLFR, smt_mc_intra_UFL, smt_mc_intra_UFR, smt_mc_intra_SLFTL, smt_mc_intra_SLTFR, smt_mc_intra_Middlecerebellarpeduncle, smt_mc_intra_Pontine, smt_mc_intra_GenuCC, smt_mc_intra_BodyCC, smt_mc_intra_SpleniumCC, smt_mc_intra_Fornix, smt_mc_intra_CorticospinaltractR, smt_mc_intra_CorticospinaltractL, smt_mc_intra_MediallemniscusR, smt_mc_intra_MediallemniscusL, smt_mc_intra_InferiorcerebellarpeduncleR, smt_mc_intra_InferiorcerebellarpeduncleL, smt_mc_intra_SuperiorcerebellarpeduncleR, smt_mc_intra_SuperiorcerebellarpeduncleL, smt_mc_intra_CerebralpeduncleR, smt_mc_intra_CerebralpeduncleL, smt_mc_intra_AnteriorlimbofinternalcapsuleR, smt_mc_intra_AnteriorlimbofinternalcapsuleL, smt_mc_intra_ PosteriorlimbofinternalcapsuleR, smt_mc_intra_PosteriorlimbofinternalcapsuleL, smt_mc_intra_RetrolenticularpartofinternalcapsuleR, smt_mc_intra_RetrolenticularpartofinternalcapsuleL, smt_mc_intra_AnteriorcoronaradiataR, smt_mc_intra_AnteriorcoronaradiataL, smt_mc_intra_SuperiorcoronaradiataR, smt_mc_intra_SuperiorcoronaradiataL, smt_mc_intra_PosteriorcoronaradiataR, smt_mc_intra_PosteriorcoronaradiataL, smt_mc_intra_PosteriorthalamicradiationR, smt_mc_intra_PosteriorthalamicradiationL, smt_mc_intra_SagittalstratumR, smt_mc_intra_SagittalstratumL, smt_mc_intra_ExternalcapsuleR, smt_mc_intra_ExternalcapsuleL, smt_mc_intra_CingulumcingulategyrusR, smt_mc_intra_CingulumcingulategyrusL, smt_mc_intra_CingulumhippocampusR, smt_mc_intra_CingulumhippocampusL, smt_mc_intra_Fornix_StriaterminalisR, smt_mc_intra_Fornix_StriaterminalisL, smt_mc_intra_SuperiorlongitudinalfasciculusR, smt_mc_intra_SuperiorlongitudinalfasciculusL, smt_mc_intra_SuperiorfrontooccipitalfasciculusR, smt_mc_intra_SuperiorfrontooccipitalfasciculusL, smt_mc_intra_UncinatefasciculusR, smt_mc_intra_UncinatefasciculusL, smt_mc_intra_TapetumR, smt_mc_intra_TapetumL, smt_mc_intra_Mean, smt_mc_intra_SD, smt_mc_intra_CINGL, smt_mc_intra_CINGR, smt_mc_extramd_ATRL, smt_mc_extramd_ATRR, smt_mc_extramd_CSTL, smt_mc_extramd_CSTR, smt_mc_extramd_CGL, smt_mc_extramd_CGR, smt_mc_extramd_FMAJ, smt_mc_extramd_FMIN, smt_mc_extramd_IFOFL, smt_mc_extramd_IFOFR, smt_mc_extramd_ILFL, smt_mc_extramd_ILFR, smt_mc_extramd_SLFL, smt_mc_extramd_SLFR, smt_mc_extramd_UFL, smt_mc_extramd_UFR, smt_mc_extramd_SLFTL, smt_mc_extramd_SLTFR, smt_mc_extramd_Middlecerebellarpeduncle, smt_mc_extramd_Pontine, smt_mc_extramd_GenuCC, smt_mc_extramd_BodyCC, smt_mc_extramd_SpleniumCC, smt_mc_extramd_Fornix, smt_mc_extramd_CorticospinaltractR, smt_mc_extramd_CorticospinaltractL, smt_mc_extramd_MediallemniscusR, smt_mc_extramd_MediallemniscusL, smt_mc_extramd_InferiorcerebellarpeduncleR, smt_mc_extramd_InferiorcerebellarpeduncleL, smt_mc_extramd_SuperiorcerebellarpeduncleR, smt_mc_extramd_SuperiorcerebellarpeduncleL, smt_mc_extramd_CerebralpeduncleR, smt_mc_extramd_CerebralpeduncleL, smt_mc_extramd_AnteriorlimbofinternalcapsuleR, smt_mc_extramd_AnteriorlimbofinternalcapsuleL, smt_mc_extramd_ PosteriorlimbofinternalcapsuleR, smt_mc_extramd_PosteriorlimbofinternalcapsuleL, smt_mc_extramd_RetrolenticularpartofinternalcapsuleR, smt_mc_extramd_RetrolenticularpartofinternalcapsuleL, smt_mc_extramd_AnteriorcoronaradiataR, smt_mc_extramd_AnteriorcoronaradiataL, smt_mc_extramd_SuperiorcoronaradiataR, smt_mc_extramd_SuperiorcoronaradiataL, smt_mc_extramd_PosteriorcoronaradiataR, smt_mc_extramd_PosteriorcoronaradiataL, smt_mc_extramd_PosteriorthalamicradiationR, smt_mc_extramd_PosteriorthalamicradiationL, smt_mc_extramd_SagittalstratumR, smt_mc_extramd_SagittalstratumL, smt_mc_extramd_ExternalcapsuleR, smt_mc_extramd_ExternalcapsuleL, smt_mc_extramd_CingulumcingulategyrusR, smt_mc_extramd_CingulumcingulategyrusL, smt_mc_extramd_CingulumhippocampusR, smt_mc_extramd_CingulumhippocampusL, smt_mc_extramd_Fornix_StriaterminalisR, smt_mc_extramd_Fornix_StriaterminalisL, smt_mc_extramd_SuperiorlongitudinalfasciculusR, smt_mc_extramd_SuperiorlongitudinalfasciculusL, smt_mc_extramd_SuperiorfrontooccipitalfasciculusR, smt_mc_extramd_SuperiorfrontooccipitalfasciculusL, smt_mc_extramd_UncinatefasciculusR, smt_mc_extramd_UncinatefasciculusL, smt_mc_extramd_TapetumR, smt_mc_extramd_TapetumL, smt_mc_extramd_Mean, smt_mc_extramd_SD, smt_mc_extramd_CINGL, smt_mc_extramd_CINGR, smt_mc_extratrans_ATRL, smt_mc_extratrans_ATRR, smt_mc_extratrans_CSTL, smt_mc_extratrans_CSTR, smt_mc_extratrans_CGL, smt_mc_extratrans_CGR, smt_mc_extratrans_FMAJ, smt_mc_extratrans_FMIN, smt_mc_extratrans_IFOFL, smt_mc_extratrans_IFOFR, smt_mc_extratrans_ILFL, smt_mc_extratrans_ILFR, smt_mc_extratrans_SLFL, smt_mc_extratrans_SLFR, smt_mc_extratrans_UFL, smt_mc_extratrans_UFR, smt_mc_extratrans_SLFTL, smt_mc_extratrans_SLTFR, smt_mc_extratrans_Middlecerebellarpeduncle, smt_mc_extratrans_Pontine, smt_mc_extratrans_GenuCC, smt_mc_extratrans_BodyCC, smt_mc_extratrans_SpleniumCC, smt_mc_extratrans_Fornix, smt_mc_extratrans_CorticospinaltractR, smt_mc_extratrans_CorticospinaltractL, smt_mc_extratrans_MediallemniscusR, smt_mc_extratrans_MediallemniscusL, smt_mc_extratrans_InferiorcerebellarpeduncleR, smt_mc_extratrans_InferiorcerebellarpeduncleL, smt_mc_extratrans_SuperiorcerebellarpeduncleR, smt_mc_extratrans_SuperiorcerebellarpeduncleL, smt_mc_extratrans_CerebralpeduncleR, smt_mc_extratrans_CerebralpeduncleL, smt_mc_extratrans_AnteriorlimbofinternalcapsuleR, smt_mc_extratrans_AnteriorlimbofinternalcapsuleL, smt_mc_extratrans_ PosteriorlimbofinternalcapsuleR, smt_mc_extratrans_PosteriorlimbofinternalcapsuleL, smt_mc_extratrans_RetrolenticularpartofinternalcapsuleR, smt_mc_extratrans_RetrolenticularpartofinternalcapsuleL, smt_mc_extratrans_AnteriorcoronaradiataR, smt_mc_extratrans_AnteriorcoronaradiataL, smt_mc_extratrans_SuperiorcoronaradiataR, smt_mc_extratrans_SuperiorcoronaradiataL, smt_mc_extratrans_PosteriorcoronaradiataR, smt_mc_extratrans_PosteriorcoronaradiataL, smt_mc_extratrans_PosteriorthalamicradiationR, smt_mc_extratrans_PosteriorthalamicradiationL, smt_mc_extratrans_SagittalstratumR, smt_mc_extratrans_SagittalstratumL, smt_mc_extratrans_ExternalcapsuleR, smt_mc_extratrans_ExternalcapsuleL, smt_mc_extratrans_CingulumcingulategyrusR, smt_mc_extratrans_CingulumcingulategyrusL, smt_mc_extratrans_CingulumhippocampusR, smt_mc_extratrans_CingulumhippocampusL, smt_mc_extratrans_Fornix_StriaterminalisR, smt_mc_extratrans_Fornix_StriaterminalisL, smt_mc_extratrans_SuperiorlongitudinalfasciculusR, smt_mc_extratrans_SuperiorlongitudinalfasciculusL, smt_mc_extratrans_SuperiorfrontooccipitalfasciculusR, smt_mc_extratrans_SuperiorfrontooccipitalfasciculusL, smt_mc_extratrans_UncinatefasciculusR, smt_mc_extratrans_UncinatefasciculusL, smt_mc_extratrans_TapetumR, smt_mc_extratrans_TapetumL, smt_mc_extratrans_Mean, smt_mc_extratrans_SD, smt_mc_extratrans_CINGL, smt_mc_extratrans_CINGR |
| Body | VAT, ASAT, ATLMV_L, ATLMV_R, PTLMV_L, PTLMV_R, Liver_PDFF, abdominal_FR, TAATi, WMR, TAT, TTMV, MFI, ATMFI_L, ATMFI_R, PTMFI_L, PTMFI_R, VATi, ASATi, ATLMV_Li, ATLMV_Ri, PTLMV_Li, PTLMV_Ri, TATi, TTMVi, hand_grip_strength_left-2.0, hand_grip_strength_right-2.0, Body_fat_percentage-2.0, Whole_body_fat_mass-2.0, Whole_body_fatfree_mass-2.0, BMI_BodyComp-2.0, Impedence_whole_body-2.0, Trunk_fat_percentage-2.0, Waist_circumference-2.0, Hip_circumference-2.0, Pulse_rate_automated-2.0, Forced_vital_capacity-2.0, Diastolic_BP_automated-2.0, Systolic_BP_automated-2.0, BMI-2.0 |

| **SI Table 2.** **Bayes Factor (BF).** Showing evidence ratio interpretations. Values of 1 can be interpreted as no evidence in either direction, with the following values indicating weight of evidence towards the alternative hypothesis: 0.3-1 (anecdotal), 0.1-0.3 (moderate), 0.03-0.1 (strong), 0.01- 0.03 (very strong), <0.01 (extreme). Contrarily, the following values indicate weight of evidence towards the null hypothesis: 1-3 (anecdotal), 3-10 (moderate), 10-30 (strong), 30-100 (very strong), >100 (extreme). | | | |
| --- | --- | --- | --- |
| Bayes factor BF_12_ | | | Interpretation |
|  | > | 100 | Extreme evidence for M_1_ |
| 30 | - | 100 | Very strong evidence for M_1_ |
| 10 | - | 30 | Strong evidence for M_1_ |
| 3 | - | 10 | Moderate evidence for M_1_ |
| 1 | - | 3 | Anecdotal evidence for M_1_ |
|  | 1 |  | No evidence |
| 0.3 | - | 1 | Anecdotal evidence for M_2_ |
| 0.10 | - | 0.3 | Moderate evidence for M_2_ |
| 0.03 | - | 0.10 | Strong evidence for M_2_ |
| 0.001 | - | 0.03 | Very strong evidence for M_2_ |
|  | < | 0.001 | Extreme evidence for M_2_ |

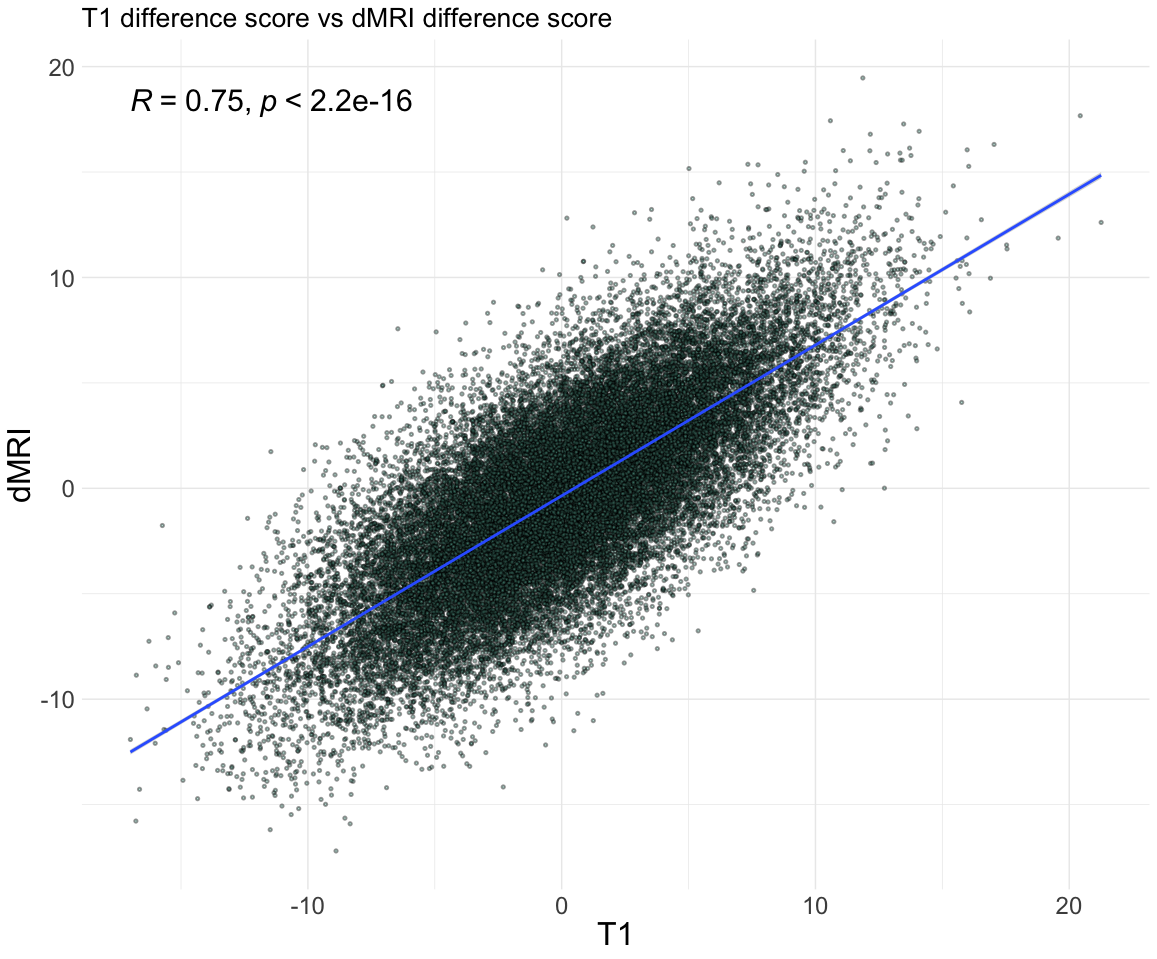
 **SI Figure 5.** Difference score of T1 and dMRI based models (difference scores calculated as predicted brain age *minus* predicted body age). The correlation value of 0.75 indicates that the changes in predictions introduced by health trait are related, but not identical, for T1 and dMRI based models.

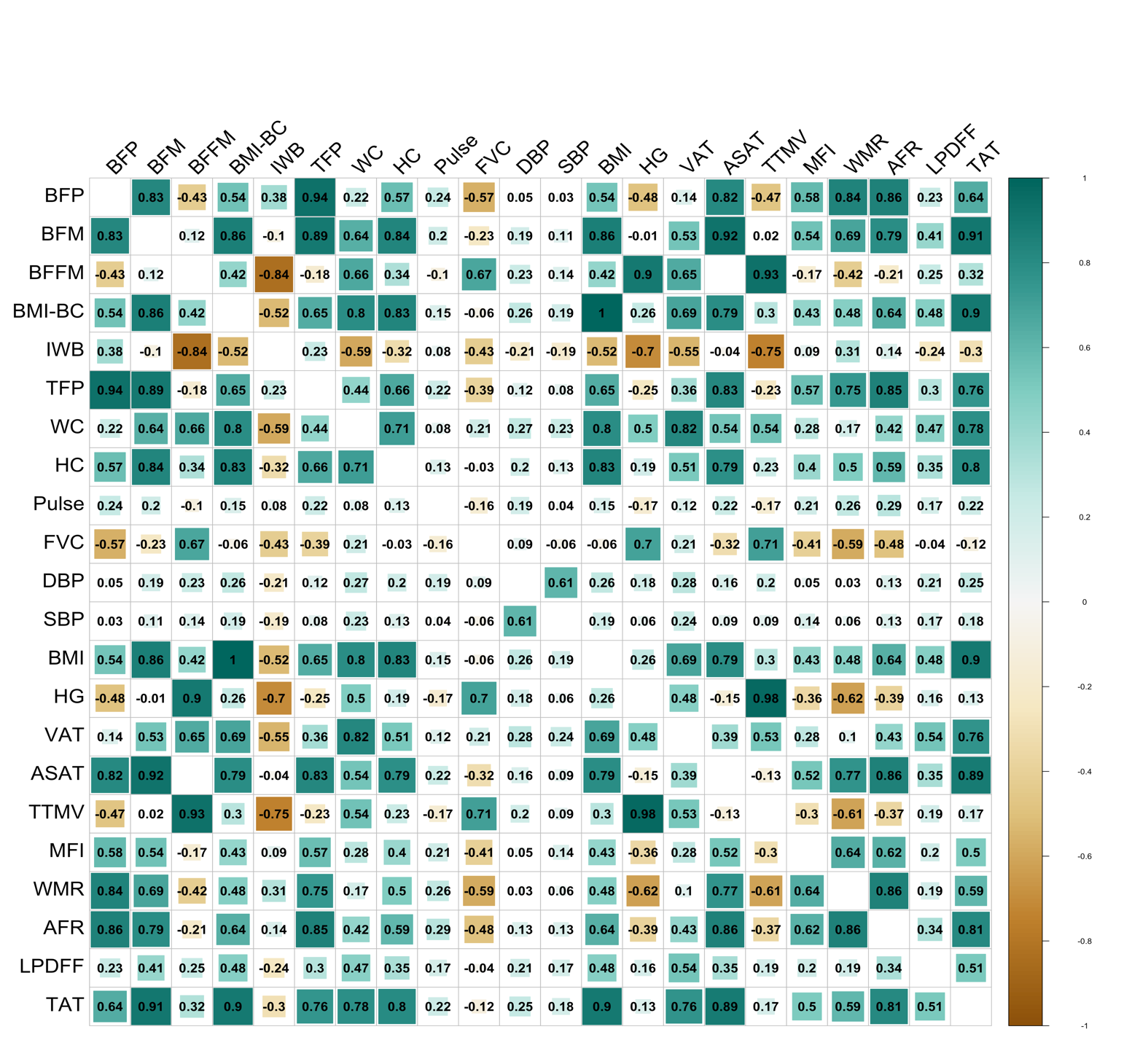

**SI Figure 6.** Correlation matrix showing all bodily health traits used in the study. Abbreviations: body fat percentage (BFP), whole body fat mass (BFM), whole body fat-free mass (BFFM), body-mass index body composition (BMI-BC), impedance of whole body (IWB), trunk fat percentage (TFP), waist circumference (WC), hip circumference (HC), forced vital capacity (FVC), diastolic blood pressure (DBP), systolic blood pressure (SBP), body-mass index (BMI), hand grip strength (HG), visceral adipose tissue (VAT), abdominal subcutaneous adipose tissue (ASAT), total thigh fat-free muscle volume (TTMV), muscle fat infiltration (MFI), weight-to-muscle ratio (WMR), abdominal fat ratio (AFR), liver fat (LPDFF), total abdominal tissue volume (TAT). Full descriptions of variables can be found in SI Section 1.

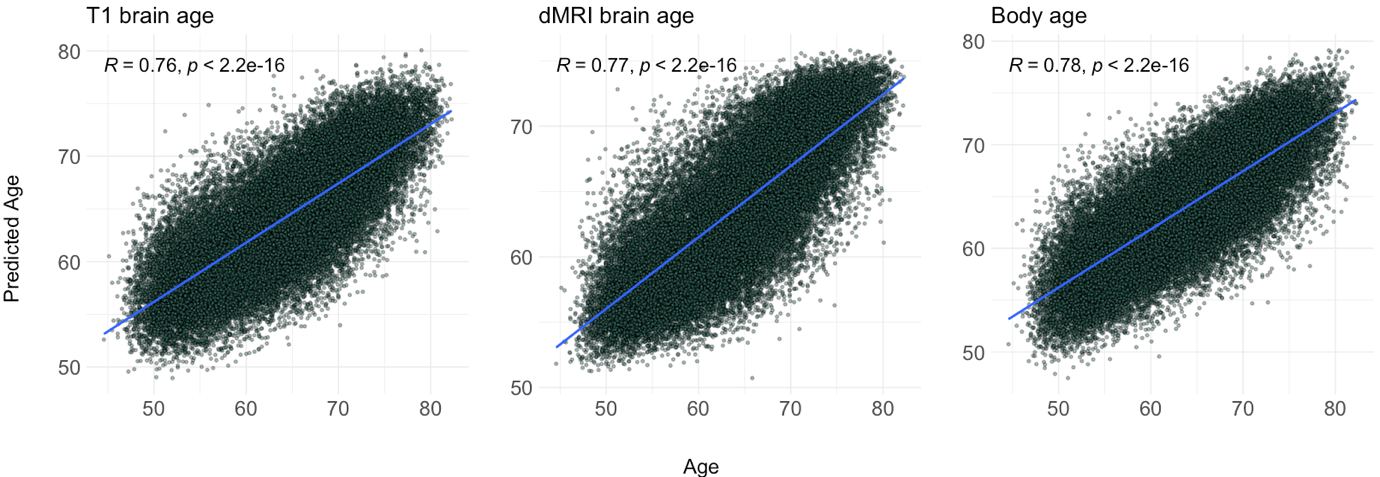

**SI Figure 7.** Predicted age of each age prediction model as a function of chronological age. R (*Pearson’s*) reflects model performance.

| **SI Table 3.** Bayesian multilevel model results for both dMRI and T1-weighted associations with body MRI measures. Table shows results of each body MRI measure for its respective MRI model, with evidence ratios under 1 indicating evidence for the alternative hypothesis and values over 1 indicating evidence for the null hypothesis. Estimate indicates directionality of evidence (positive/negative association). See SI Table 1 for precise interpretation of evidence ratio values. | | | | | | | |
| --- | --- | --- | --- | --- | --- | --- | --- |
| **Model** | **Measure** | **Estimate** | **Lower 95** | **Upper 95** | **P higher** | **P lower** | **Evidence** |
| Diffusion-weighted | VAT | -0.029 | -0.042 | -0.017 | 0 | 1 | 0 |
| Diffusion-weighted | ASAT | 0.067 | 0.056 | 0.079 | 1 | 0 | 0 |
| Diffusion-weighted | ATMV | 0.481 | 0.461 | 0.499 | 1 | 0 | 0 |
| Diffusion-weighted | PTMV | 0.216 | 0.197 | 0.235 | 1 | 0 | 0 |
| Diffusion-weighted | ATMFI | -0.225 | -0.238 | -0.212 | 0 | 1 | 0 |
| Diffusion-weighted | PTMFI | -0.157 | -0.169 | -0.145 | 0 | 1 | 0 |
| Diffusion-weighted | MFI | -0.224 | -0.236 | -0.213 | 0 | 1 | 0 |
| Diffusion-weighted | WMR | -0.108 | -0.122 | -0.095 | 0 | 1 | 0 |
| Diffusion-weighted | AFR | -0.055 | -0.067 | -0.043 | 0 | 1 | 0 |
| Diffusion-weighted | LPDFF | 0.029 | 0.018 | 0.04 | 1 | 0 | 0 |
| Diffusion-weighted | TTMV | 0.334 | 0.315 | 0.354 | 1 | 0 | 0 |
| Diffusion-weighted | TAT | 0.031 | 0.02 | 0.042 | 1 | 0 | 0 |
| T1-weighted | VAT | -0.02 | -0.032 | -0.007 | 0.001 | 0.999 | 0.295 |
| T1-weighted | ASAT | 0.083 | 0.072 | 0.095 | 1 | 0 | 0 |
| T1-weighted | ATMV | 0.462 | 0.443 | 0.481 | 1 | 0 | 0 |
| T1-weighted | PTMV | 0.218 | 0.2 | 0.236 | 1 | 0 | 0 |
| T1-weighted | ATMFI | -0.197 | -0.21 | -0.186 | 0 | 1 | 0 |
| T1-weighted | PTMFI | -0.138 | -0.15 | -0.126 | 0 | 1 | 0 |
| T1-weighted | MFI | -0.197 | -0.209 | -0.185 | 0 | 1 | 0 |
| T1-weighted | WMR | -0.087 | -0.101 | -0.073 | 0 | 1 | 0 |
| T1-weighted | AFR | -0.044 | -0.055 | -0.032 | 0 | 1 | 0 |
| T1-weighted | LPDFF | 0.038 | 0.027 | 0.048 | 1 | 0 | 0 |
| T1-weighted | TTMV | 0.328 | 0.308 | 0.347 | 1 | 0 | 0 |
| T1-weighted | TAT | 0.045 | 0.034 | 0.055 | 1 | 0 | 0 |

| **SI Table 4.** Bayesian multilevel model results for both dMRI and T1-weighted associations with cardiometabolic, anthropometric, and bioimpedance measures. Table shows results of each health trait for its respective MRI model, with evidence ratios under 1 indicating evidence for the alternative hypothesis and values over 1 indicating evidence for the null hypothesis. | | | | | | | |
| --- | --- | --- | --- | --- | --- | --- | --- |
| **Model** | **Measure** | **Estimate** | **Lower 95** | **Upper 95** | **P higher** | **P lower** | **Evidence** |
| Diffusion-weighted | WC | 0.015 | 0.003 | 0.027 | 0.992 | 0.008 | 2.798 |
| Diffusion-weighted | SBP | -0.15 | -0.163 | -0.138 | 0 | 1 | 0 |
| Diffusion-weighted | HC | 0.033 | 0.022 | 0.043 | 1 | 0 | 0 |
| Diffusion-weighted | DBP | 0.089 | 0.077 | 0.101 | 1 | 0 | 0 |
| Diffusion-weighted | BFP | 0.027 | 0.013 | 0.042 | 1 | 0 | 0.047 |
| Diffusion-weighted | IWB | -0.019 | -0.035 | -0.006 | 0.004 | 0.996 | 1.226 |
| Diffusion-weighted | HG | 0.229 | 0.212 | 0.246 | 1 | 0 | 0 |
| Diffusion-weighted | BFM | 0.054 | 0.043 | 0.066 | 1 | 0 | 0 |
| Diffusion-weighted | TFP | 0.012 | 0 | 0.024 | 0.973 | 0.027 | 7.783 |
| Diffusion-weighted | BMI | 0.037 | 0.027 | 0.048 | 1 | 0 | 0 |
| Diffusion-weighted | Pulse | 0.027 | 0.015 | 0.039 | 1 | 0 | 0.006 |
| T1-weighted | WC | 0.029 | 0.017 | 0.042 | 1 | 0 | 0 |
| T1-weighted | SBP | -0.192 | -0.205 | -0.181 | 0 | 1 | 0 |
| T1-weighted | HC | 0.046 | 0.035 | 0.057 | 1 | 0 | 0 |
| T1-weighted | DBP | 0.04 | 0.027 | 0.051 | 1 | 0 | 0 |
| T1-weighted | BFP | 0.042 | 0.027 | 0.056 | 1 | 0 | 0 |
| T1-weighted | IWB | -0.038 | -0.053 | -0.024 | 0 | 1 | 0 |
| T1-weighted | HG | 0.194 | 0.178 | 0.211 | 1 | 0 | 0 |
| T1-weighted | BFM | 0.069 | 0.057 | 0.08 | 1 | 0 | 0 |
| T1-weighted | TFP | 0.022 | 0.01 | 0.034 | 1 | 0 | 0.015 |
| T1-weighted | BMI | 0.055 | 0.043 | 0.066 | 1 | 0 | 0 |
| T1-weighted | Pulse | 0.019 | 0.006 | 0.031 | 0.999 | 0.001 | 0.455 |

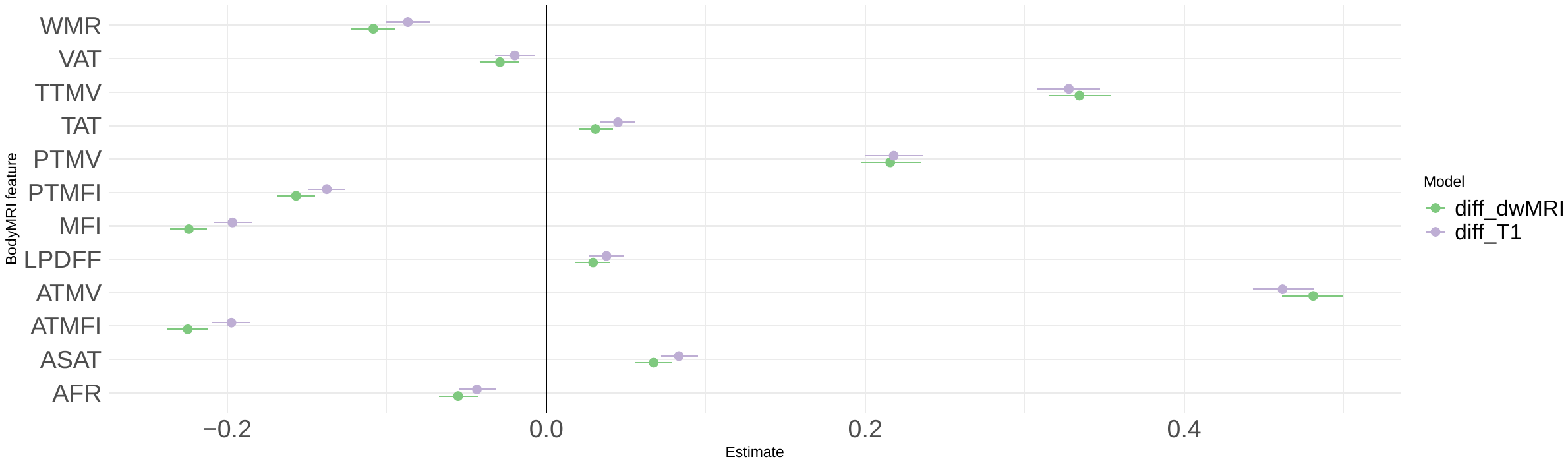

**SI Figure 8. Estimate credible intervals for body MRI.** Figure shows estimates with 95% credible intervals. T1-weighted associations are represented by green points, and dMRI-weighted associations by lilac points.

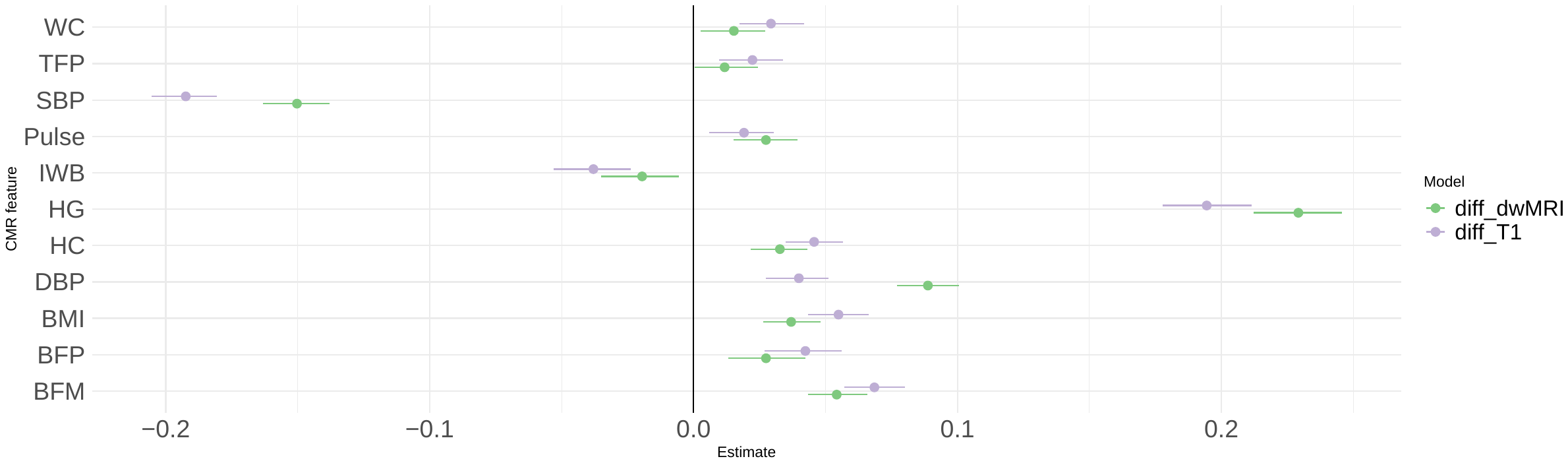

**SI Figure 9. Estimate credible intervals for cardiometabolic, anthropometric, and bioimpedance health traits.** Figure shows estimates with 95% credible intervals. T1-weighted associations are represented by lilac points, and dMRI-weighted associations by green points.

**SI Table 5.** Full (left) linear regressions (Reg) and partial linear regressions (P Reg) for age and each health trait, with the partial regressions being adjusted for T1- (middle) and dwMRI- (right) -weighted predicted age.

| **Health trait** | **Reg** | **Std Error** | **P value** | **P Reg T1** | **Std Error** | **P value** | **P Reg dwMRI** | **Std Error** | **P value** |
| --- | --- | --- | --- | --- | --- | --- | --- | --- | --- |
| **VAT** | 0.35 | 0.019 | 0 | 0.001 | 0.013 | 0.946 | -0.082 | 0.012 | 0 |
| **ASAT** | -0.193 | 0.013 | 0 | -0.095 | 0.009 | 0 | -0.028 | 0.009 | 0.001 |
| **LPDFF** | 0.02 | 0.011 | 0.08 | -0.065 | 0.007 | 0 | -0.07 | 0.007 | 0 |
| **AFR** | 5.411 | 0.37 | 0 | 1.894 | 0.242 | 0 | 3.486 | 0.237 | 0 |
| **TAATi** | 0.043 | 0.026 | 0.103 | -0.092 | 0.017 | 0 | -0.008 | 0.017 | 0.63 |
| **WMR** | 0.547 | 0.032 | 0 | 0.226 | 0.021 | 0 | 0.427 | 0.02 | 0 |
| **TAT** | -0.01 | 0.009 | 0.302 | -0.047 | 0.006 | 0 | -0.032 | 0.006 | 0 |
| **TTMV** | -0.382 | 0.016 | 0 | -0.221 | 0.011 | 0 | -0.332 | 0.01 | 0 |
| **MFI** | 1.218 | 0.023 | 0 | 0.443 | 0.016 | 0 | 0.524 | 0.015 | 0 |
| **VATi** | 1.368 | 0.059 | 0 | 0.103 | 0.04 | 0.009 | -0.092 | 0.039 | 0.02 |
| **ASATi** | -0.327 | 0.035 | 0 | -0.166 | 0.023 | 0 | 0.049 | 0.023 | 0.03 |
| **TATi** | 0.082 | 0.026 | 0.002 | -0.076 | 0.017 | 0 | 0.007 | 0.017 | 0.67 |
| **TTMVi** | -1.628 | 0.07 | 0 | -1.08 | 0.046 | 0 | -1.469 | 0.045 | 0 |
| **BFP** | -0.003 | 0.005 | 0.61 | 0.007 | 0.003 | 0.055 | 0.042 | 0.003 | 0 |
| **BFM** | -0.041 | 0.005 | 0 | -0.028 | 0.003 | 0 | -0.009 | 0.003 | 0.01 |
| **BFFM** | -0.044 | 0.004 | 0 | -0.038 | 0.003 | 0 | -0.063 | 0.002 | 0 |
| **BMI_BC** | -0.053 | 0.01 | 0 | -0.087 | 0.007 | 0 | -0.074 | 0.006 | 0 |
| **IWB** | -0.006 | 0 | 0 | 0.001 | 0 | 0.075 | 0.003 | 0 | 0 |
| **TFP** | 0.024 | 0.006 | 0 | 0.011 | 0.004 | 0.003 | 0.035 | 0.004 | 0 |
| **WC** | 0.048 | 0.003 | 0 | -0.008 | 0.002 | 0 | -0.018 | 0.002 | 0 |
| **HC** | -0.031 | 0.005 | 0 | -0.019 | 0.003 | 0 | -0.01 | 0.003 | 0.002 |
| **Pulse** | 0.016 | 0.004 | 0 | -0.001 | 0.003 | 0.794 | 0 | 0.002 | 0.90 |
| **FVC** | -2.078 | 0.046 | 0 | -0.838 | 0.031 | 0 | -1.119 | 0.03 | 0 |
| **DBP** | -0.024 | 0.004 | 0 | -0.03 | 0.003 | 0 | -0.058 | 0.003 | 0 |
| **SBP** | 0.118 | 0.002 | 0 | 0.043 | 0.002 | 0 | 0.027 | 0.002 | 0 |
| **BMI** | -0.055 | 0.01 | 0 | -0.088 | 0.007 | 0 | -0.076 | 0.006 | 0 |
| **ATMV** | -2.986 | 0.086 | 0 | -1.627 | 0.057 | 0 | -2.198 | 0.055 | 0 |
| **PTMV** | -0.832 | 0.052 | 0 | -0.523 | 0.034 | 0 | -0.867 | 0.033 | 0 |
| **ATMVi** | -13.281 | 0.345 | 0 | -7.735 | 0.228 | 0 | -9.664 | 0.22 | 0 |
| **PTMVi** | -2.861 | 0.225 | 0 | -2.373 | 0.146 | 0 | -3.541 | 0.143 | 0 |
| **ATMFI** | 1.222 | 0.023 | 0 | 0.444 | 0.016 | 0 | 0.526 | 0.015 | 0 |
| **PTMFI** | 0.831 | 0.017 | 0 | 0.264 | 0.012 | 0 | 0.314 | 0.012 | 0 |
| **HG** | -0.12 | 0.004 | 0 | -0.054 | 0.003 | 0 | -0.084 | 0.003 | 0 |

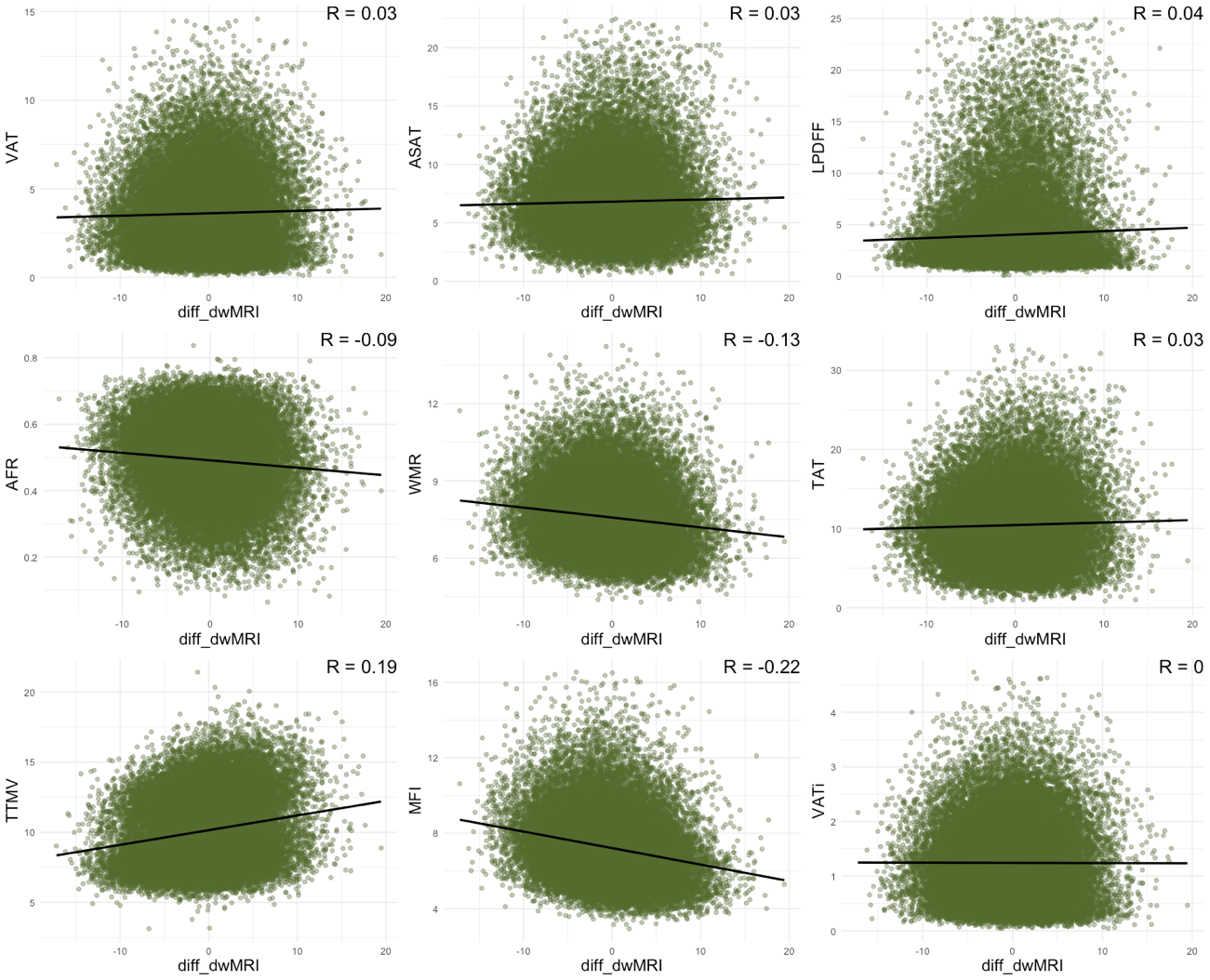

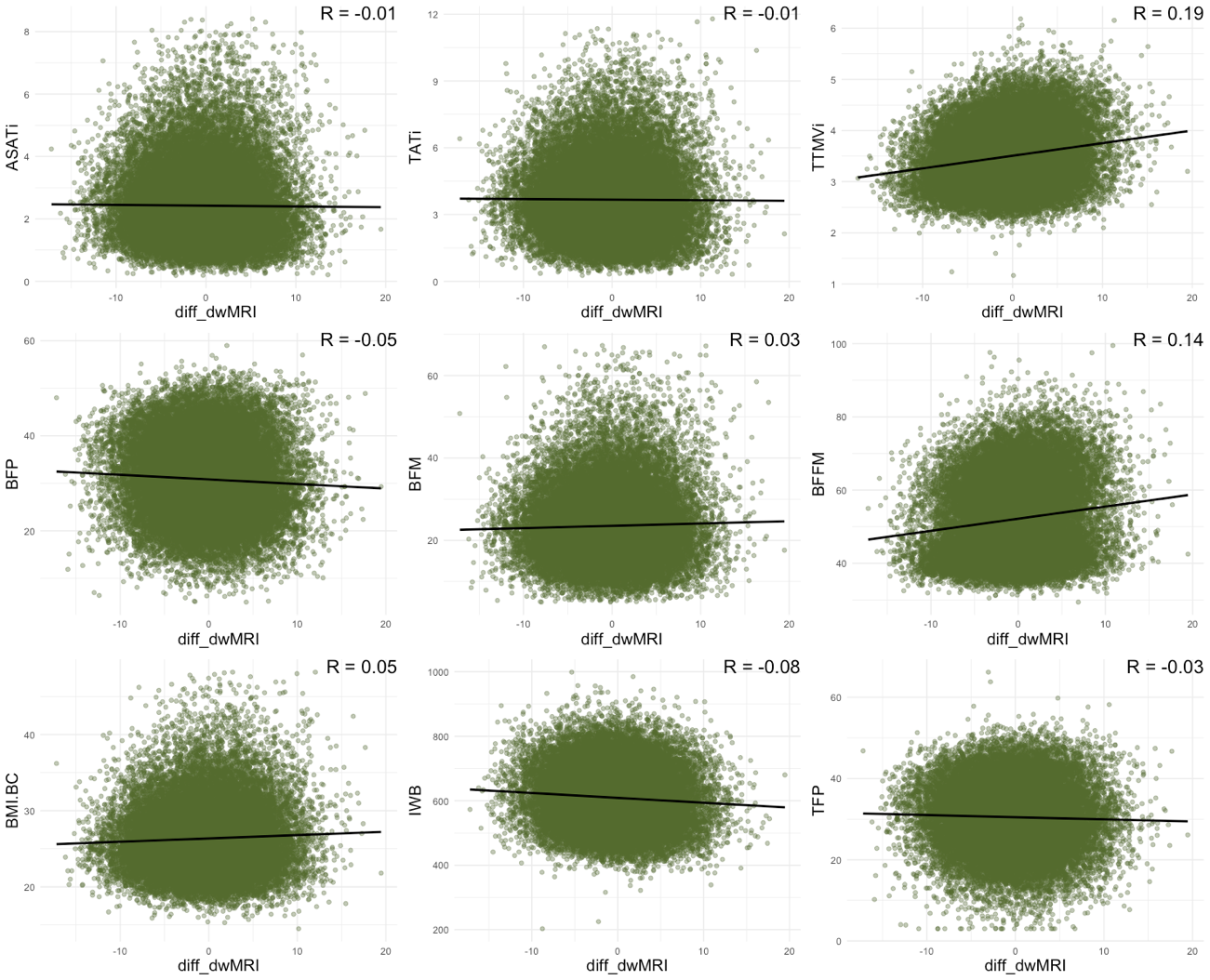

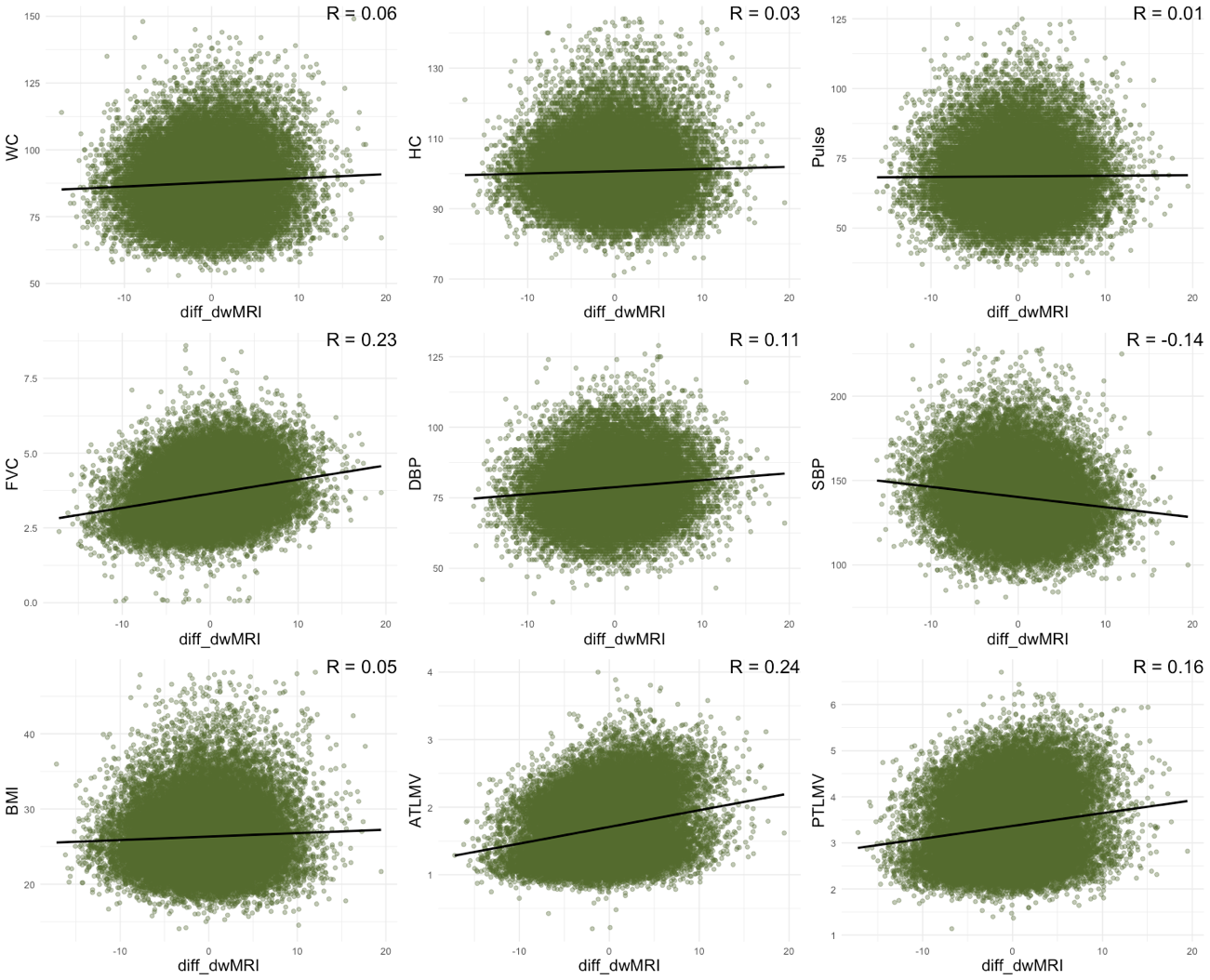

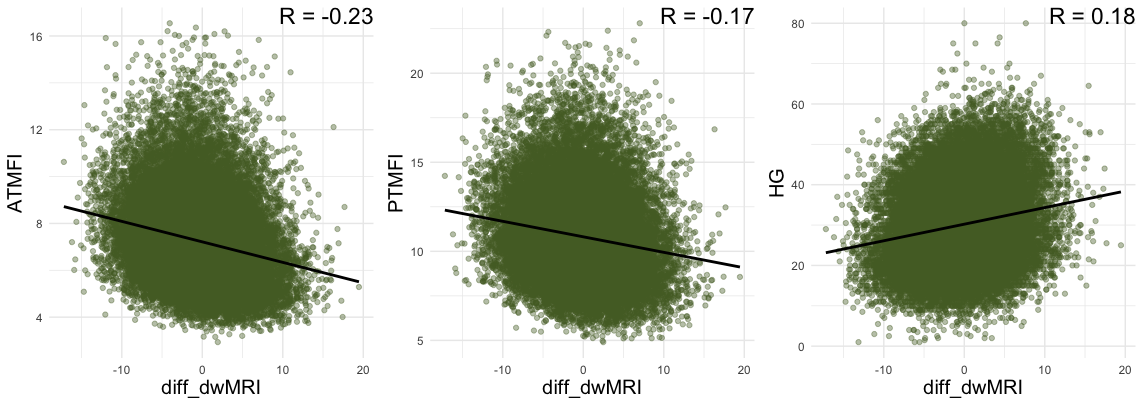

**SI Figure 10.** Showing scatterplots and Pearson’s R values for associations between health traits and difference scores between brain age and body age models for the dMRI models.

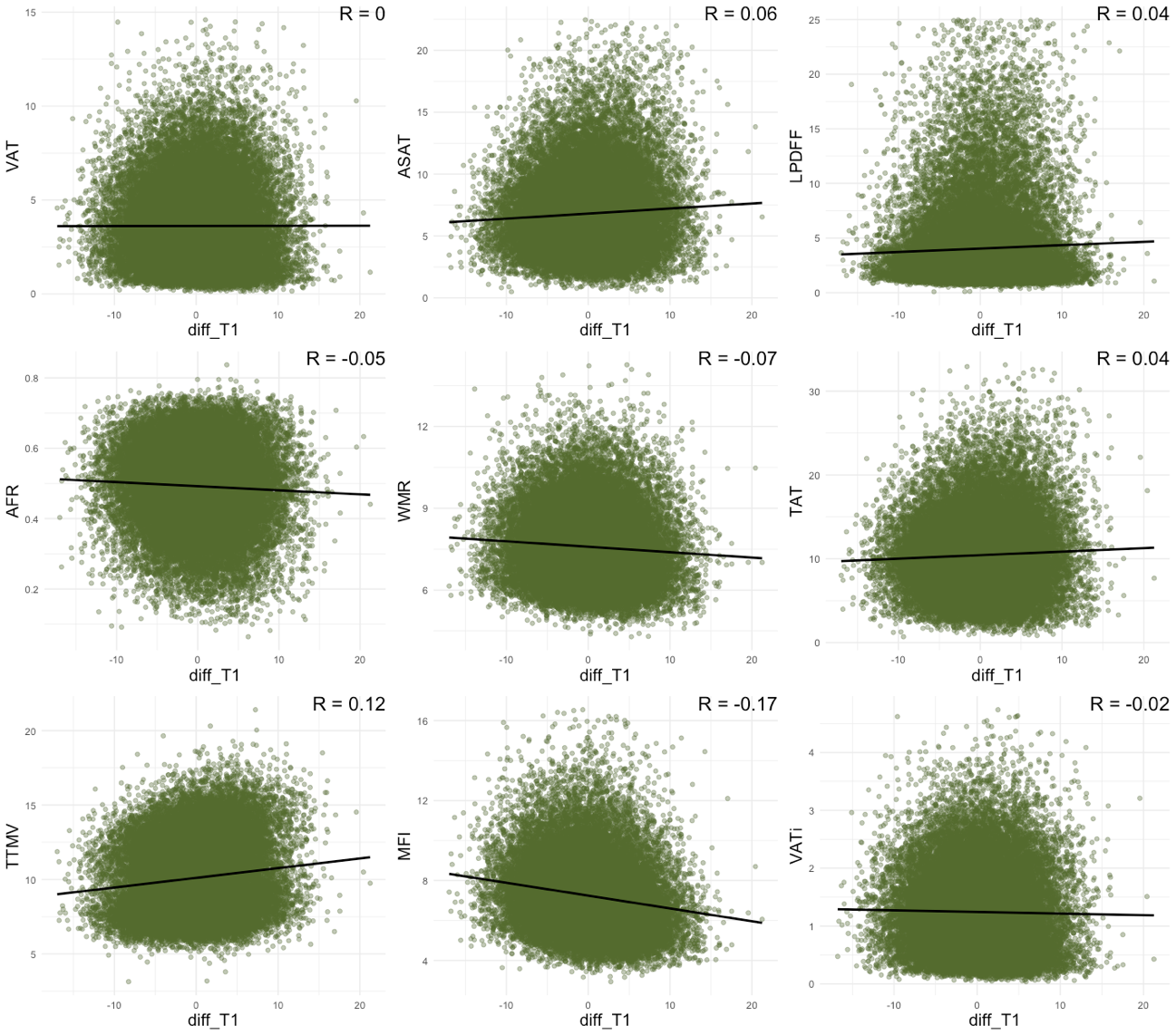

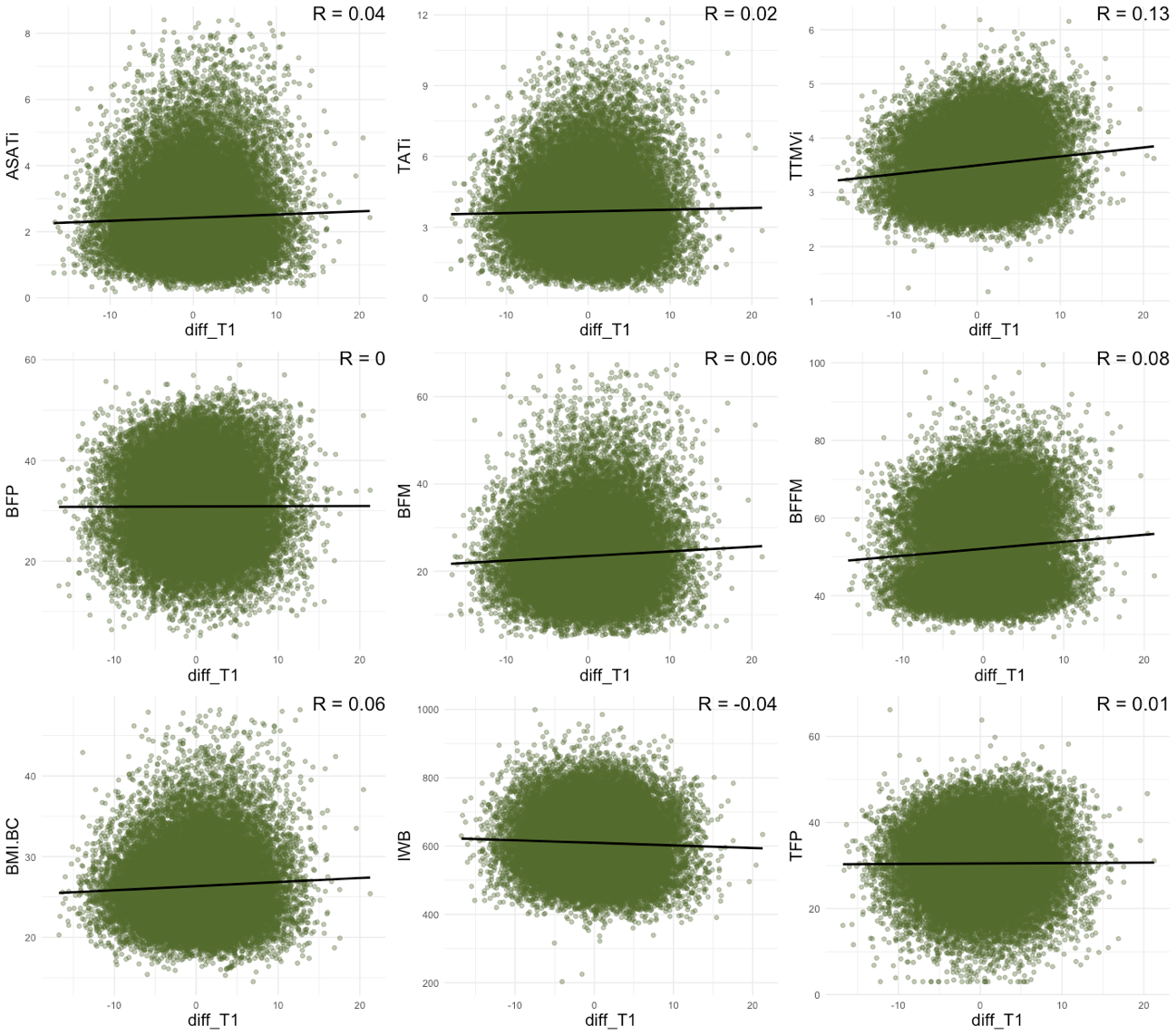

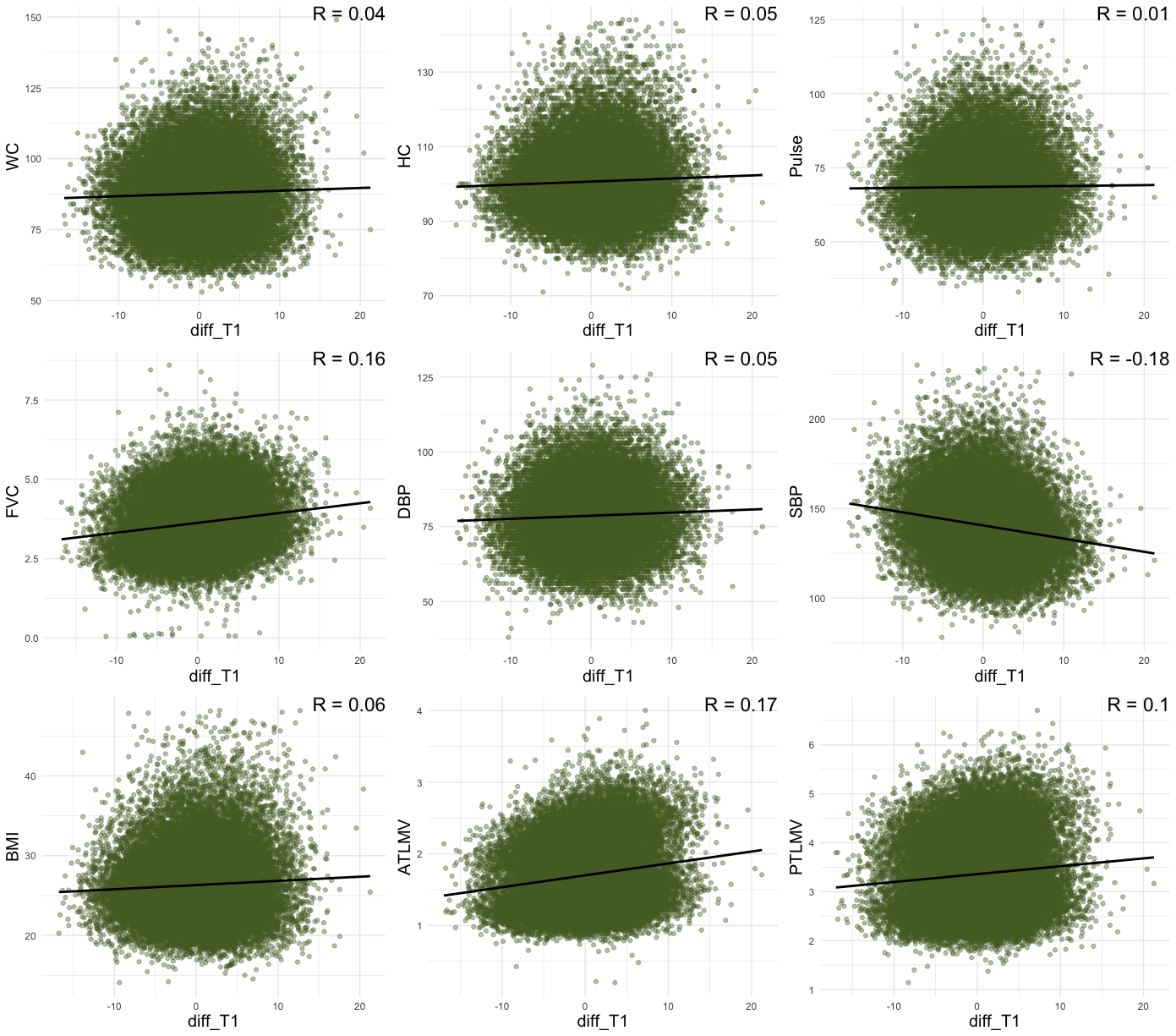

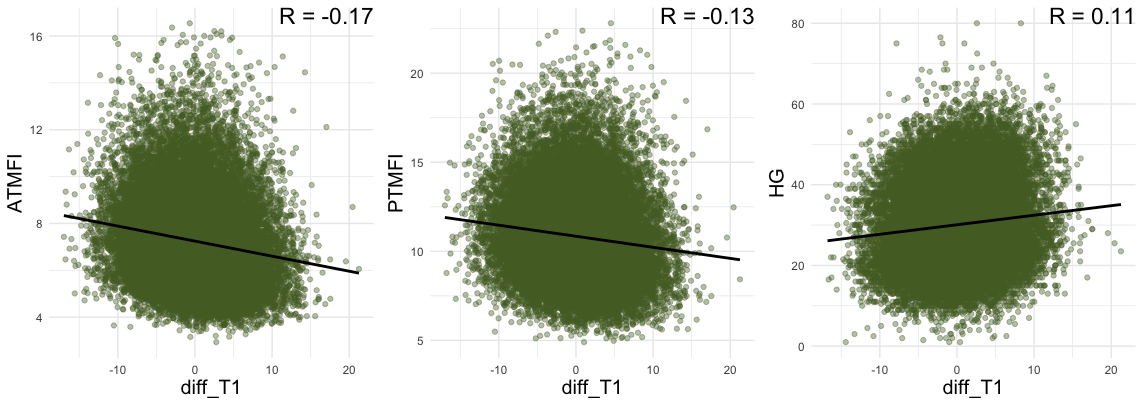

**SI Figure 11.** Showing scatterplots and Pearson’s R values for associations between health traits and difference scores between brain age and body age models for the T1-weighted models.

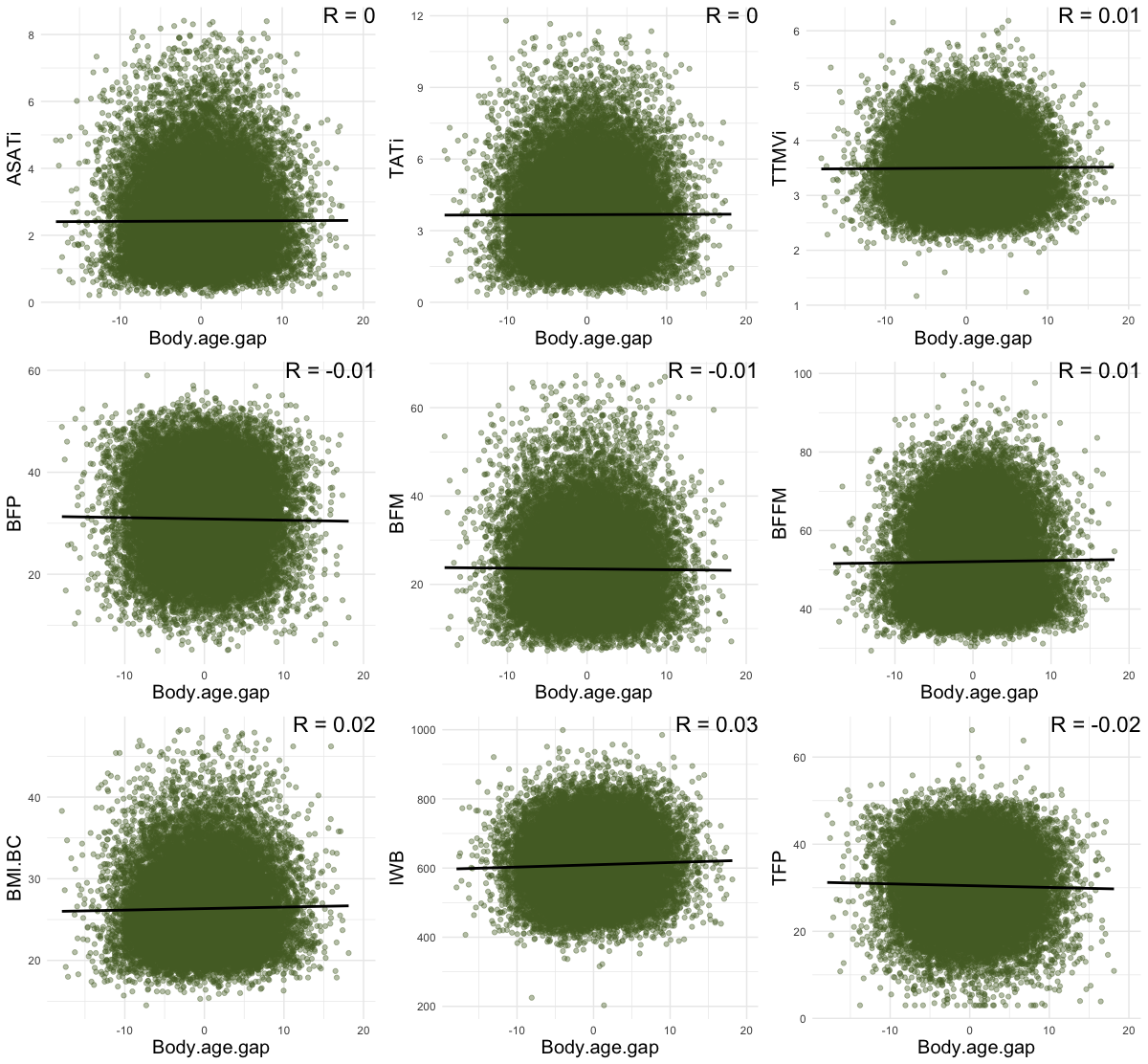

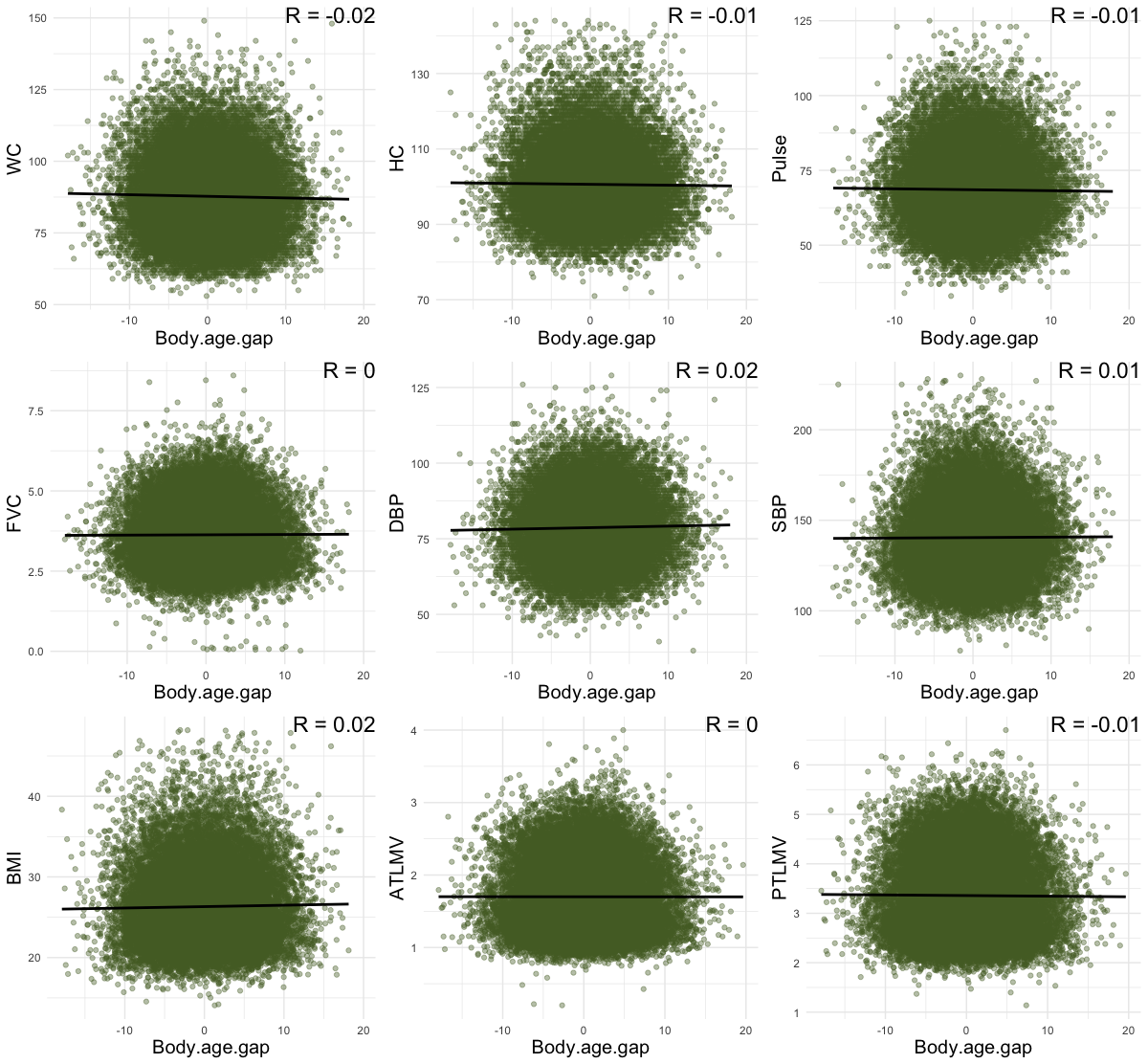

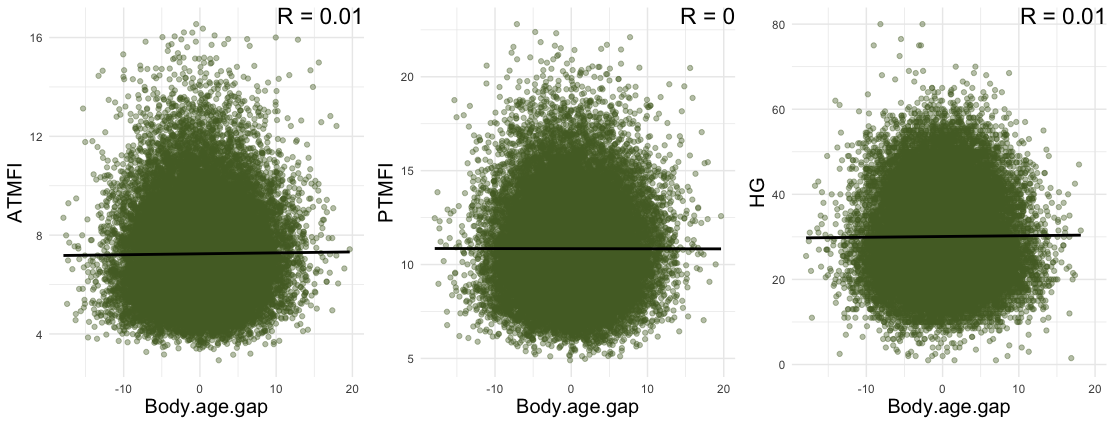

**SI Figure 12.** Showing scatterplots and Pearson’s R values for associations between health traits and body age gap.

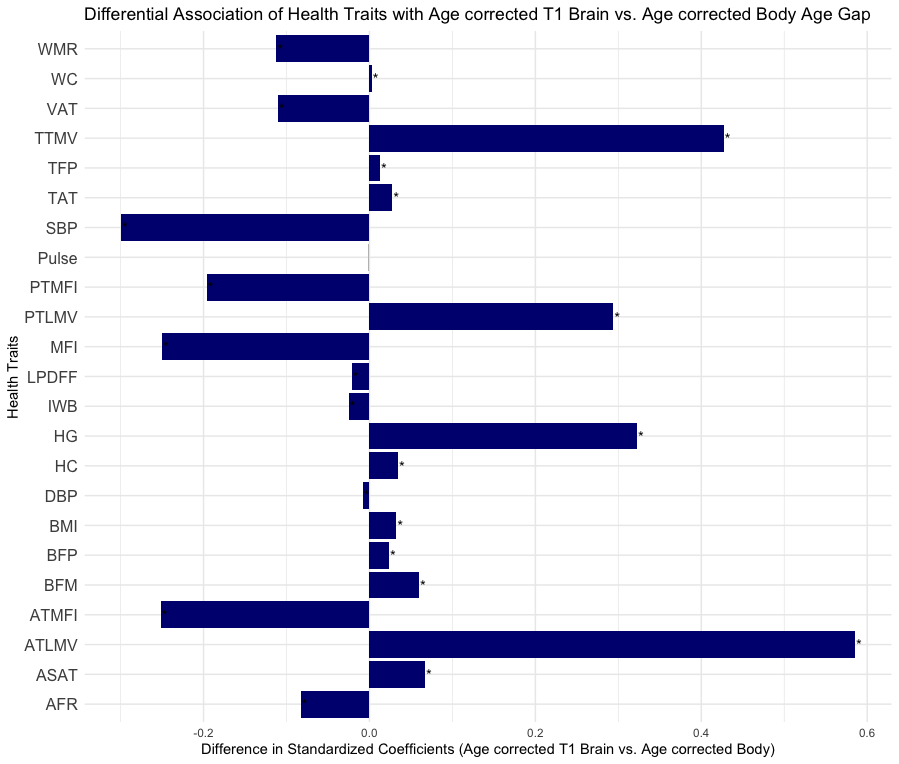

**SI Figure 13**. Results for a linear mixed effects model formula including age-bias-corrected-difference-score ~ health trait + Sex (|ID). The age-bias correction used is described in detail in de Lange et al. (2022).

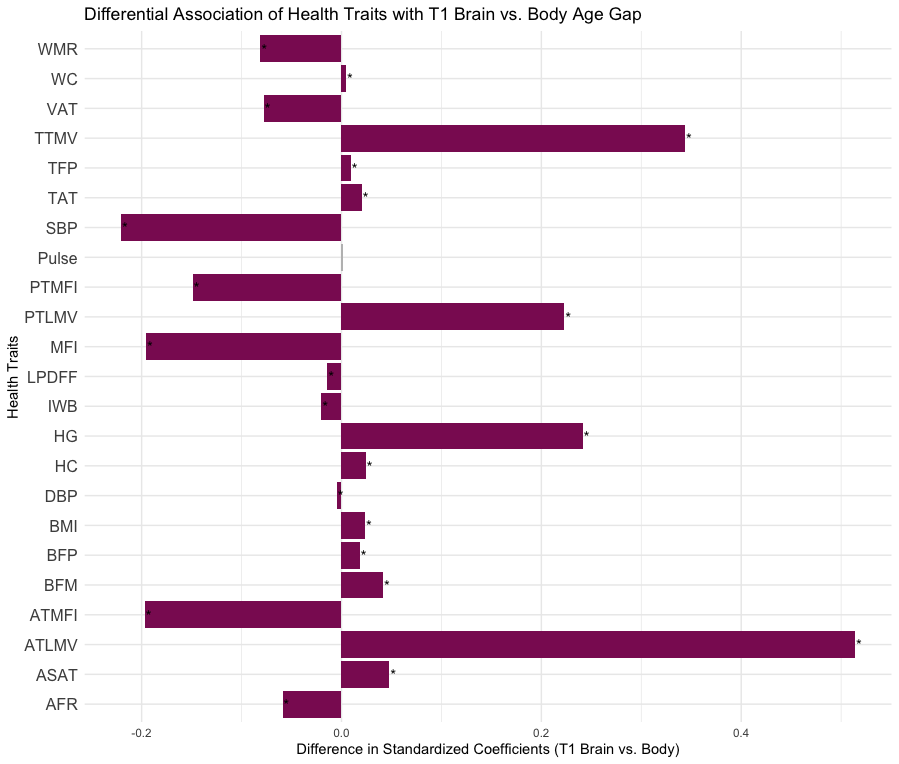

**SI Figure 14**. Results for a linear mixed effects model formula including difference score ~ health trait + Age + Sex (|ID).

**References**

de Lange, A.-M. G., Anatürk, M., Rokicki, J., Han, L. K. M., Franke, K., Alnæs, D., Ebmeier, K. P., Draganski, B., Kaufmann, T., Westlye, L. T., Hahn, T., & Cole, J. H. (2022). Mind the gap: Performance metric evaluation in brain-age prediction. *Human Brain Mapping*, *43*(10), 3113–3129. https://doi.org/10.1002/hbm.25837
